# RNA methyltransferase SPOUT1/CENP-32 links mitotic spindle organization with the neurodevelopmental disorder SpADMiSS

**DOI:** 10.1101/2024.01.09.23300329

**Authors:** Avinash V. Dharmadhikari, Maria Alba Abad, Sheraz Khan, Reza Maroofian, Tristan T. Sands, Farid Ullah, Itaru Samejima, Martin A. Wear, Kiara E. Moore, Elena Kondakova, Natalia Mitina, Theres Schaub, Grace K. Lee, Christine H. Umandap, Sara M. Berger, Alejandro D. Iglesias, Bernt Popp, Rami Abou Jamra, Heinz Gabriel, Stefan Rentas, Alyssa L Rippert, Kosuke Izumi, Laura K. Conlin, Daniel C. Koboldt, Theresa Mihalic Mosher, Scott E. Hickey, Dara V.F. Albert, Haley Norwood, Amy Feldman Lewanda, Hongzheng Dai, Pengfei Liu, Tadahiro Mitani, Dana Marafi, Davut Pehlivan, Jennifer E. Posey, Natalie Lippa, Natalie Vena, Erin L Heinzen, David B. Goldstein, Cyril Mignot, Jean-Madeleine de Sainte Agathe, Nouriya Abbas Al-Sannaa, Mina Zamani, Saeid Sadeghian, Reza Azizimalamiri, Tahere Seifia, Maha S. Zaki, Ghada M.H. Abdel-Salam, Mohamed Abdel-Hamid, Lama Alabdi, Fowzan Sami Alkuraya, Heba Dawoud, Aya Lofty, Peter Bauer, Giovanni Zifarelli, Erum Afzal, Faisal Zafar, Stephanie Efthymiou, Daniel Gossett, Meghan C. Towne, Raey Yeneabat, Sandeep N. Wontakal, Vimla S. Aggarwal, Jill A. Rosenfeld, Victor Tarabykin, Shinya Ohta, James R. Lupski, Henry Houlden, William C. Earnshaw, Erica E. Davis, A. Arockia Jeyaprakash, Jun Liao

**Author notes:** contributed equally (Avinash V. Dharmadhikari, Maria Alba Abad, Sheraz Khan). corresponding authors Correspondence to: Jun Liao A. Arockia Jeyaprakash, Erica E. Davis.

## Abstract

*SPOUT1/CENP-32* encodes a putative SPOUT RNA methyltransferase previously identified as a mitotic chromosome associated protein. SPOUT1/CENP-32 depletion leads to centrosome detachment from the spindle poles and chromosome misalignment. Aided by gene matching platforms, we identified 24 individuals with neurodevelopmental delays from 18 families with bi-allelic variants in *SPOUT1/CENP-32* detected by exome/genome sequencing. Zebrafish *spout1/cenp-32* mutants showed reduction in larval head size with concomitant apoptosis likely associated with altered cell cycle progression. *In vivo* complementation assays in zebrafish indicated that *SPOUT1/CENP-32* missense variants identified in humans are pathogenic. Crystal structure analysis of SPOUT1/CENP-32 revealed that most disease-associated missense variants mapped to the catalytic domain. Additionally, SPOUT1/CENP-32 recurrent missense variants had reduced methyltransferase activity *in vitro* and compromised centrosome tethering to the spindle poles in human cells. Thus, *SPOUT1/CENP-32* pathogenic variants cause an autosomal recessive neurodevelopmental disorder: SpADMiSS (*SPOUT1* Associated Development delay Microcephaly Seizures Short stature) underpinned by mitotic spindle organization defects and consequent chromosome segregation errors.

## INTRODUCTION

Error-free chromosome segregation is crucial for cell proliferation, tissue repair and organismal growth. The process requires the formation of a mitotic spindle, a bipolar structure formed by microtubules originating from a pair of separated centrosomes in somatic animal cells. Spindle microtubules interact with kinetochores formed on the centromeric region of chromosomes to ensure proper chromosome segregation during cell division. In most animal cells, centrosomes act as major microtubule organising centres (MTOCs) and form poles of a bipolar spindle where microtubules converge ^[1-3]^. Although higher plant cells and oocytes of many animals can assemble bipolar mitotic and meiotic spindles without centrosomes, centrosome removal in somatic animal cells typically results in mitotic delay and chromosome segregation errors ^[4, 5]^.

Multiple proteins and pathways are involved in mitotic spindle organization and microtubule dynamics. Centrosomes contain a pair of cylindrical centrioles surrounded by a matrix of pericentriolar material (PCM) that promotes efficient microtubule nucleation. In addition, chromatin-mediated and microtubule-mediated microtubule nucleation pathways have also been elucidated^[1],[6]^. Several PCM proteins recruit and anchor the γ-tubulin ring complex (γ-TuRC: a key nucleator of microtubule assembly) to centrosomes and also recruit microtubule nucleation effectors. As microtubules nucleate, they form a network that ultimately becomes the mitotic spindle. Bipolar spindle assembly is essential for the accurate and timely progression of mitosis ^[7]^.

Even though all dividing cells segregate chromosomes on a mitotic spindle, pathogenic variants in genes important for centrosome and spindle function, such as *CDK5RAP2* (MIM: 608201), *PCNT* (MIM: 605925), *WDR62* (MIM: 613583), and *ASPM* (MIM: 605481), cause a spectrum of human disorders that predominantly affect brain development ^[8-11]^. These disorders include autosomal recessive primary microcephaly, autosomal recessive microcephalic osteodysplastic primordial dwarfism type II (MOPD-II) and autosomal recessive Seckel syndrome ^[12]^. Despite their phenotypic variability, all these disorders share two common clinical features, reduction in cerebral cortex size and intellectual disability. Thus, the brain is particularly susceptible to defects affecting centrosome and mitotic spindle assembly and function. A proposed disease mechanism is that centrosome dysfunction causes spindle disorganization, chromosome segregation errors, and/or mitotic delay, that in turn lead to exhaustion of neural progenitor cells and defective brain growth by premature differentiation and cell death ^[13]^.

SPOUT1, also known as CENP-32 or C9ORF114, contains a putative SpoU-TrmD (SPOUT) RNA methyltransferase (MTase) domain. Its role in the centrosome and mitotic spindle was first discovered by a large-scale proteomics-based bioinformatics analysis of proteins associated with mitotic chromosomes ^[14]^. SPOUT1/CENP-32 localized to mitotic spindles and kinetochores, and its depletion by siRNA resulted in unusual centrosome detachment from the mitotic spindle poles, delayed anaphase onset, and chromosome segregation errors ^[14, 15]^. An independent large-scale CRISPR-Cas9 screen in human cells showed that SPOUT1/CENP-32 is essential for cell viability and is associated with RNA modification ^[16]^.

We report the association of bi-allelic *SPOUT1/CENP-32* variants with a complex neurodevelopmental disorder in 18 unrelated families involving 24 affected individuals. Common phenotypes in these individuals include microcephaly, seizures, intellectual disability, and varying degrees of developmental delays. These findings characterize a hitherto unreported autosomal recessive neurodevelopmental disorder (NDD); we term as SpADMiSS (*SPOUT1* Associated Development delay Microcephaly Seizures Short stature). Genetic ablation of *spout1/cenp-32* in zebrafish recapitulates phenotypes observed in humans; genetically stable mutant larvae display head size defects, augmented cell death, and altered cell cycle progression. Further, *in vivo* complementation assays demonstrate that missense *SPOUT1/CENP-32* changes identified in humans are pathogenic. *In vitro* studies demonstrate that the *SPOUT1/CENP-32* variants observed in affected individuals result in a reduction of the methyltransferase activity of the protein and compromise its function required for ensuring the tethering of centrosomes to the spindle poles in a human cell line. In summary, bi-allelic variants in *SPOUT1/CENP-32* cause an autosomal recesssive neurodevelopmental disorder SpADMiSS and highlight its roles in mitotic spindle organization and brain development.

## RESULTS

### Individuals with bi-allelic rare variants in *SPOUT1/CENP-32* display neurodevelopmental delays, seizures, microcephaly and short stature (SpADMiSS)

Through a global collaboration aided by GeneMatcher ^[17]^, we identified 24 individuals from 18 unrelated families harboring rare bi-allelic variants in *SPOUT1/CENP-32* (Supplementary case reports, Figure S1 and Table S1). These individuals included 14 females, 10 males, with ages ranging from 11-months to 20-years, and diverse ancestries including European, African American/Asian, German/Malaysian, Yemeni, Afghani, Egyptian, and Turkish, French, and Arab ethnicities (Saudi Arabian, Arab Iranian, and Syrian). Of note, family B has been previously reported in a cohort of 152 consanguineous families with NDDs.^[18]^

The *SPOUT1/CENP-32* variant spectrum in these families included 13 different missense variants, one frameshift variant, one nonsense variant and one in-frame deletion (Figure 1A; Table S2). The following variants were recurrent, p.N86D (2 families), p.G98S (8 families), p.G293S (2 families), and p.T353M (2 families). All variants were absent or rare in gnomAD. The majority of the variants occurred at amino acid residues of the protein conserved across species, and are predicted to be intolerant to variation, with 11/13 missense variants with scores >0.6 and predicted to be pathogenic by the recently described AlphaMissense predictor ^[19]^. (Figure 1B, Table S2). All affected individuals had bi-allelic variants with 10/18 families segregating homozygous variants whereas 8/18 pedigrees had compound heterozygous variants. Of the 10 families with homozygous variants, 7 were homozygous for the recurrent p.G98S variant. Sanger sequencing was performed for confirmation and segregation of *SPOUT1/CENP-32* variants in selected families as mandated by research or clinical workflows (Figure S2A).

**Figure 1:**
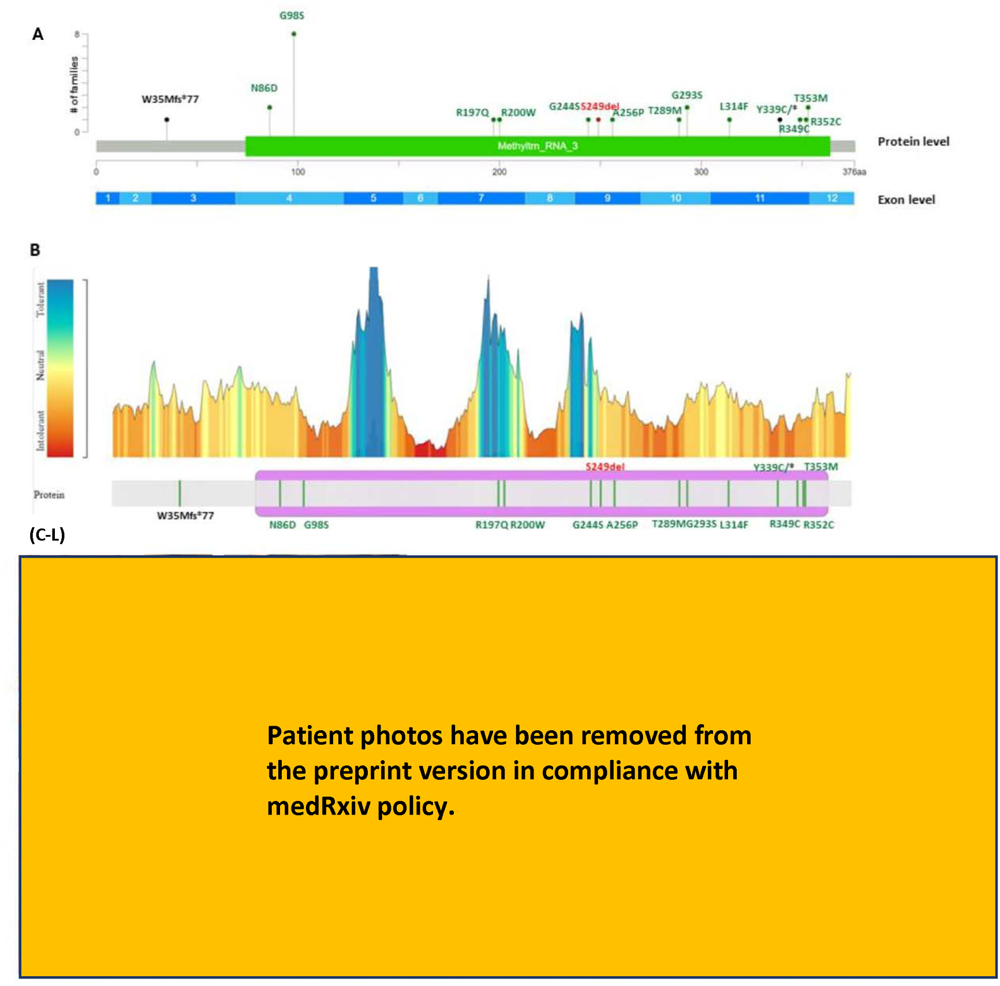
Majority of *SPOUT1/CENP-32* variants identified in affected individuals are located in the RNA methyltransferase domain of the protein and at amino acid residues intolerant to variation. (A) Schematic of *SPOUT1/CENP-32* at the exon level (NM_016390.4) and protein level (NM_057474.2) with positions of the variants identified in our cohort and number of alleles in unrelated families identified for each variant. Missense variants are displayed in green, loss-function variants in black and the in-frame deletion in red. The single annotated domain is from Pfam and reflects the RNA methyltransferase domain. (B) Overlay of SPOUT1/CENP-32 variants identified in this cohort with the protein intolerance landscape of SPOUT1/CENP-32 from MetaDome. MetaDome analyses the mutation tolerance at each position of the protein ^[99]^. (C-L) Variable dysmorphic features were seen in 58% (14/24) individuals including high arched palate, prominent ears, upturned nostrils, tented upper lip and high forehead.

Commonly observed clinical findings in individuals with bi-allelic *SPOUT1/CENP-32* variants included neurodevelopmental phenotypes of global developmental delay (DD) -100% (24/24), intellectual disability (ID) -100% (10/10), and seizures - 67% (16/24) (Table 1). Global development delays included motor delay, speech and language delay, and variable cognitive delays ranging from mild to severe ID. In many families, affected individuals were non-verbal and non-ambulatory. Measurements for height, weight, and head circumference were significantly lower than average in ∼ 80% of affected individuals (Table 1).

**Table 1:** Summary of clinical features in 24 individuals with bi-allelic *SPOUT1/CENP-32* variants. The most common features were developmental delay, intellectual disability, seizures, microcephaly, short stature, and failure to thrive. Additional findings included variable dysmorphic features, eye abnormalities, abnormal brain MRI findings, feeding issues, hypotonia, and hypertonia or spasticity. See Table S1 for further details.

| Clinical feature | Frequency in cohort (positive individuals/individuals evaluated, %) |
| --- | --- |
| Developmental delay | 24/24 (100%) |
| Intellectual disability | 10/10 (100%) |
| Epilepsy<br>-Epileptic encephalopathy (Lennox Gastaut syndrome and infantile spasms/hypsarrhythmia) | 16/24 (67%)<br>-8/24 (33%) |
| Poor weight gain | 17/18 (94%) |
| Short stature | 14/15 (93%) |
| Microcephaly | 15/18 (83%) |
| Variable dysmorphic features | 14/24 (58%) |
| Eye abnormalities* | 11/24 (46%) |
| Abnormal brain MRI findings | 14/24 (58%) |
| Feeding issues | 10/24 (42%) |
| Hypertonia/spasticity | 11/24 (46%) |
| Hypotonia | 8/24 (33%) |

\*Eye abnormalities included microphthalmia, nystagmus, slit-like eyes, leukocoria, and cataracts.

Electroencephalography (EEG) tracings were available for 5 individuals, which demonstrated a relatively consistent electroclinical phenotype, including disorganization in wakefulness, discontinuity in sleep, and abundant posterior-predominant sleep-potentiated spikes and polyspikes (Figure S2B). Close to a third, 29% (7/24) individuals met criteria for hypsarrhythmia. Typical seizure types included infantile epileptic spasms, tonic seizures, and focal-onset seizures. Abnormal MRI findings were found in 58% (14/24) individuals, with notable findings including cerebral atrophy with enlarged ventricles, T2 hyperintensity in the white matter and thinning of the corpus callosum (Figure S2C). These patients with early onset epilepsy and epileptic spams suggest the *SPOUT1* spectrum of neurodevelopmental phenotypes include developmental and epileptic encephalopathy.

Importantly, this constellation of features in the *SPOUT1/CENP-32* cohort overlaps with phenotypes seen in other “centrosome-based diseases” ^[20]^, caused by genes involved in centrosome or spindle function including *CDK5RAP2*, *PCNT*, *WDR62*, and *ASPM* ^[12]^.

### Zebrafish *spout1/cenp-32* depletion causes neuroanatomical defects

To determine whether loss of *SPOUT1/CENP-32* is causative for phenotypes observed in affected individuals, we generated zebrafish models of *spout1/cenp-32* by ablation (CRISPR/Cas9) or suppression (morpholino, MO). We and others reported previously that zebrafish is a robust model to investigate neurodevelopmental defects in humans ^[21-26]^. Reciprocal BLAST with the human SPOUT1/CENP-32 protein sequence (NP_057474.2) against the translated zebrafish genome identified a sole *SPOUT1/CENP-32* ortholog, encoding three *spout1/cenp-32* transcripts (canonical isoform; ENSDART00000099535.5; GRCz11; Figure S3A) for which the encoded protein has 76% identity and 88% similarity to human SPOUT1/CENP-32. Consistent with human expression data (GTEx; Human Protein Atlas), RNA *in situ* hybridization studies in zebrafish larvae showed *spout1/cenp-32* in a near ubiquitous pattern from the one-cell to pec-fin stages with a prominent expression pattern in the brain ^[27]^. Accordingly, *spout1/cenp-32* RNAseq data from zebrafish heads report abundant *spout1/cenp-32* transcript at 3 days post-fertilization (dpf) ^[28]^, and the zebrafish expression atlas ^[29]^ reports detectable transcript up to larval day 5. Thus, *spout1/cenp-32* expression in zebrafish is spatio-temporally appropriate to model early neurodevelopmental effects.

To determine the consequences of *spout1/cenp-32* depletion, we engineered genetically stable mutants using CRISPR/Cas9 genome editing using two single guide (sg) RNAs (sgRNA1 and sgRNA3) targeting exons 5 and 7 of the canonical isoform, respectively (Figure S3A and Table S3). We injected either sgRNA with or without recombinant Cas9 protein into single-cell stage embryos, and allowed them to develop until 2 dpf. Using a combined strategy of heteroduplex analysis, molecular cloning, and sequencing of PCR amplicons flanking the sgRNA target site, we estimated an average mosaicism of 81% for sgRNA1 and 76% for sgRNA3 in F0 crispants (Figure S3B, C). As a proxy for microcephaly and short stature, we acquired bright field lateral images of F0 zebrafish larvae at 3 dpf to measure the head size and body length. Similar to the clinical phenotypes exhibited by affected individuals with *SPOUT1/CENP-32* variants (Table 1; Table S1), mosaic F0 mutants displayed a significant reduction in head size area (Figure S4A-D).

Next, we outcrossed F0 to WT adult zebrafish and isolated animals harboring a frameshifting 1 bp insertion allele in exon 7 (c.543_544insA; p.R215Kfs*7); these F1 heterozygotes were incrossed and F2 offspring were used for subsequent phenotyping (WT denoted as ‘+/+’; heterozygous as ‘+/-’; and homozygous mutant as ‘-/-‘). Consistent with F0 crispant data (Figure S4A-F), *spout1/cenp-32* stable mutants exhibited a significantly reduced head size (20% reduction, q<0.0001 for *spout1/cenp-32*^-/-^ vs *spout1/cenp-32*^+/+^); with modest but significant reduction in body length (Table S4; Figure 2A-C). Using qPCR to monitor mRNA expression in *spout1/cenp-32* mutant heads compared to sibling larvae, we observed a ∼3-fold depletion of *spout1/cenp-32* mRNA in mutants compared to WT counterparts (Figure 2D). Notably, we observed expected Mendelian ratios, but *spout1/cenp-32* homozygous mutants die by 8 dpf (n=3 independent clutches; n=300 F2 larvae including all genotypes). Together, these data support the involvement of *spout1/cenp-32* in the early development of anterior structures.

**Figure 2:**
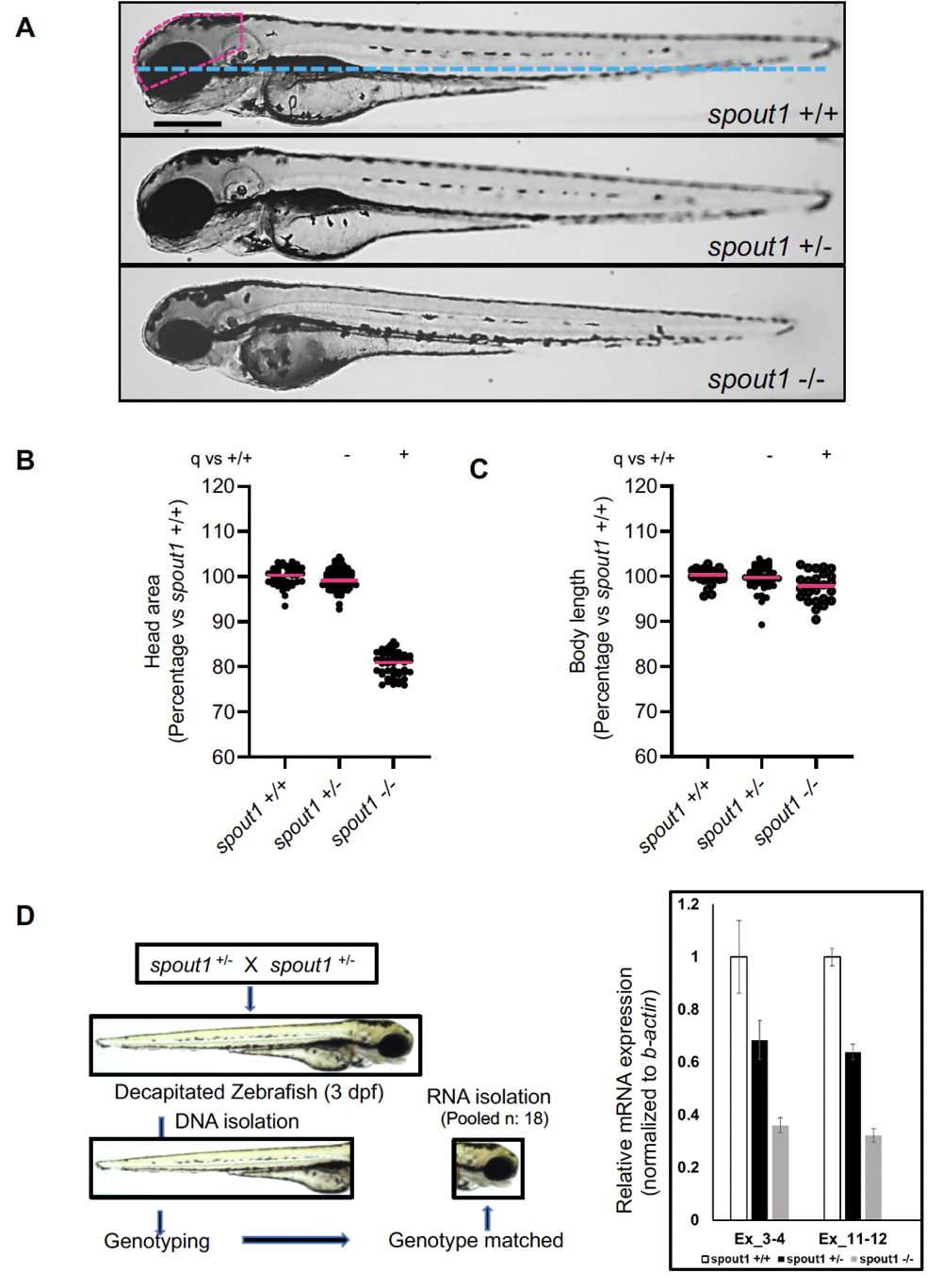
*spout1/cenp-32* mutant larvae display reduced head area. (A) Representative bright field lateral images of *spout1/cenp-32*^+/+^, *spout1/cenp-32*^+/-^ and *spout1/cenp-32*^-/-^ are shown at 3 days post-fertilization (dpf); pink dashed outline depicts head size measured, and the blue dotted line shows the body length measured. (B-C) Quantification of lateral head size (left) and body length (right) measurements were analyzed from combined experimental batches (n=2 biological replicates). (D) Schematic representation (left) of sample preparation for qPCR to monitor endogenous *spout1/cenp-32* transcript (right); error bars show standard deviation. Scale bar, 300 µM. Statistical differences were calculated using non-parametric ANOVA with Kruskal-Wallis test followed by Dunn’s multiple comparisons test by controlling False Discovery Rate (original FDR method of Benjamini and Hochberg); (+) and (-) indicate significant and non-significant differences, respectively. Median values are shown with pink horizontal lines. See Table S4 for exact adjusted q-values and numbers of larvae.

To validate the specificity of our stable mutant and F0 crispant phenotype data and to complement our observations with an independent method, we synthesized a splice-blocking morpholino (sbMO) targeting the splice donor site of *spout1/cenp-32* exon 4 in the canonical transcript (e4i4; Figure S3A; Table S3). We determined the efficiency of the sbMO by RT-PCR on total RNA harvested from MO-injected embryos at 2 dpf. The MO induced retention of intron 4, resulting in a premature termination codon and frameshift (Figure S3D, E). We injected e4i4 MO into WT embryos at increasing doses (3 ng, 6 ng, and 9 ng), and performed live imaging for morphometric analysis using the same paradigm as stable mutants or F0 crispants. We observed a dose-dependent decrease in the head area in comparison to uninjected controls (q<0.0001 vs UC; Figure S5A, B; Table S4). Importantly, the head size phenotype was rescued by co-injecting WT human *SPOUT1/CENP-32* mRNA (q<0.0001 vs MO; Figure 3A, B; Figure S5C, D; and Table S4). Thus, two independent approaches reveal that *spout1/cenp-32* reduction results in neuroanatomical deficits.

**Figure 3.**
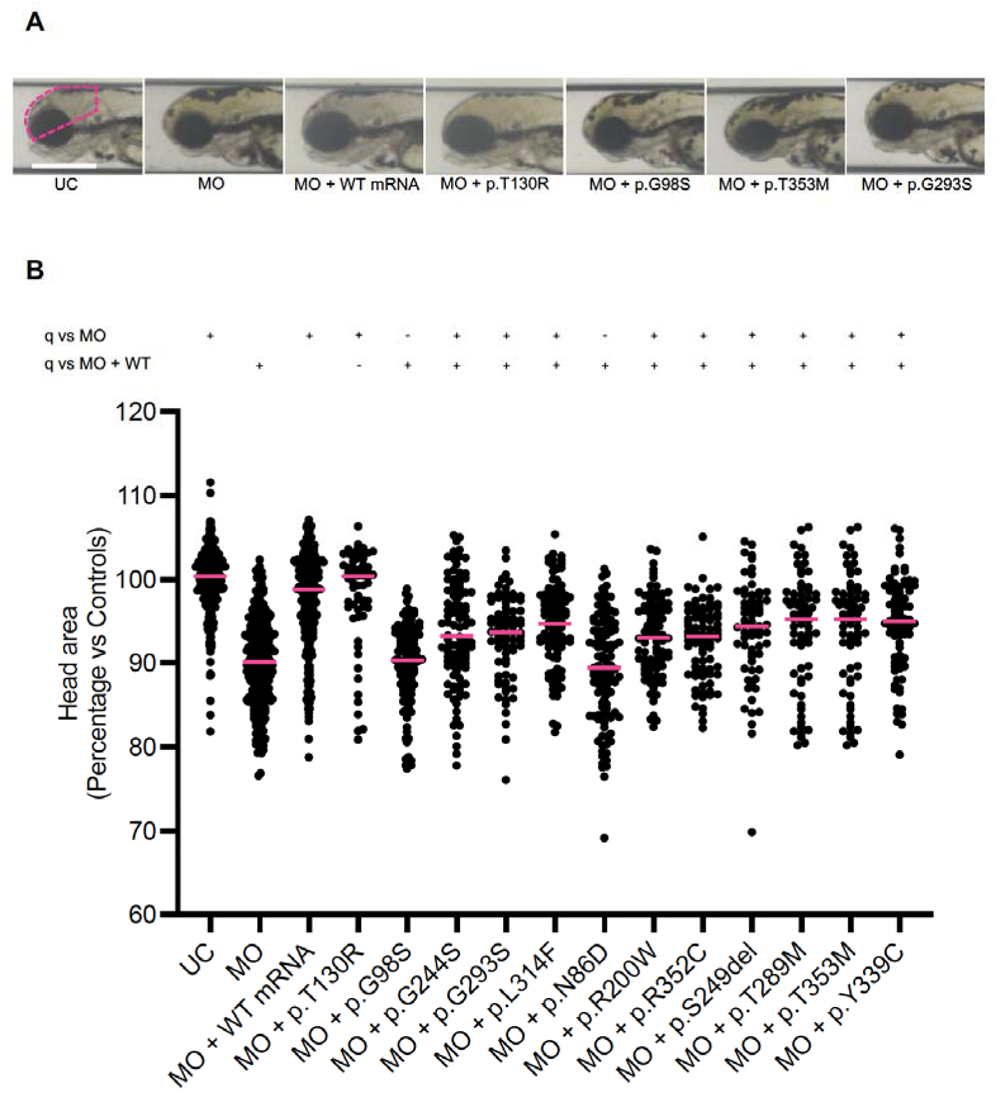
*In vivo* complementation assay indicates that variants identified in affected individuals are pathogenic. (A) Representative bright field images of uninjected control (UC, morphants (MO), and MO plus mRNA-injected larvae at 3 dpf. (B) *In vivo* complementation data show that variants identified in affected individuals are pathogenic. p.T130R, rs6478854 is a presumed benign variant used as a negative control. Scale bar, 300 µM. Statistical differences were calculated using non-parametric ANOVA with Kruskal-Wallis test followed by Dunn’s multiple comparisons test by controlling False Discovery Rate (original FDR method of Benjamini and Hochberg); (+) and (-) indicate significant and non-significant differences, respectively. Median values are shown with pink horizontal lines. See Table S4 for exact adjusted q-values and numbers of larvae.

To probe further whether *spout1/cenp-32* suppression perturbs specific anterior structures, specifically CNS defects of the affected individuals in our cohort (microcephaly and thin corpus callosum); we painted axon tracts with anti-α-acetylated tubulin immunostaining in zebrafish larvae at 3 dpf. Quantification of optic tecta size and number of commissural axon tracts are established proxies in zebrafish for microcephaly and corpus callosum anomalies, respectively ^[22, 24, 30]^. Consistent with our bright field lateral head size measurements, quantification of dorsal fluorescent signal of both optic tecta showed significant reduction in size between *spout1/cenp*-*32*^-/-^ compared to their WT or heterozygous siblings (q<0.0001 vs WT; Figure 4A, B; and Table S4). Further, we counted fewer commissural axons crossing the midline between optic tecta for *spout1/cenp-32*^-/-^ compared to WT or heterozygous siblings (q<0.0001 vs UC; Figure 4A, C, and Table S4). CNS defects observed in *spout1/cenp-32* mutants were consistent with our transient suppression model, and co-injection of WT *SPOUT1/CENP-32* mRNA rescued both CNS abnormalities as compared to MO alone (q<0.0001 vs MO; Figure S6A, C, and Table S4).

**Figure 4.**
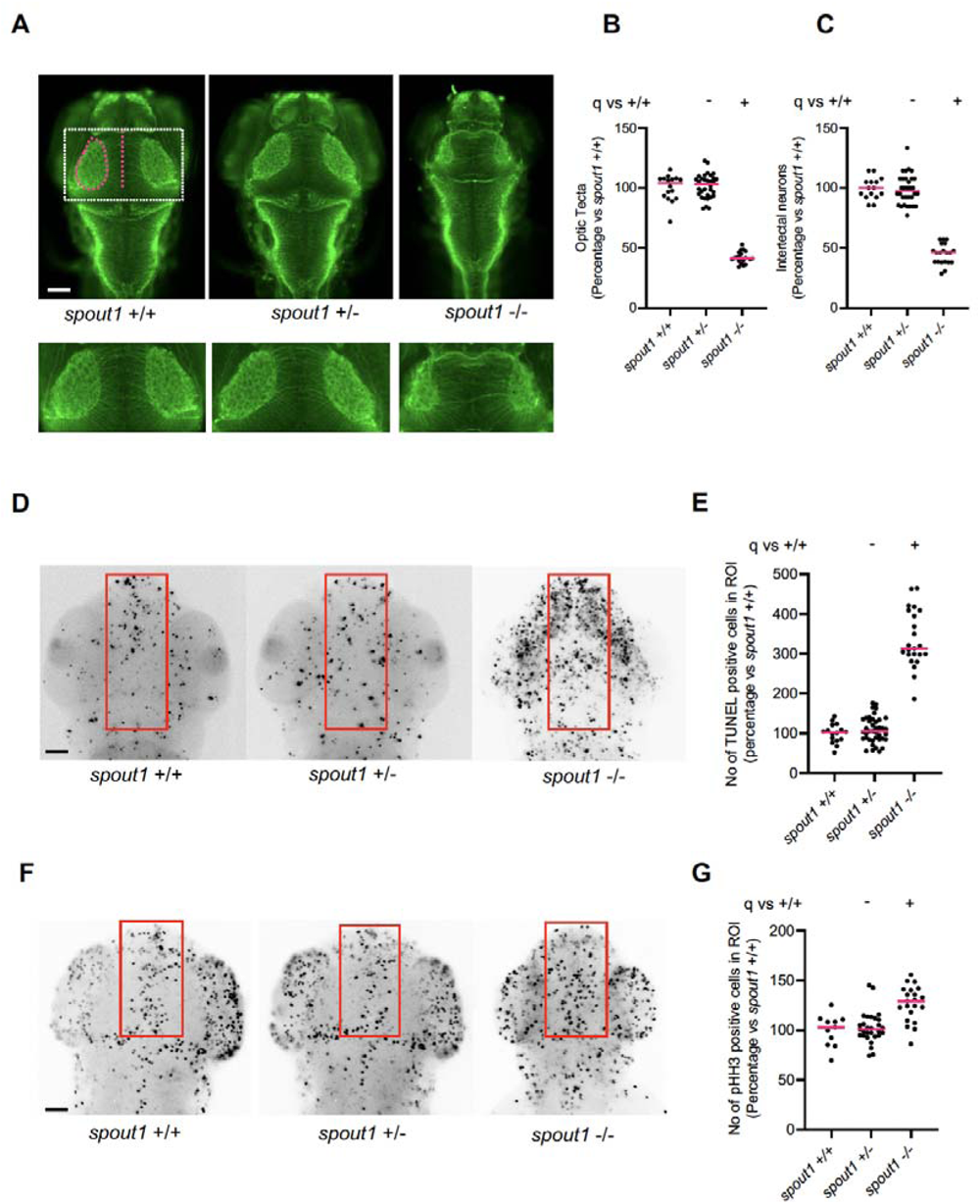
*spout1/cenp-32^-/-^*mutant larvae have CNS defects, apoptosis and altered cell cycle progression. (A) Representative dorsal images of wholemount larvae fixed at 3 dpf and immunostained for acetylated tubulin to demarcate axon tracts. We counted commissural axons crossing the midline between the optic tecta (pink dashed line); and the area of optic tecta (pink dashed oval). Dashed white box (upper panel) represents magnified image in lower panel. Scale bar, 100 µM. (B,C) Quantification of optic tecta size and intertectal neuron number, respectively. (D) Representative dorsal inverted fluorescent images of zebrafish larvae marked by TUNEL at 2 dpf. The region of interest (ROI) quantified is shown with a pink rectangle. Scale bar, 100 µM. (E) Quantification of TUNEL stained cells as measured in the ROI. (F). Representative dorsal inverted fluorescent images showing phospho-histone H3 (pHH3) positive cells at 2 dpf. The ROI quantified is shown with a pink rectangle. Scale bar, 100 µM. (G) Quantification of pHH3 positive cells as measured in ROI. Data shown in B, C, E, and G are combined from two biological replicates. Statistical differences were calculated using non-parametric ANOVA with Kruskal-Wallis test followed by Dunn’s multiple comparisons test by controlling False Discovery Rate (original FDR method of Benjamini and Hochberg); (+) and (-) indicate significant and non-significant differences, respectively. Median values are shown with pink horizontal lines. See Table S4 for exact adjusted q-values and numbers of larvae.

### *spout1/cenp-32* depletion *in vivo* causes increased apoptosis and prolonged cell cycle

Pathognomonic neuroanatomical defects have been associated with apoptosis and/or cell cycle arrest in the developing zebrafish larval head ^[21], [23], [26, 31-33]^. *SPOUT1/CENP-32* depletion in human cell lines leads to centrosome detachment from the spindle pole and chromosome misalignment ^[14, 15]^. Additionally, mice lacking the overlapping *Spout1/Cenp-32* and *EndoG* loci show abnormal male germ cell apoptosis and embryonic lethality before implantation^[34]^(MGI: 106544). Based on these data combined, we hypothesized that the decreased head size in zebrafish *spout1/cenp-32* models could be due to altered cell cycle progression with concomitant cell death.

Indeed, we observed increased cell death in *spout1/cenp-32* stable mutants compared to WT siblings (q<0.0001 *spout1/cenp-32^-/-^* vs WT) at 2 dpf when we performed whole mount TUNEL staining and quantification of positive cells within a region of interest of the dorsal forebrain between the eyes (Figure 4D,E; Table S4). Monitoring cell cycle progression using whole mount pHH3 (histone H3-S10ph) staining revealed a significant increase in pHH3-positive cells in a defined dorsal region of the head (q<0.0001, *spout1/cenp-32^-/-^*vs WT; Figure 4F, G, and Table S4). In independent experiments, *spout1/cenp-32* morphants showed concordant apoptosis and altered cell cycles progression defects (q<0.0001 vs MO; Figure S6D-G, and Table S4). Together, these data support the possibility that diminished *SPOUT1/CENP-32* results in altered cell cycle progression that may result in apoptosis. In order to confirm the zebrafish findings in a mammalian neural system and confronted with the embryonic lethality preceding implantation resulting from knockout of the overlapping *Spout1/Cenp-32* and *EndoG* loci^[34]^, we used a transient knock-down strategy for knocking out *Spout1/Cenp-32* in mice. Our results showed that down-regulation of *Spout1* in cortical stem cells in mice causes disruption of cell cycle progression (Figure S7, Figure S8). These results indicate that depletion of *Spout1* in neural cells leads to increased apoptosis with concomitant defects in cell cycle progression.

### *SPOUT1/CENP-32* variants identified in humans are pathogenic according to zebrafish *in vivo* complementation assays

To bolster the evidence that the rare *SPOUT1/CENP-32* coding variants in our human cohort are pathogenic, we performed *in vivo* complementation assays on eleven variants ^[30, 35, 36]^. We co-injected *spout1/cenp-32* sbMO with human *SPOUT1/CENP-32* mRNA harboring each disease-associated variant (p.G98S, p.T353M, p.N86D, p.G293S, p.R200W, p.R352C, p.G244S, p.L314F, p.Y339C, p.T289M and p.S249del), and compared the rescue efficiency to WT human *SPOUT1/CENP-32* mRNA using head size as a phenotypic readout. We also tested p.T130R, a common variant in gnomAD, expected to be benign (dbSNP ID: rs6478854; 16,933 homozygotes of 130,923 individuals, negative control). We found that mRNA encoding disease-associated variants p.G98S or p.N86D failed to rescue the MO-induced head size phenotype, and were indistinguishable from morphants, suggesting a loss of function (Figure 3A, B; Figure S10 and Table S4). Further, we observed partial rescue of the phenotype in larvae co-injected with MO and mRNAs encoding each of p.G244S, p.G293S, p.R200W, p.R352C, p. L314F, p.S249del, p.T289M, p.T353M, and p.Y339C (Figure 3A, B; Figure S10 and Table S4). Notably, all three recurrent variants (p.G98S, p.G293S and p.T353M) scored as pathogenic and p.T130R scored as benign, supporting the sensitivity and specificity of our assay. We did not detect marked differences in body length phenotype in the *in vivo* complementation assays (Figure S9A, B; Figure S10; and Table S4). Additionally, ectopic expression of WT or mutant *SPOUT1/CENP-32* mRNA in the absence of sbMO was indistinguishable from un-injected controls (Figure S11A, B; Table S4).

Together, our zebrafish modeling studies support a critical role for *SPOUT1/CENP-32* in neurodevelopment and confirm the pathogenicity of eleven of the missense variants in our cohort.

### Structural analysis of SPOUT1/CENP-32 reveals a SAM-dependent SPOUT methyltransferase domain suitable for catalytic activity

In order to determine a molecular mechanism for the pathogenicity of the variants seen in our patient cohort and confirmed by *in vivo* studies using zebrafish, we characterized the structure of the SPOUT1/CENP-32 protein (Figure 5). This protein has a predicted SPOUT domain with an oligonucleotide binding (OB) fold inserted into the catalytic domain (Figure 5A and Figure S12A). It also contains a highly basic N-terminal region predicted to form an α-helix (amino acid residues 20-64; pI = 9.48; Figures 5A and S12A). To gain insights into SPOUT1/CENP-32 structure and function, we crystallised the predicted methyltransferase (MTase) domain (SPOUT1/CENP-32 71-376) made recombinantly in *E. coli* (Figure S13A). The crystal structure was determined to a resolution of 2.38 Å using Single Anomalous Dispersion (SAD) phasing using Seleno (Se) -methionine labelled SPOUT1/CENP-32 (Figure 5B and Table S5).

**Figure 5:**
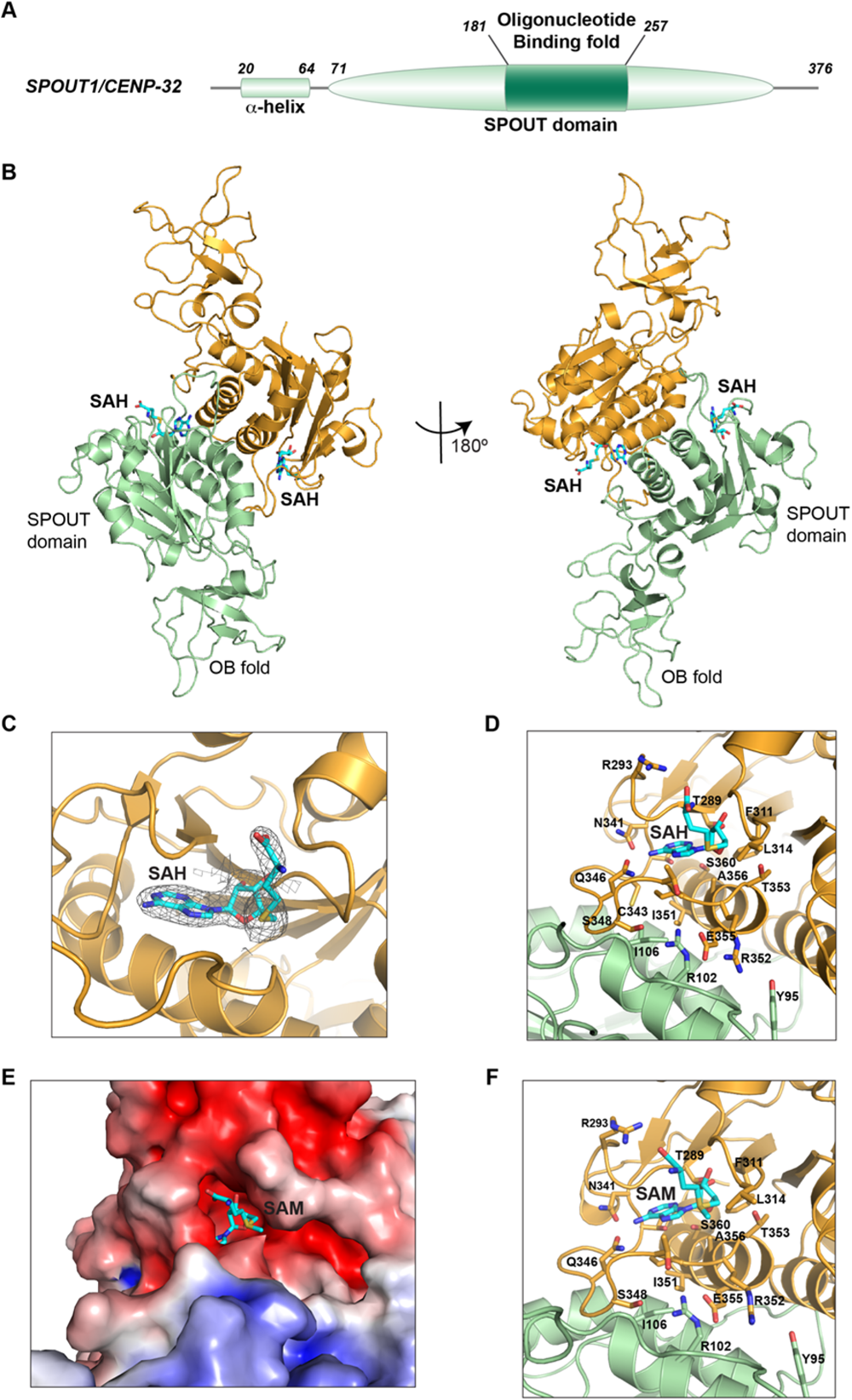
Structural characterisation reveals that SPOUT1/CENP-32 is a SPOUT Methyltransferase that dimerizes through its catalytic domain. (A) Domain architecture of SPOUT1/CENP-32 with the structural domains of SPOUT1/CENP-32 highlighted. (B) Cartoon representation of the crystal structure of SPOUT1/CENP-32 71-376 bound to SAH in two orientations (left and right panel). The main domains of SPOUT1/CENP-32 (SPOUT domain and OB fold) are highlighted. S-adenosyl homocysteine (SAH) is bound to the cofactor pocket, indicating that SPOUT1/CENP-32 could be an active methyltransferase. Our structural analysis is consistent with the structure that was deposited by SGC while this manuscript was in preparation (PDB: 4RG1). (C, D and F) Close-up of the active site of SPOUT1/CENP-32 with SAH (C and D) and with SAM (F) showing the electron density map for SAH (C) and the amino acid residues that are responsible for the interactions that stabilise cofactor binding. The binding of SAH and SAM is similar. (E) Close-up of the electrostatic surface potential of SPOUT1/CENP-32 71-end bound to SAM showing the deep pocket formed by the trefoil knot.

Structural analysis confirmed that SPOUT1/CENP-32 is a SPOUT MTase with a trefoil knot fold consisting of parallel six stranded β-sheets sandwiched between two layers of α-helices (Figure 5B) that promote dimerization of the protein. A common feature of SPOUT MTases is the presence of additional domains fused to their N-or C-terminus or inserted between domains within the SPOUT MTase. The SPOUT1/CENP-32 OB fold ^[37-39]^ is inserted into the SPOUT MTase domain (amino acid residues 181 to 257) (Figure 5B and S12A). Like the OB fold in SPOUT1/CENP-32, most of these additional domains in other SPOUT MTases are involved in nucleic acid recognition, suggesting that SPOUT methyltransferases are mostly involved in nucleic acid binding and methylation ^[40]^. Notably, SPOUT1/CENP-32 is the only SPOUT superfamily member with the OB fold insertion within the catalytic domain ^[40]^.

SPOUT MTases use S-adenosyl-L-methionine (SAM) as the methyl group donor for the methyltransferase reaction. The products of this reaction are the methylated substrate and S-adenosyl-homocysteine (SAH) ^[40, 41]^. Although we did not supplement SPOUT1/CENP-32 with either SAM or SAH during protein purification or crystallisation, our crystal structure showed SAH bound within the cofactor binding site (Figure 5B-D). This strongly suggests that SPOUT1/CENP-32 is an active MTase that stably retains the SAH by-product of its methyltransferase reaction (Figure S13B).

The cofactor binding site is a deep pocket formed between the six-stranded beta-sheet and the helices involved in dimerization. SAH is stabilised via extensive hydrophobic and hydrogen bonding interactions involving the adenine and ribose moieties of the SAH and catalytic site residues (Figure 5D). To determine whether SPOUT1/CENP-32 can also bind the cofactor, SAM, and to gain further structural insights into the substrate binding site, we soaked the SPOUT1/CENP-32 crystals in a cryo-protectant solution containing a molar excess of SAM prior to freezing the crystals for X-ray data collection. These crystals diffracted X-rays beyond 2.62 Å (Figure 5E, 5F and S12C). The SAM-bound structure of SPOUT1/CENP-32 was determined using molecular replacement. The difference electron density map calculated for the refined model showed a well-defined electron density for the bound SAM (Figure S13C). The mode of SAM binding is similar to that of SAH with a methyl group pointing out of the deep cofactor binding pocket (Figure 5D-F).

Our SPOUT1/CENP-32 structures also reveal that SPOUT1/CENP-32 dimerises via a helical segment in the catalytic domain that packs almost perpendicularly against an equivalent segment from its dimeric counterpart (Figure 5B and S12B). This mode of dimerization stabilises the conformation of the substrate binding pocket and hence is likely crucial for the enzymatic activity. To ensure that SPOUT1/CENP-32 dimerization is not an artifact of crystal packing and to test whether SPOUT1/CENP-32 is a homodimer in solution we performed size exclusion chromatography combined with multi-angle light scattering (SEC-MALS). The molecular weight measured via SEC-MALS is 66.06 ± 1.26 kDa, which matches with a calculated molecular weight for a homodimeric assembly (67 kDa) (Figure S12C). This is consistent with reports that all studied SPOUT family members are known to form dimers, with dimerization suggested to be essential for MTase activity ^[40]^.

### SPOUT1/CENP-32 is an active RNA MTase

SPOUT MTases are the second largest family of RNA MTases, with most members involved in RNA metabolism and ribosome biosynthesis ^[42]^. The crystal structure suggests that SPOUT1/CENP-32 is likely an active enzyme, so we evaluated its activity by performing *in vitro* MTase-Glo^TM^ assays (Promega) ^[43]^ using recombinant SPOUT1/CENP-32. Recombinant NSUN6, a well characterised MTase ^[44]^ and Survivin, a protein with no nucleic acid binding activity, were used as positive and negative controls, respectively (Figure 6A). The bioluminescence signal obtained in the MTase assay directly corresponds to the amount of SAH produced in the reaction. Our results reveal that SPOUT1/CENP-32 has intrinsic activity that is comparable to the MTase activity of NSUN6 for total RNA extract (Figure 6A). As expected, Survivin showed no MTase activity (Figure 6A). This, for the first time, demonstrates that SPOUT1/CENP-32 retains enzymatic activity towards RNA.

**Figure 6:**
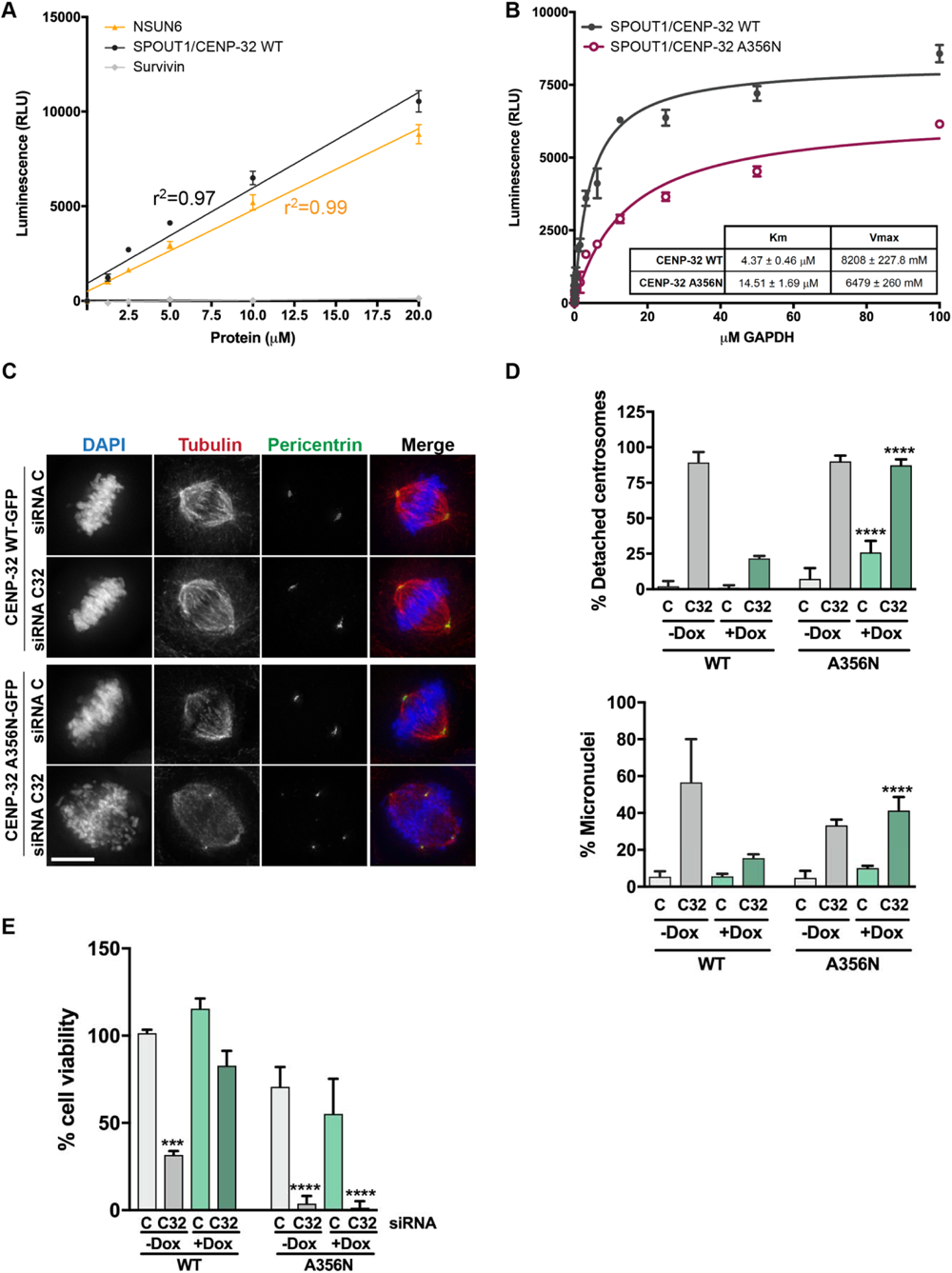
SPOUT1/CENP-32 is an active Methyltransferase and its activity is crucial for its mitotic function. (A) Titration of methyltransferases (SPOUT1/CENP-32 and NSUN6) to assess the methyltransferase activity of SPOUT1/CENP-32 *in vitro* were Survivin protein was used as a negative control. Increasing amounts of either methyltransferase (NSUN6, a well-known RNA methyltransferase, or SPOUT1/CENP-32) or Survivin protein (0 -20 μM) were incubated with 1 μg of total RNA extract (isolated from NSUN6 depleted or SPOUT1/CENP-32 depleted U2OS cells) and 20 μM SAM for 30 min at room temperature. RLU (Relative Luminescence unit). Data represented as mean ± SEM. (B) Titration of SPOUT1/CENP-32 WT and A356N mutant. Increasing amounts of SPOUT1/CENP-32 proteins (0 - 100 μM) were incubated with 1 μg of total RNA extracts (isolated from SPOUT1/CENP-32 depleted U2OS cells) and 20 μM SAM for 30 min at room temperature. RLU (Relative Luminescence unit). Data represented as mean ± SEM. (C) Representative immunofluorescence images of SPOUT1/CENP-32 inducible U2OS cell lines for the analysis of centrosome detachment upon SPOUT1/CENP-32 (C32) depletion using siRNA oligos and rescue with either induction of SPOUT1/CENP32 WT-GFP or SPOUT1/CENP32 A356N-GFP expression. Conditions with doxycycline only. Conditions without doxycycline in Figure S14B. (D) Quantification for the analysis of the centrosome detachment phenotype (% of cells with detached centrosomes; top panel) and chromosome segregation errors (% of micronuclei; bottom panel) in inducible U2OS cell lines expressing SPOUT1/CENP-32 WT-GFP or SPOUT1/CENP-32 A356N-GFP. Data are representative of a minimum of three biological replicates, mean ± SD. Scale bar, 10 μm; Fisher’s exact test with Bonferroni correction (****, P ≤ 0.0001). (E) Cell viability evaluation of inducible U2OS cell lines expressing SPOUT1/CENP-32 WT-GFP or SPOUT1/CENP-32 A356N-GFP assessed by the MTT assay. Three independent experiments, mean ± SEM. Non-parametric ANOVA with Kruskal-Wallis followed by Dunn’s multiple comparisons test (***, P ≤ 0.001; ****, P ≤ 0.0001).

To assess if the observed activity is specific and depends on SPOUT1/CENP-32’s ability to bind its cofactor SAM, we mutated Ala356, an amino acid residue located at the centre of the SAM binding pocket, to Asn (an amino acid residue with a relatively longer side chain; A356N) and characterised this variant by determining its crystal structure and by subjecting it in our MTase activity assay (Figure S13D and Figure 6B). The crystal structure of SPOUT1/CENP-32 A356N determined at 2.5 Å resolution, showed that while this variant does not affect the overall structure or the conformation of SAM binding pocket, the variant protein failed to stably bind SAH in the expression host (*E. coli*), confirming the perturbation of cofactor binding (Figure S13B-D). We tested the SPOUT1/CENP-32 A356N variant in our MTase activity assays and observed that this variant shows reduced activity indicating that the specific catalytic activity of SPOUT1/CENP-32 depends on cofactor binding (Figure 6B and Figure S14A).

### Catalytic activity of SPOUT1/CENP-32 is necessary for centrosome tethering to the spindle poles

Consistent with previous studies ^[14, 15]^, depletion of SPOUT1/CENP-32 using siRNA oligos results in cells showing a unique centrosome detachment phenotype with more than 80% of the cells showing centrosomes detached from spindle poles of bipolar spindles and often migrating towards the metaphase plate (Figure 6C, 6D, S13E, S13F and S14B). Expression of wild type (WT) SPOUT1/CENP-32-GFP in U2OS stable cell lines significantly rescues this phenotype when endogenous SPOUT1/CENP-32 is depleted (Figure 6C, 6D, S13G and S14B).

Incorrectly segregated chromosomes often result in the formation of micronuclei. SPOUT1/CENP-32 depletion leads to an increase in the percentage of micronucleus formation that can be rescued with induction of SPOUT1/CENP-32 WT-GFP expression, indicating that SPOUT1/CENP-32 is necessary for accurate chromosome segregation (Figure 6D). We tested the A356N catalytic variant in our siRNA rescue assays to check whether the methyltransferase activity of SPOUT1/CENP-32 is necessary for its function. Depletion of SPOUT1/CENP-32 and induction of SPOUT1/CENP32 A356N-GFP expression in our stable cell lines failed to rescue the centrosome detachment and micronucleus formation phenotype (Figure 6C, 6D, S13E-G, and S14B). Thus, SPOUT1/CENP-32 methyltransferase activity is required for maintaining spindle integrity (Figure 6D).

We also assessed the effect of SPOUT1/CENP-32 depletion on cell proliferation using the MTT (3-(4,5-Dimethylthiazol-2-yl)-2, 5-Diphenyltetrazolium bromide) colorimetric assay. Consistent with our zebrafish cell death data (Figure 4D-E; Figure S6D-E), SPOUT1/CENP32 depletion in U2OS cells led to a decrease in cell viability (31.7 ± 2.3 % for siRNA C32 vs 101.5 ± 1.9 % for siRNA C; Figure 6E). As expected, we also observed reduced cell viability for cells expressing SPOUT1/CENP32 catalytic variant (SPOUT1/CENP32 A356N-GFP) with reduced methyltransferase activity (Figure 6E).

### Disease-associated SPOUT1/CENP-32 missense variants show reduced methyltransferase activity

To understand the effect of the SPOUT1/CENP-32 patient variants on the activity and function of SPOUT1/CENP-32, we first mapped the amino acid variations onto the SPOUT1/CENP-32 crystal structure (Figure 7A). Several of these variants are clustered around the cofactor binding pocket (N86D, T289M, G293S, L314F, Y339C, R352C and T353M) while others are present at the dimeric interface (G98S) and within the OB fold (R200W, G244S and S249del) (Figure 7A). Considering the proximity of the variants to the catalytic site and the nature of the amino acid changes, and recurrence in affected individuals, we assessed the effect of N86D, G98S, T289M, G293S and T353M missense variants on the catalytic activity of SPOUT1/CENP-32. We also assessed the G244S missense variant as this change is located in the OB fold and might affect substrate binding. We used SPOUT1/CENP-32 T130R, a common variant expected to be benign and to behave like the WT protein, as a negative control.

**Figure 7:**
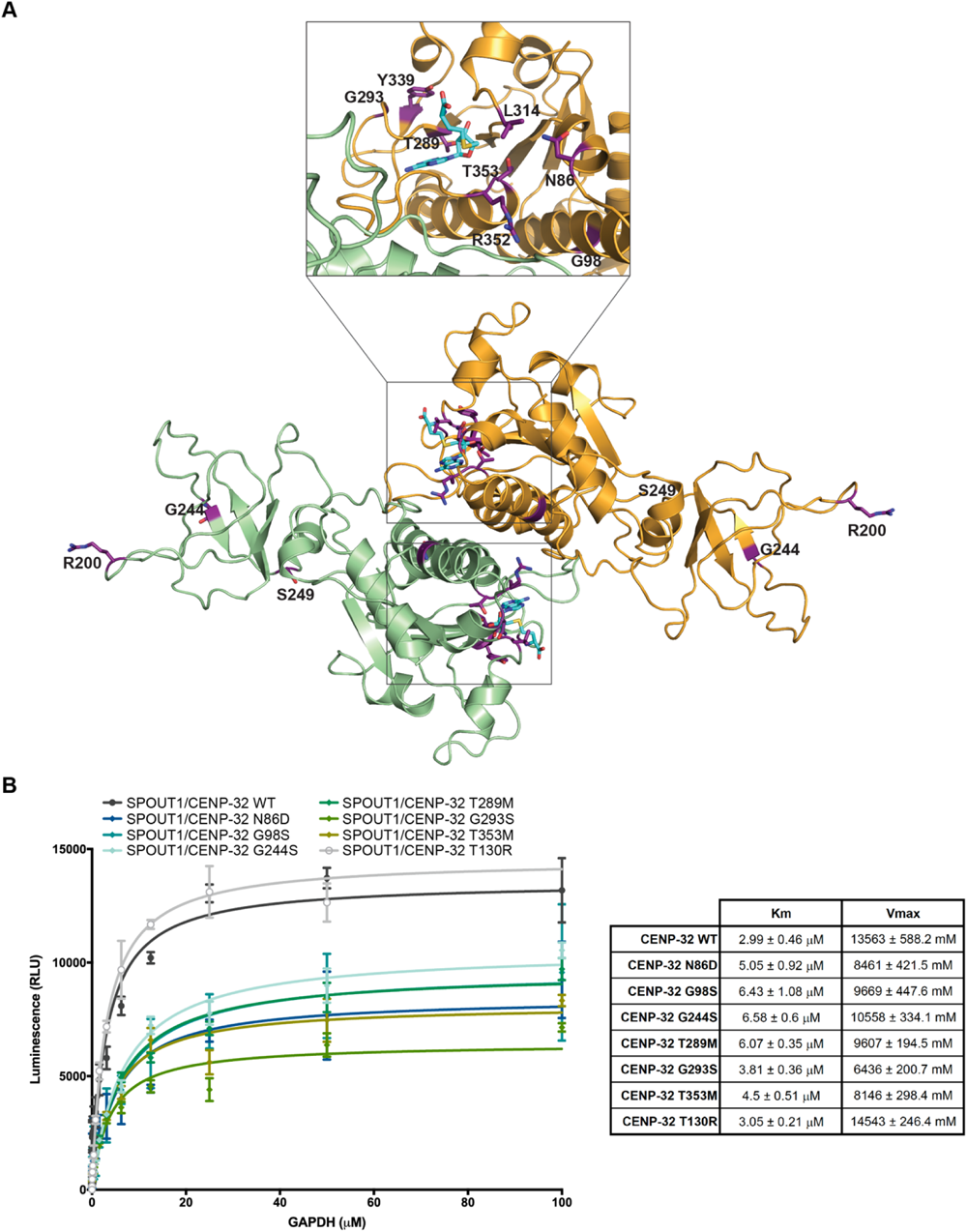
SPOUT1/CENP-32 patient variants show decreased methyltransferase activity. (A) Mapping of the SPOUT1/CENP-32 variants in the SPOUT1/CENP-32 71-376 SAH-bound structure. Variant residues are highlighted in purple. (B) Methyltransferase assay to determine the Km and Vmax of SPOUT1/CENP-32 WT and the variants in the presence of constant SAM (40 μM) and increasing amounts of a *GAPDH* mRNA hairpin as substrate (GCCCCCUCUGCUGAUGCCCCCAU GUUCGUCAUGGGUGUGAA; 0 – 100 μM; left panel). Km and Vmax values for all SPOUT1/CENP-32 proteins are depicted in the table (right panel). R square values are as follows: SPOUT1/CENP-32 WT – 0.79, SPOUT1/CENP-32 N86D – 0.83, SPOUT1/CENP32 G98S – 0.87, SPOUT1/CENP-32 G244S – 0.93, SPOUT1/CENP-32 T289M – 0.97, SPOUT1/CENP-32 T353M – 0.9, SPOUT1/CENP-32 T130R – 0.98).

We assessed the MTase activity of the selected SPOUT1/CENP-32 patient variants in our *in vitro* MTase activity assays. It has been shown previously that SPOUT1/CENP-32 recognises secondary structures of RNA, specifically to the RNA stem [45]. We used the hairpin formed by a 41nt sequence of the *GAPDH* mRNA as a substrate and indeed, including this substrate in the assay stimulated the catalytic activity of SPOUT1/CENP-32 WT. The SPOUT1/CENP-32 variants seen in patients show a 2-fold increase in Km and/or a decrease in the Vmax, while T130R showed comparable Km and Vmax to SPOUT1/CENP-32 WT. Thus, we conclude that substrate binding and turnover of the SPOUT1/CENP-32 variants are affected, making them less efficient enzymes (Figure 7B and S14A) in affected individuals.

### SPOUT1/CENP-32 missense variants affect the role of SPOUT1/CENP-32 in maintaining spindle integrity

To further understand whether the decrease in MTase activity observed for SPOUT1/CENP-32 variants is linked to the role of SPOUT1/CENP32 in ensuring proper spindle organization, we performed siRNA rescue assays (Figure 8 and S14C). SPOUT1/CENP-32 was depleted in our U2OS stable cell lines using siRNA oligos targeting the endogenous mRNA and replaced with SPOUT1/CENP-32-GFP WT or SPOUT1/CENP-32-GFP variants (SPOUT1/CENP-32-GFP N86D, SPOUT1/CENP-32-GFP G98S, SPOUT1/CENP-32-GFP G244S, SPOUT1/CENP-32-GFP T289M, SPOUT1/CENP-32-GFP G293S, SPOUT1/CENP-32-GFP T353M) (Figure S13E-G). Consistent with the reduced activity observed for the patient variants in the *in vitro* MTase assays and zebrafish rescue experiments, induction of expression of the SPOUT1/CENP32-GFP variants failed to rescue both the detached centrosome phenotype and the appearance of micronuclei (Figure 8 and S14C). The T353M variant, which affects one of the key amino acid residues coordinating cofactor binding, showed the strongest phenotype of all the variants in our siRNA rescue assays with striking spindle abnormalities and centrosome splitting when it was expressed in endogenous SPOUT1/CENP-32 depleted cells (Figure 8). Expression of T353M led to an increased number of cells with detached centrosomes and an increased number of micronuclei even in the siRNA control condition (Figure 8). Similarly, when the effect of the SPOUT1/CENP-32-GFP variants on cell proliferation was assessed using the MTT colorimetric assay we observed that all variants failed to rescue the reduction on cell viability induced by SPOUT1/CENP-32 depletion, and the T353M was the variant that showed the strongest effect (Figure 8D).

**Figure 8:**
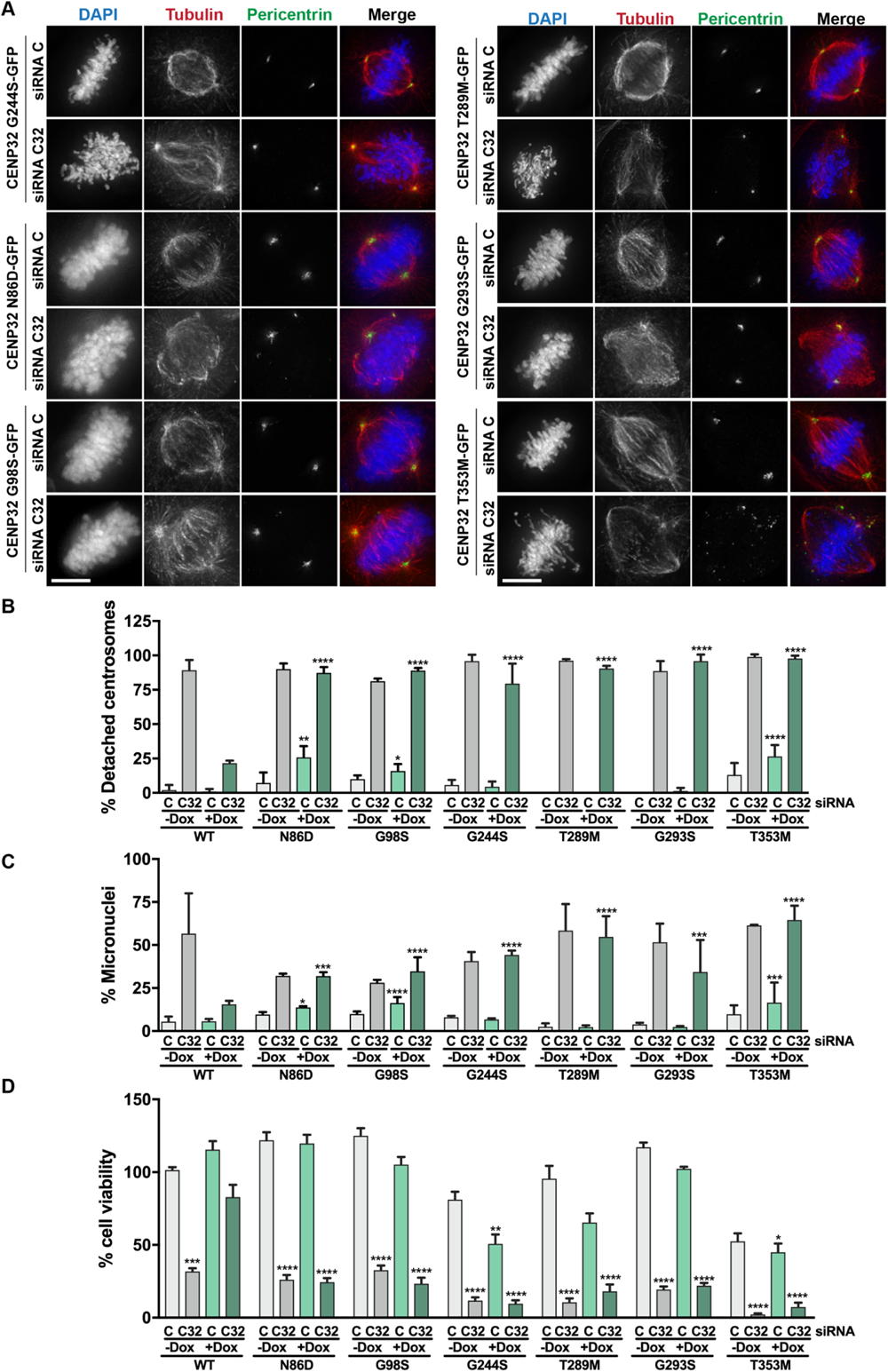
SPOUT1/CENP-32 patient variants lead to alterations of spindle organisation. (A) Representative fluorescence images of a SPOUT1/CENP-32 (C32) siRNA rescue assay in stable U2OS cell lines with inducible expression of different SPOUT1/CENP-32-GFP constructs. Conditions with doxycycline only; conditions without doxycycline in Figure S11C. (B, C) Quantification for the analysis of the centrosome detachment phenotype (% of cells with detached centrosomes; top panel) and chromosome segregation errors (% of micronuclei; bottom panel) in inducible U2OS cell lines expressing SPOUT1/CENP-32 WT-GFP or the SPOUT1/CENP-32 patient variants. Three independent experiments, *n* ≥ 70 cells analyzed in total per treatment, mean ± SD. Scale bar, 10 μm; Fisher’s exact test. For easy direct comparison with the SPOUT1/CENP-32 variant conditions, the SPOUT1/CENP-32 WT conditions shown in Figure 6D are also shown in this figure. Data are representative of a minimum of three biological replicates, mean ± SD. Scale bar, 10 μm. Fisher’s exact test with Bonferroni correction (*, P ≤ 0.05; **, P ≤ 0.01; ***, P ≤ 0.001; ****, P ≤ 0.0001). (D) Cell viability evaluation of inducible U2OS cell lines expressing SPOUT1/CENP-32 WT-GFP or SPOUT1/CENP-32 patient variants assessed by the MTT assay. For easy direct comparison with the SPOUT1/CENP-32 variant conditions, the SPOUT1/CENP-32 WT conditions shown in Figure 6E are also shown in this figure. Three independent experiments, mean ± SEM. Non-parametric ANOVA with Kruskal-Wallis followed by Dunn’s multiple comparisons test (*, P ≤ 0.05; **, P ≤ 0.01; ***, P ≤ 0.001; ****, P ≤ 0.0001).

Overall, our characterization of SPOUT1/CENP-32 patient variants in *in vitro* MTase assays and in functional rescue assays in a human cell line demonstrates that these patient-derived variants directly affect SPOUT1/CENP-32 MTase activity and compromise its role in maintaining the integrity and function of the mitotic spindle.

## DISCUSSION

Collectively, the spectrum of disorders caused by disruption of centrosome/spindle related proteins have been referred to as “centrosome-based diseases” [20] and the neurodevelopmental disorder associated with *SPOUT1/CENP-32* variants described in this study expands this group in a novel way. Most centrosome-based diseases are caused by pathogenic null variants in centrosomal genes such as *ASPM*, *WDR62*, *PCNT*, and *CDK5RAP2* ^[46-48]^. *SPOUT1/CENP-32* differs, because most of the variants reported in this study are missense or in-frame variants with decreased catalytic activity. Our newly generated zebrafish *spout1/cenp-32* mutants are lethal by 8 dpf; and we were unable to generate a homozygous *spout1/cenp-32* null strain in *C. elegans* (Figure S15). Furthermore, mice homozygous for knockout of the overlapping *Spout1/Cenp-32* and *EndoG* loci exhibit embryonic lethality before implantation (MGI: 106544) ^[34]^. Therefore, all evidence indicates that *SPOUT1/CENP-32* is an essential gene for organismal viability^[16]^.

Our structure/function analysis reveals that SPOUT1/CENP-32 possesses a SPOUT methyltransferase domain. *In vitro* assays confirm that SPOUT1/CENP-32 is indeed an active RNA-methyltransferase. Importantly, this RNA methyltransferase activity of SPOUT1/CENP-32 is crucial for mitotic spindle assembly and accurate chromosome segregation. Strikingly, most *SPOUT1/CENP-32* bi-allelic variants found in patients are located close to the catalytic site, perturbing the methyltransferase activity of SPOUT1/CENP-32 *in vitro* and result in the detachment of centrosomes from mitotic spindle poles in a human cell line. Of note, the recurrent variants N86D, G98S and T353M showed the most significant defects in the functional studies, which correlate with *in silico* prediction scores (Table S2). The G98S variant significantly reduced methyltransferase activity *in vitro* and failed to rescue the morpholino-induced head size phenotype in zebrafish, and caused chromosome segregation defects in cultured cells. This variant was found in 8/18 families in the cohort indicating that it is likely a founder variant in the Middle-eastern Arab population. Given that the G98S variant is located at the dimeric interface of the protein, it further highlights and supports the importance of dimerization for the enzymatic activity of the protein.

The most striking cellular phenotypes were observed for the T353M variant, seen in 2/18 families in the cohort, which showed an increased number of cells with detached centrosomes and an increased number of micronuclei. In addition to these cellular phenotypes, the reduction in head size phenotype in zebrafish was only partially rescued by the T353M variant, suggesting that it is a hypomorph, with some residual activity. Thus, both assays were consistent with the pathogenicity of the T353M variant.

Tethering of chromosomes to spindle poles is crucial to maintain the bipolar architecture of the mitotic spindle required for accurate chromosome segregation. Several molecular players, including dynein-dynactin, NuMA, CDK5RAP2/centrosomin, WDR62 and HSET have been implicated in centrosome-spindle pole attachment ^[49-58]^. Depletion of WDR62 or CDK5RAP2 disrupts centrosome attachment to spindle poles but does not affect centrosomal nucleation of microtubules ^[49, 50]^. These observations suggest that centrosomal microtubules alone are insufficient to connect centrosomes to spindle poles, and the centrosome-spindle pole connection is likely maintained through physical tethering. Interestingly, SPOUT1/CENP-32 depletion did not abolish the centrosome association of CDK5RAP2, a component suggested to physically tether centrosome to spindle poles, but changed the abundance of CG-NAP, a PCM component, which failed to accumulate on the dissociated centrosomes in SPOUT1/CENP-32 depleted cells ^[15]^. However, CG-NAP depletion alone did not phenocopy SPOUT1/CENP-32 depletion, suggesting that SPOUT1/CENP-32 regulates multiple centrosome components essential for centrosome-spindle pole attachment through mechanisms that are yet to be determined.

The precise mechanism by which SPOUT1/CENP-32 RNA methyltransferase activity ensures centrosome-spindle pole tethering remains to be elucidated. We hypothesise that SPOUT1/CENP-32 catalytic activity modulates the abundance and interactions of centrosomal proteins or RNAs essential for tethering centrosomes to spindle poles. RNA methyltransferases are known to regulate RNA stability, localisation and translation efficiency ^[59]^. Most of the well characterised SPOUT MTases are involved in 2’-O-ribose methylation, which is mostly found in ribosomal and small nuclear RNAs, that mainly occurs in functionally important regions of the ribosome and spliceosome ^[40]^. Moreover, RNA and RNA-binding proteins have also been implicated in centrosome function, as well as in the formation of the mitotic spindle ^[60-69]^. Some examples include NSUN2, a nucleolar RNA MTase required for spindle formation ^[62]^ and RAE1, an RNA-binding protein that regulates spindle assembly in an RNA-dependent manner ^[68]^. In support of our hypothesis, the SPOUT1/CENP-32 homologue in fission yeast was shown to interact with a newly identified centrosomal protein, Wdr8, which is involved in anchoring microtubules to the centrosome ^[70, 71]^ and with dynactin 4 (DCNT4), a subunit of the dynactin complex ^[72]^. Future studies will aim to elucidate the mechanistic links between SPOUT1/CENP-32 methyltransferase activity and centrosome-spindle pole tethering.

Besides the centrosome detachment phenotype, SPOUT1/CENP-32 depleted cells also showed chromosome segregation errors, as indicated by the formation of micronuclei. Accurate chromosome segregation is essential for maintaining genomic stability during mitosis, and aneuploidy resulting from chromosome mis-segregation is a cancer hallmark considered to be a driving force for tumorigenesis and tumor evolution ^[73, 74]^. However, we karyotyped >50 metaphase cells in peripheral blood cultures from individual A-III-2, but no aneuploidy was detected (Figure S16). It is possible that those aneuploid cells are eliminated by apoptosis *in vivo* in humans. Further studies will be required to determine whether individuals with *SPOUT1/CENP-32*-associated neurodevelopmental disorder have an increased risk of developing aneuploidy *in vivo* or even cancers.

In conclusion, we report here that bi-allelic variants of the *SPOUT1/CENP-32* gene in humans are associated with a novel autosomal-recessive complex neurodevelopmental disorder: SpADMiSS. Through extensive experimental studies from biochemical and cellular functions to animal models, we demonstrate that the variants identified in affected individuals reduce the RNA methyltransferase activity of the SPOUT1/CENP-32 protein, causing centrosome detachment from spindle pole and chromosome mis-segregation. We propose that these defects delay the cell cycle and trigger apoptosis in neural progenitor cells, leading to neurodevelopmental defects in humans (Figure 9).

**Figure 9:**
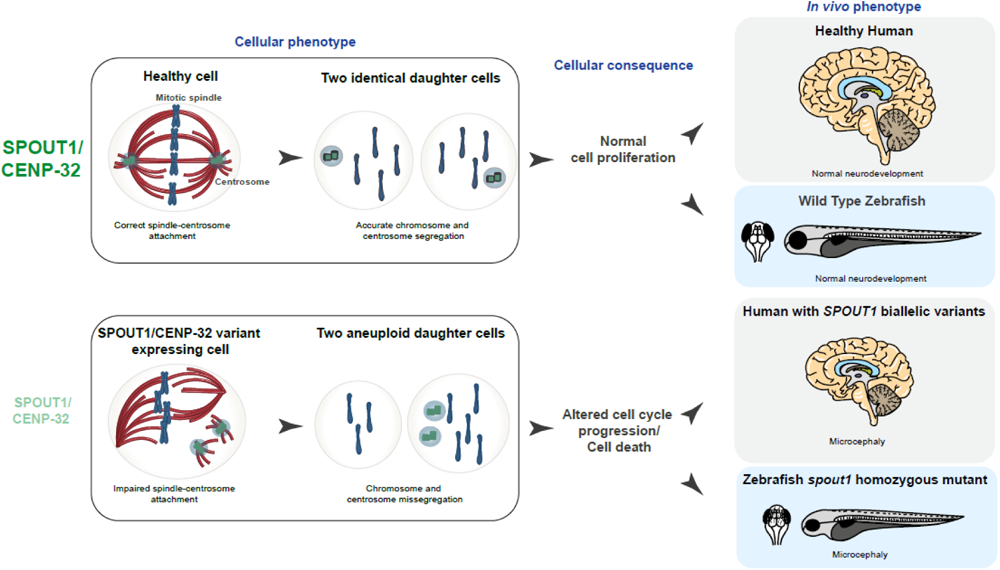
*SPOUT1/CENP-32* bi-allelic variants disrupt centrosome-spindle pole tethering, causing chromosome segregation errors, altering cell cycle progress, and increasing apoptosis in neuronal progenitor cells. Schematic summarizing the proposed differences between healthy cells and *SPOUT1/CENP-32* variant cells leading to aneuploid daughter cells, altered cell cycle/ cell death and microcephaly in affected individuals and mutant zebrafish.

## MATERIALS AND METHODS

### Ethics statement, recruitment of families and evaluation

Informed consent was obtained from all families. Clinical evaluation and genetic analyses were approved by respective institutional ethics committees from the following participating centers: Columbia University Medical Center (IRB# AAAR1159), Children’s Hospital of Philadelphia (IRB #16-013231), Baylor College of Medicine (IRB# H-29697), and Nationwide Children’s Hospital (IRB# 11-00215). All participating clinical centers were connected through GeneMatcher ^[17]^. Each participating patient in this study was given a subject ID, which cannot be associated with their identity by anyone outside the research group. Peripheral blood was obtained from participants and genomic DNA was extracted as per standard protocols. A pediatric epileptologist (T.T.S.) reviewed the genetic test results and clinical reports, and evaluated the magnetic resonance imaging (MRI) and electroencephalographic (EEG) recordings, where available.

### Genetic analyses

Exome sequencing (ES)/ Genome sequencing (GS) and variant calling were performed on DNA extracted from peripheral blood mononuclear cells using commercial exome libraries and Illumina, and standard clinical and research bioinformatic pipelines (see Supplementary case reports). Single nucleotide variants (SNVs) and indels were classified in accordance with the ACMG/AMP sequence interpretation guidelines ^[75]^. Sanger dideoxy DNA sequencing was used to verify candidate variants identified by ES/GS, determine inheritance and segregation within families. Additional genetic studies included karyotype analysis, chromosomal microarray, homozygosity mapping, and mitochondrial testing following standard protocols.

### Zebrafish husbandry and embryo maintenance

We performed all zebrafish experiments according to protocols approved by the Northwestern University Institutional Animal Care and Use Committee (IACUC). Adult zebrafish were housed in standard husbandry conditions in a 14h light-10h dark cycle. We obtained fertilized embryos by natural mating of wild type (WT) NIH or *-1.4col1a1:egfp* transgenic reporter adults. The embryos were maintained in fresh E3 medium (0.581 g/L NaCl, 0.026 g/L KCL, 0.096 g/L CaCl_2._2H_2_O, 0.163 g/L MgCl_2._2H_2_O, and 0.0002% methylene blue) at 28°C until phenotype endpoint. Larvae were evaluated for apoptosis and cell cycle progression at 2 dpf and head size and brain structures at 3 dpf.

### CRISPR/Cas9 mediated genome editing in zebrafish embryos

We performed reciprocal BLAST of human SPOUT1/CENP-32 (NP_057474.2) against the translated zebrafish genome and identified a single *spout1/cenp-32* zebrafish ortholog (Ensembl ID: GRCz11: ENSRDARG00000019707) with 76% identity and 88% similarity. To identify single guide (sg) RNA sites targeting *spout1/cenp-32*, we used CHOPCHOP (https://chopchop.cbu.uib.no/) ^[76-78]^ (Table S3). Using synthetic oligos as template, we synthesized sgRNAs targeting exon 5 and exon 7 of ENSDART00000099535.5 using the GeneArt Precision gRNA Synthesis Kit (ThermoFisher Scientific), according to the manufacturer’s guidelines. We injected 1 nL cocktail containing 100 pg sgRNA with or without 200 pg Cas9 protein (PNA Bio, CP01) into the cell of single-cell stage embryos. To determine sgRNA targeting efficiency, we extracted genomic DNA from individual F0 embryos at 2 dpf by proteinase K digestion (Life Technologies, AM2548). The region flanking the sgRNA target was PCR-amplified, denatured, slowly reannealed, and then migrated on a 20% polyacrylamide gel (ThermoFisher Scientific) to separate heteroduplexes ^[79]^ (n = 2 uninjected controls and n = 6 F0 mutants tested). To determine F0 mosaicism, we cloned the PCR-amplified product into TOPO-TA cloning vector (ThermoFisher Scientific), and individual colonies were sequenced (n = 16 colonies/embryo) with BigDye Terminator 3.1 chemistry (Applied Biosystems). To generate *spout1/cenp-32* mutants, we outcrossed F0 mosaic mutants with WT (NIH) adults and isolated F1 heterozygous animals carrying a 1 bp insertion (Ensembl ID: ENSDART00000099535.5 c.543_544insA; p.R215Kfs*7) in exon 7.

### DNA extraction from Dent’s or paraformaldehyde fixed embryos

If required, fixed larvae were genotyped after wholemount imaging as described ^[80, 81]^. Briefly, larvae were placed in PCR tubes and incubated in 40 µl of lysis buffer (25 mM NaOH, 0.2 mM EDTA) for 45 min at 95°C. The tubes were cooled to 4°C before adding an equal volume of HotSHOT neutralization buffer (40 mM Tris-HCl); 5 µl DNA lysate was used for PCR.

### Quantitative real-time PCR (qPCR)

For qPCR analysis, we incrossed F1 heterozygous mutants and then decapitated larvae at 3 dpf. Tails were used for DNA extraction and genotyping; matched pooled heads (n=18) were used for total RNA extraction using Trizol (ThermoFisher Scientific). We reverse transcribed 1 µg RNA to cDNA using QuantiTect reverse transcription kit (Qiagen) according to the manufacturer’s protocol. qPCR was performed on a QuantStudio 3 real time PCR instrument (Applied Biosystems), using SYBR green chemistry (Applied Biosystems) according to manufacturer’s instructions. The primer pairs used for qPCR annealed to exons 3-4 (upstream) and exon 11-12 (downstream) of the mutation site (listed in Table S3). The thermocycler conditions were: 95 °C for 10 mins; 39 cycles (95°C for 15 sec and 60°C for 1 min); 95°C for 15 sec; 60°C for 15 sec; 95°C for 15 sec. The relative gene expression was calculated by 2^-ΔΔCt^ and normalized to *b-actin*. Two biological and 6 technical replicates were performed for each sample.

### Transient suppression of *spout1/cenp-32* by Splice Blocking Morpholino (sb-MO)

We designed a morpholino (MO) antisense oligonucleotide targeting the *spout1/cenp-32* exon 4-intron 4 junction (e4i4: 5’-GTCAAAATAGCAGCTCACTTTGCGT-3’) to block the exon 4 splice donor site (Gene Tools, LLC). We obtained fertilized embryos from natural matings of adult fish and injected 1 nL MO into the yolk at the one-to-four cell stage with increasing doses of e4i4 (3 ng, 6 ng, and 9 ng). To assess MO efficiency, embryos were harvested for total RNA extraction using Trizol reagent (ThermoFisher Scientific) at 2 dpf (20 embryos/condition, pooled) according to manufacturer’s instructions. cDNA was generated with QuantiTect Reverse Transcription Kit (Qiagen) according to the manufacturer’s protocol, followed by PCR with primers flanking the MO target locus (Table S3), and migration of the amplified product on a 1% agarose gel. Resulting PCR bands were gel purified (QIAquick^®^ Gel Extraction Kit, Qiagen) and cloned into TOPO-TA cloning vector (ThermoFisher Scientific) and resulting colonies were Sanger sequenced the (n = 4/PCR band).

### Molecular cloning of human ORF *(SPOUT1/CENP-32)*, site directed mutagenesis and *in-vitro* transcription

We acquired a Gateway-compatible human *SPOUT1/CENP-32* full length open reading frame (ORF) entry clone (GenBank: BC033677.1 ThermoFisher Scientific; IOH40219) and cloned it into a Gateway-compatible pCS2^+^ destination vector via LR clonase-mediated recombination (ThermoFisher Scientific). Next, we used it as a template to generate mutant constructs harboring *SPOUT1/CENP-32* variants identified in affected individuals (p.G98S, p.T353M, p.N86D, p.G293S, p.R200W, p.R352C, p.G244S, p.L314F, p.Y339C, p.T289M and p.S249del) or in gnomAD (dbSNP ID: rs6478854, p.T130R; negative control) by site-directed mutagenesis as described ^[36]^ (Table S3). The integrity of all constructs was confirmed by Sanger sequencing. To generate capped mRNA for rescue experiments, we linearized pCS2^+^ constructs with NotI and used the resulting template for *in vitro* transcription with the mMessage mMachine SP6 Transcription kit (ThermoFisher Scientific) according to manufacturer’s guidelines. We used 100 pg *SPOUT1/CENP-32* mRNA with 6 ng MO for *in vivo* complementation studies.

### Zebrafish phenotyping

#### Live imaging of zebrafish larvae and bright field morphometrics

To assess the head size and body length of *spout1/cenp-32* models, we anesthetized larvae with 0.2 mg/mL Tricaine (MS-222) at 3 dpf prior to loading into a 50 ml conical tube attached to the Vertebrate Automated Screening Technology (VAST) Bioimager (Union Biometrica). We passed larvae sequentially through a 600 μm capillary in the stage-mounted detection platform on an AxioImager.M2m microscope (Zeiss). We created dorsal and lateral image templates, and imaged them live with VAST software (version 1.2.6.7) with >60% minimum similarity for the pattern-recognition algorithm. Bright field (Dorsal and lateral) images were acquired with the on-board camera and VAST software using default parameters as described ^[21, 82, 83]^. Head size area and body length were measured on lateral bright field images with ImageJ (NIH) software (version 1.53a). Specifically, head size was arbitrarily defined with three consistent landmarks and connected with a continuous line from the posterior otolith (line 1) to the nearest dorsal point from the otolith (line 2) to the most anterior point of the head at the mouth (line 3), and returning to the posterior otolith. Body length was measured by drawing a straight line from the most anterior part to the most posterior part.

### Whole-mount TUNEL assay

Terminal deoxynucleotidyl transferase biotin-dUTP nick labeling (TUNEL, ApopTag Red In Situ Apoptosis Detection Kit, Millipore Sigma) was used to detect apoptotic cells in whole mount zebrafish larvae ^[23, 28, 84]^. Briefly, embryos were dechorionated at 2 dpf and fixed in 4% paraformaldehyde (PFA) overnight at 4°C, then dehydrated in 100% methanol at −20 °C overnight. The larvae were then gradually rehydrated in methanol in PBS and 0.1% Tween (PBST) in the following percent volume/volume ratios: 75/25; 50/50; and 25/75 for 10 min each at room temperature (RT). After rehydration in methanol/PBS, the larvae were bleached for 10 min in bleaching solution (0.5% KOH and 3% H_2_O_2_ in PBST) before permeabilization with proteinase K (10 µg/ mL) and post-fixation with 4% PFA for 20 min at RT. The larvae were then treated with equilibration buffer (Millipore Sigma) for 1h at room temperature followed by overnight incubation with TdT enzyme in a humidified incubator. The next day larvae were washed with PBST, and then treated with the anti-digoxigenin-rhodamine (Millipore Sigma) for 2h at RT and washed three times for 10 min each with PBST and processed for imaging.

### Immunostaining on whole-mount zebrafish larvae

Whole mount anti-phospho-histone H3 and acetylated tubulin immunostaining were performed as described ^[22, 24, 85]^. In brief, embryos were dechorionated at 2 dpf (pHH3), or larvae at 3 dpf (acetylated tubulin), and were fixed overnight in 4% PFA or Dent’s solution (80% MeOH + 20% DMSO), respectively. Larvae were then rehydrated in PBS with stepwise reduced concentration of methanol at RT as mentioned above, followed by bleaching for 10-15 mins described elsewhere ^[21, 28]^. The larvae were permeabilized with proteinase K (10 µg/ mL) before post fixation with 4% PFA for 20 mins at RT, and washed twice in IF buffer (1% BSA, 0.1% Tween-20 in 1xPBS) for 10 mins. The larvae were then incubated in blocking solution (IF buffer and 10% FBS) for 1 h, followed by overnight incubation in primary antibody (anti-pHH3, 1:500, Santa Cruz Biotechnology, sc-374669; anti-α-acetylated tubulin, 1:1,000 Sigma Aldrich, T7451) at 4 °C. After two washes in IF buffer, the larvae were then treated with secondary antibody (Alexa Flour 488 goat anti-rabbit IgG, 1:500; Alexa Flour 594 goat anti-mouse IgG, 1:500) in blocking solution for 2 hrs at RT. The embryos were then washed three times with IF buffer, and processed for imaging.

### Fluorescence microscopy and image processing

We captured Z-stacked images of the fluorescent signal with a Nikon AZ100 microscope equipped with Nikon camera and Nikon NIS Elements software (TUNEL, pHH3, and α-acetylated tubulin). We counted apoptotic and proliferative cells in a consistently defined head region, using the image-based tool for counting nuclei (ICTN) plugin with the following parameters: width: 12, min distance: 10, and threshold: 1 in ImageJ (version 1.53a) ^[28]^. We also measured the area of the optic tecta, and number of intertectal neurons crossing the midline between optic tecta as described ^[22, 24]^.

### Statistical analysis for zebrafish studies

Each experiment was performed at least twice, and measurements were taken with the investigator blinded to experimental condition (see Table S4 for experimental data). All zebrafish morphometric data were analyzed using Prism 9 software (GraphPad, San Diego, California, USA). Data were first evaluated using descriptive statistics to check equality of variances and normal distributions (e.g., D’Agostino-Pearson omnibus test). When necessary, we used a non-parametric ANOVA (Kruskal-Wallis test) followed by Dunn’s multiple comparisons test. Correction for multiple comparisons was accomplished using the original method of Benjamini and Hochberg (Benjamini and Hochberg, 1995) to control the False Discovery Rate (FDR) which was set at 5% (Q = 0.05).

### Mouse care and experimentation

All animal experiments were approved by the Bioethics Committee of Lobachevsky State University of Nizhny Novgorod (protocol No. 14 dated 01.19.2018). C3H mice were used for in utero electroporation experiments. C57BL/6 mice at e14.5, e16.5, e18.5 were used to prepare slices for in situ hybridization. The day on which the plug was detected was considered embryonic day 0.5 (E0.5).

### In utero electroporation

In utero electroporation was performed as previously described (https://doi.org/10.1093/cercor/bhab172) using a FemtoJet 4i microinjector (Eppendorf, Germany) and a NEPA21 electroporator (NEPA GENE, Japan). Following surgery, animals were kept under observation until full recovery and the embryos collected at the pregnancy stages indicated in the experiments. The plasmids used in this study are presented in section cloning of target vectors (Supplementary methods).

### In situ hybridization

Sectioning, in situ hybridization, washing were performed as described (https://doi.org/10.1093/nar/gkad703). Three independent in situ analyses were performed for each stage. The primers used to generate the RNA probes had the sequences as listed: Spout1 fw-GACCTCGAGAAGGGTTGAATGGCGAAAATGG, Spout1 rw-AATTAACCCTCACTAAAGGGCGGCCGCCACAGTTGACCAAAGAGCCATG

### Immunohistochemistry

Immunohistochemistry and tissue processing were performed as previously described (https://doi.org/10.1016/j.ydbio.2006.06.040) The primary antibodies used were a-BrDU (mouse), a-GFP (goat), a-Ki67(rabbit).

### Image acquisition and processing

The slides resulting from in situ hybridization were imaged on a Zeiss Axio Vert.A1. For the immunofluorescently stained tissue preparations, we used a LSM 800 (Carl Zeiss) confocal laser scanning system. For each electroporated litter, brain sections were matched for anteroposterior and lateromedial position of the electroporation site. Counting was performed blinded.

### Statistical analysis for mouse embryo studies

Statistical analysis for laboratory experiments was performed using the Prism software (GraphPad), in accordance with the nature of the experimental setup and employing the tests indicated in the experiments.

### Protein expression and purification of SPOUT1/CENP-32 for crystallisation

The methyltransferase domain of SPOUT1/CENP-32 (SPOUT1/CENP-32 71-376) was cloned into a pEC-K-3C-His vector as an N-terminally His-tagged protein with a 3C site. The catalytic site variant (A356N) was obtained using the Quikchange site-directed mutagenesis procedure (Agilent). Both selenomethionine-labelled and native SPOUT1/CENP-32 71-376 were expressed in BL21 gold *E.Coli* strain by overnight induction at 20 °C or 18 °C with 0.35 mM IPTG, respectively. Cells were lysed by sonication on lysis buffer containing 20 mM Tris pH 8, 500 mM NaCl, 35 mM Imidazole and 2 mM beta-mercaptoethanol. The pre-cleared lysate was loaded onto a 5 ml HisTrap^TM^ HP column (Cytiva), washed with 50 cv of lysis buffer, 20 cv of high salt buffer (20 mM Tris pH 8, 1 M NaCl, 35 mM Imidazole, 50 mM KCl, 10 mM MgCl_2_, 2 mM ATP and 2 mM beta-mercaptoethanol) and eluted with elution buffer (20 mM Tris pH 8, 200 mM NaCl, 400 mM Imidazole and 2 mM beta-mercaptoethanol). Proteins were then cleaved overnight at 4 °C with 3C protease while dialyzing against 20 mM Tris pH 8, 200 mM NaCl and 5 mM DTT. Proteins were further purified by size-exclusion chromatography in 20 mM Tris pH 8, 200 mM NaCl and 5 mM DTT (Superdex 75 10/300 GL, Cytiva).

### SPOUT1/CENP-32 crystallisation, data collection, crystal structure solution and refinement

Gel filtrated selenomethionine SPOUT1/CENP-32 71-376 and native SPOUT1/CENP-32 71-376 A356N were concentrated using Vivaspin^TM^ Turbo 10 kDa MWCO concentrators to 16 – 20 mg/ml. Crystallisation trials were performed with a nanoliter Crystal Gryphon robot (Art Robins) and grown by vapour diffusion using the Morpheus^TM^ screen (Molecular Dimensions). SPOUT1/CENP-32 71-376 proteins crystallised in several conditions of the Morpheus screen. The best resolution for the selenomethionine-labelled SPOUT1/CENP-32 71-376 was obtained from the crystals grown in conditions containing: 0.06 M MgCl_2_, CaCl_2_ or 0.09 M NaF, NaBr, NaI with 0.1 M Imidazole, MES (acid) pH 6.5 and 30 % Ethylene glycol, PEG 8K. To obtain the SPOUT1/CENP-32 71-376 – SAM structure, selenomethionine-labelled SPOUT1/CENP-32 71-376 crystals were soaked with 10 time’s molar excess of SAM (ab142221, Abcam). The crystals that diffracted better for native SPOUT1/CENP-32 71-376 A356N protein grew in 0.09 M NaF, NaBr, NaI; 0.1 M Sodium HEPES, MOPS (acid) pH 7.5 and 30 % Ethylene glycol, PEG 8K.

Diffraction data were collected on beamlines i03 (SPOUT1/CENP-32 71-376 SAH and SPOUT1/CENP-32 71-376 SAM) and i04-1 (SPOUT1/CENP-32 71-376 A356N) at the Diamond Light Source (Didcot, UK). All datasets were processed using the software pipeline available at Diamond Light Source that utilises XDS, CCP4, CCTBX, AutoPROC and STARANISO ^[86-91]^. SPOUT1/CENP-32 71-376 SAH structure was determined using Single Anomalous Dispersion method using Se-Met phasing. SPOUT1/CENP-32 71-376 SAM and SPOUT1/CENP-32 71-376 A356N structures were determined by molecular replacement with the program PHASER ^[92]^ using the coordinates of SPOUT1/CENP-32 71-376 SAH structure. Structures were refined using the PHENIX and CCP4 suite of programs ^[93, 94]^. Model building was done using COOT ^[95]^ and figures were prepared using PyMOL (http://www.pymol.org). Data collection, phasing and refinement statistics are shown in Table S5.

The structural coordinates and structure factors reported in this paper have been deposited in the PDB (http://www.rcsb.org/) with the following accession numbers: 8QSU for SAH-bound CENP-32 71-376, 8QSV for SAM-bound CENP-32 71-376 and 8QSW for CENP-32 71-376 catalytic mutant (A356N).

### Methyltransferase assays

NSUN6 was cloned in a pET-His6-MBP vector (pET His6 MBP TEV cloning vector with BioBrick polycistronic restriction sites (9C) was a gift from Scott Gradia; Addgene plasmid #48286; http://n2t.net/addgene:48286 ; RRID:Addgene_48286) and purified as previously described ^[44]^ with a few variations. Briefly, His_6_-MBP-NSUN6 was expressed in *E. coli* BL21 (DE3) pLySs cells by induction with 0.5 mM IPTG at 18 °C for 16 hrs. Cells were lysed by sonication in a lysis buffer containing 30 mM KPi pH 7, 300 mM KCl, 10 % Glycerol, 25 mM Imidazole and 1 mM beta-mercaptoethanol. The lysate was then cleared by centrifugation at 22,500 rpm for 50 min 4 °C and loaded onto a 5 ml HisTrap^TM^ HP column (Cytiva). The protein-bound column was washed with 60 cv of lysis buffer and 40 cv of high salt buffer (30 mM KPi pH 7, 500 mM KCl, 10 % Glycerol, 35 mM Imidazole and 1 mM beta-mercaptoethanol). Protein was eluted with elution buffer containing 30 mM KPi pH 7, 300 mM KCl, 10 % Glycerol, 200 mM Imidazole and 1 mM beta-mercaptoethanol and the pool of elutions was dialysed overnight at 4 °C into storage buffer (30 mM KPi pH 7, 100 mM KCl, 50 % Glycerol, 1 mM DTT and 0.1 mM EDTA).

Survivin was used as a negative control in our methyltransferase assays as it does not have methyltransferase activity. Full length Survivin was cloned into a pEC-K-3C-His vector as an N-terminally His-tagged protein with a 3C cleavage site. The vector was transformed in *E. coli* BL21 Gold strain and grown in Super broth media at 37°C until O.D 1-1.5. Cultures were induced over night at 18 °C with 0.35 mM IPTG. Cells were lysed in a buffer containing 20 mM Tris–HCl pH 8, 150 mM NaCl, 25 mM imidazole and 2 mM 2-mercaptoethanol. The protein was purified by affinity chromatography using a 5 ml HisTrap^TM^ HP column (Cytiva). The protein-bound column was washed with 40 column volumes of lysis buffer followed by 20 column volumes of high salt buffer (20 mM Tris–HCl pH 8, 1 M NaCl, 25 mM imidazole and 2 mM 2-mercaptoethanol). The protein was eluted and dialysed overnight at 4 °C in 20 mM Tris–HCl pH 8, 150 mM NaCl, 2 mM DTT while cleaving with 3C protease. The cleaved protein was concentrated with a Vivaspin^TM^ 10 kDa MWCO concentrator (Millipore), and the concentrated protein was loaded onto a Superdex 200 increase 10/300 GL column (Cytiva) equilibrated with 20 mM Hepes pH 7.5, 100 mM NaCl and 2 mM DTT.

Full length SPOUT1/CENP-32 WT, A356N, T130R, and patient variants (N86D, G98S, G293S, G244S, T289M, T353M) were cloned as described above in a pEC-K-His vector as N-terminally His-tagged proteins and expressed using the *E.Coli* BL21 Gold strain in 250 ml of Super Broth media. Harvested cells were lysed by sonication in lysis buffer containing 20 mM Tris pH 8, 500 mM NaCl, 35 mM Imidazole and 2 mM beta-mercaptoethanol, supplemented with RNase, DNase and PMSF. Lysates were incubated with 2 ml of HisPur^TM^ Ni-NTA resin (Thermo Fisher Scientific) for 2 h at 4 °C and protein-bound beads were washed with 30 cv of lysis buffer followed by 20 cv of high salt buffer (20 mM Tris pH 8, 1 M NaCl, 35 mM Imidazole and 2 mM beta-mercaptoethanol). Proteins were eluted with 20 mM Tris pH 8, 300 mM NaCl, 400 mM Imidazole and 2 mM beta-mercaptoethanol and pools of elutions were dialysed over night against 20 mM Hepes pH 8, 300 mM NaCl, 1 mM DTT and 20 % Glycerol. Methyltransferase assays were performed straight after dialysis.

The total RNA used as a substrate in the methyltransferase assays was isolated from asynchronous SPOUT1/CENP-32 depleted (using predesigned siRNA oligos directed against human SPOUT1/CENP-32 (target sequence: TCGCAGGACCCTCGCACCAAA; Qiagen) or NSUN6 depleted (target sequence: TTCCTGTTATTGGACCCAGAA)^[44]^, U2OS cultures via the TRIzol^TM^ (Thermo Fisher Scientific) method that involves homogenisation of the cell lysate, phase separation, RNA precipitation and RNA washes. The RNA hairpin of GAPDH mRNA used as a substrate in the methyltransferase assays was synthesised by Integrated DNA Technologies: GCCCCCUCUGCUGAUGCCCCCAU GUUCGUCAUGGGUGUGAA.

Methyltransferase assays were performed using the MTase-Glo^TM^ Methyltransferase assay (Promega Corporation) following manufacturer’s instructions. Briefly, assays were performed at room temperature with reaction buffer containing 20 mM Tris pH 8, 50 mM NaCl, 1 mM EDTA, 3 mM MgCl_2_, 0.1 mg/ml BSA and 1 mM DTT in flat bottom non-binding 96 well microplates (Greiner BioOne). The proteins were incubated with the substrates (either total RNA or *GAPDH* mRNA oligo) for 30 min, when 0.5 % TFA was used to stop the reaction. 5 μL of 6× MTase-Glo Reagent were then added to each well and incubated for 30 min at room temperature. For the generation of the bioluminescent signal, 30 μL of MTase-Glo detection reagent were added to the reaction and incubated for a further 30 min at room temperature. Luminescence was measured using SpectraMax M5 Multi-mode microplate reader (Molecular Devices). Experiments were performed at least in triplicate, and values expressed as means ± the standard error of the mean (SEM). Data were analyzed using nonlinear regression to determine Michaelis-Menten kinetics in GraphPad Prism version 7.0.

### DNA constructs and generation of SPOUT1/CENP-32-GFP stable cell lines

For CRISPR-mediated double-strand break site formation, 54-bp DNA fragments were generated by annealing two complementary single-stranded oligonucleotides. The fragments were inserted into the AgeI (#R3552, New England BioLabs [NEB]) site of pTORA14 ^[96]^ using the In-Fusion HD Cloning Kit, yielding pTORA14AAVS1 which were used to induce DNA cleavage at adeno-associated virus integration site 1 (AAVS1). The DNA sequences targeted by the guide RNAs are listed in Table S6.

pDEST243NGFP was generated by inserting a DNA fragment encoding EGFP amplified (primers are listed in Table S7) from pDEST131NGFP ^[96]^ by PCR, between the BglII (#R0144, NEB) and MluI (#R0198, NEB) sites of pMK243 ^[97]^ with the In-Fusion HD Cloning Kit (#Z9648N, Takara Bio). The DNA fragment encoding human SPOUT1/CENP-32^71-end^ were PCR-amplified (primers are listed in Table S7) from pEGFP-N1-CENP32_71-end. The fragments were inserted into pDEST243NGFP between the BglII (#R0144, NEB) and SalI (#R0138, NEB) sites using the In-Fusion HD Cloning Kit, yielding pDEST243CENP32^71-end^. The resultant plasmids were used to generate a human cell line expressing the SPOUT1/CENP32^71-end^:GFP fusion protein via CRISPR-mediated homologous recombination (HR) at AAVS1. The fragments generated from the digestion of pEGFP-N1-CENP-32 WT, pEGFP-N1-CENP-32 A356N, pEGFP-N1-CENP-32 G244S, pEGFP-N1-CENP-32 T289M, pEGFP-N1-CENP32 T353M or pEGFP-N1-CENP32 G293S by BglII and AgeI were inserted into pDEST243CENP32^71-376^ between the BglII and AgeI sites using the T4 DNA ligase (#M0202, NEB), yielding pDEST243CENP32^WT^, pDEST243CENP32^A356N^, pDEST243CENP32^G244S^, pDEST243CENP32^T289M^, pDEST243CENP32^T353M^, pDEST243CENP32^G293S^. The resultant plasmids were used to generate a human cell line expressing the CENP32^WT^, CENP32^A356N^, CENP32^G244S^, CENP32^T289M^, CENP32^T353M^ or CENP32^G293S^:EGFP fusion protein via CRISPR-mediated homologous recombination (HR) at AAVS1.

A U2OS-<u>c</u>onditional <u>o</u>ver<u>e</u>xpressing (cOX) SPOUT1/CENP-32 and its derivative*s* cell line, which is the parental cell line of *CENP-32*:cOX cells, was generated by transfecting U2OS cells with pTORA14AAVS1 and pDEST243CENP32 using Lipofectamine LTX Reagent (#A12621, Thermo Fisher Scientific) followed by puromycin selection (1 µg/mL, #10-2100, Focus Biomolecules). The genome editing was confirmed by PCR-amplification from the genomic DNA using Tks Gflex DNA Polymerase (#R060A, Takara Bio) and the primers listed in Table S7. Also the expression of the GFP fusion protein was confirmed by microscopic observation.

### *In vitro* rescue experiments and Immunofluorescence microscopy

The U2OS human osteosarcoma cells were routinely maintained in DMEM (Thermo Fisher Scientific) supplemented with 10% fetal bovine serum and penicillin/streptomycin (10,000 U/ml; Thermo Fisher Scientific).

Lipofectamine RNAimax was used for depletion of endogenous SPOUT1/CENP-32 using predesigned siRNA oligos directed against human *SPOUT1/CENP-32* (target sequence: TCGCAGGACCCTCGCACCAAA; Qiagen). Luciferase targeting was used as a control (5′-CGUACGCGGAAUACUUCGAdTdT-3′; All Star, Qiagen) ^[98]^. Cells were plated in glass coverslips in 12 well plates and 16 h after plating they were transfected with 15 pmols of siRNA oligos and incubated for 48 hrs. For analysis of the centrosome detachment phenotype, cells were fixed with ice-cold methanol for 5 min rehydrated with PHEM buffer (60 mM PIPES pH 6.9, 25 mM Hepes, 10 mM EGTA and 2 mM MgCl_2_) and blocked with 1 % BSA in PHEM buffer for 30 min at RT. Cells were stained with mouse anti-tubulin (B512; 1:1000; T5168; Sigma) and rabbit anti-pericentrin (1:1000; ab4448; Abcam). The secondary antibodies used were donkey anti-mouse Cy5 (1:300, 715-175-151; Jackson Laboratories) and goat anti-rabbit TRITC (1:300; 111-025-006; Jackson Laboratories). Slides were mounted with Vectashield with DAPI (H-1200, Vector Labs). A minimum of 70 cells per condition were quantified. The same slides were used to quantify the percentage of cells with micronuclei. Imaging was performed at room temperature using a wide-field DeltaVision Elite (Applied Precision) microscope with Photometrics Cool Snap HP camera and 100× NA 1.4 Plan Apochromat objective with oil immersion (refractive index = 1.514) using SoftWoRx 3.6 (Applied Precision) software. The acquired images were processed by constrained iterative deconvolution using SoftWoRx 3.6 software package (Applied Precision). Shown images are deconvolved and maximum-intensity projections. Statistical significance of the data was established by a Fisher’s exact test and a Bonferroni correction using R studio.

### Cell viability assays

U2OS human osteosarcoma cells were plated in 96 well plates and 16 h after plating they were transfected with 3.5 pmols of siRNA oligos (Control or SPOUT1/CENP-32) using Lipofectamine siRNA max. 72 h after siRNA transfection, 10 μl of MTT labelling reagent were added to each well (Cell Proliferation kit I-MTT, Roche) and incubated for 4 h at 37°C and 5 % CO_2_. Then 100 μl of Solubilisation buffer (Cell Proliferation kit I-MTT, Roche) were added to each well and incubated at 37 °C for 1 h to allow for total solubilization of the purple formazan crystals. Microplates were evaluated using the Spectramax M5 microplate reader (Molecular devices) at 570 nm with a reference wavelength of 700 nm. Statistical significance of the data was established by a non-parametric ANOVA with Kruskal-Wallis followed by Dunn’s multiple comparisons test using GraphPad Prism version 7.0.

### Size Exclusion chromatography coupled to multi-angle light scattering

SizeLJexclusion chromatography (ÄKTALJMicro™, GE Healthcare) coupled to UV, static light scattering and refractive index detection (Viscotek SECLJMALS 20 and Viscotek RI Detector VE3580; Malvern Instruments) was used to determine the molecular mass of SPOUT1/CENP-32 71-end in solution. 200 μl of 1 mg/ml SPOUT1/CENP-32 71-end (∂A_280 nm_/∂c = 0.667 AU.ml/mg) was run on a Superdex 200 increase 10/300 GL sizeLJexclusion column preLJequilibrated in 50 mM HEPES pH 8.0, 350 mM NaCl and 5 mM DTT at 22°C with a flow rate of 0.5 ml/min. Light scattering, refractive index (RI) and A_280 nm_ were analysed by a homoLJpolymer model (OmniSEC software, v5.02; Malvern Instruments) using the parameters stated for SPOUT1/CENP-32 71-end, ∂*n*/∂c = 0.185 ml/g and a buffer RI value of 1.335. The mean standard error in the mass accuracy determined for a range of protein–protein complexes spanning the mass range of 6–600 kDa is ± 1.9%.

### Western Blot

To determine SPOUT1/CENP-32 levels after siRNA oligo treatment and to test the expression levels of each of the SPOUT1/CENP-32 inducible stable cell lines, U2OS cell lines were transfected in 12-well dishes as described above for the rescue experiments, lysed in 1× Laemmli buffer, boiled for 5 min, and analyzed by SDS-PAGE followed by Western blotting. The antibodies used for the immunoblot were rabbit anti-CENP-32 (HPA022990; 1:250; Atlas Antibodies), rabbit anti-GFP (ab290; 1:5,000, Abcam) and mouse anti-tubulin (ab18251; 1:10,000; Abcam). Secondary antibodies used were goat anti-mouse 680, donkey anti-rabbit 800, and donkey anti-mouse 800 (LI-COR) at 1:2,000. Immunoblots were imaged using the Odyssey CLx system, and band intensities were quantified using ImageJ, uncalibrated OD values. Uncalibrated OD values were corrected by the corresponding tubulin levels (loading control) and normalized to siRNA control values. Three experimental replicates were analyzed.

## Data Availability

All data produced in the present study are available upon reasonable request to the authors

## ACKNOWLEDGEMENTS

We thank Elizabeth LeClair for assistance on statistical analyses of zebrafish data; and Jaewon Lee for help with illustrations in Figure 9. This work was supported by a grant from the US National Institute of Health (NIH) (R01MH106826) to EED, in part by US NIH (NS105078) to JRL and US NIH (UM1 HG011758) to JRL and JEP. SK is funded by an International Research Support Initiative Program (IRSIP) fellowship from the Higher Education Commission of Pakistan. EED is the Ann Marie and Francis Klocke, MD Research Scholar. IS and WCE are funded by Wellcome Principal Research Fellowship to WCE (grant no. 107022). We thank Natalia Kochanova and Shaun Webb for their help with the statistical analyses of the *in vitro* U2OS rescue experiments and we also thank Elizabeth Blackburn for helpful discussions on the Methyltransferase assays. MAA is funded by a grant from the Medical Research Council (MRC, United Kingdom; MR/X001245/1) to AAJ. AAJ and his team are co-funded by the European Union (ERC Advanced Grant, CHROMSEG, 101054950). Views and opinions expressed are, however, those of the authors only and do not necessarily reflect those of the European Union or the European Research Council. Neither the European Union nor the granting authority can be held responsible for them. DM was supported by a medical genetics research fellowship program through the US NIH (T32 GM007526-42). JEP was supported by NHGRI K08 HG008986. TM was supported by the Uehara Memorial Foundation. DP is supported by Doris Duke Charitable Foundation with grant# 2023-0235, and NINDS NS125126-01A1. SNW was supported in part by K08NS119567.

