## Supplemental Table 2 for "RNA methyltransferase SPOUT1/CENP-32 links mitotic spindle organization with the neurodevelopmental disorder SpADMiSS"

**Table S2: Comparison of in silico (CADD, REVEL, AlphaMissense scores ), in vitro (cellular phenotypes rescue), and in vivo (zebrafish head size phenotype rescue) results for *SPOUT1/CENP-32* missense/in-frame deletion variants**

| # | SPOUT1/CENP-32 variant (NM_016390.4) | | Number of families in the cohort (Het/Hom) | gnomAD (v4.0.0) allele frequency | CADD score | REVEL score | AlphaMissense_pathogenicity | AlphaMissense_class | Location in protein structure | Protein stability prediction (mCSM) | MTase activity (Km) vs WT* | MTase activity (Vmax) vs WT[#] | Cellular phenotypes rescue (detached centrosomes and micronuclei) [&] | Zebrafish head size phenotype rescue |
|---|---|---|---|---|---|---|---|---|---|---|---|---|---|---|
| 1 | c.256A>G | p.N86D | 2 (1/1) | 6.84E-07 | 25.8 | 0.35 | 0.8974 | pathogenic | cofactor binding pocket | stabilizing | 1.7 fold increase | 1.6 fold decrease | strong phenotype | failed to rescue |
| 2 | c.292G>A | p.G98S | 8 (1/7) | 8.67E-06 | 28.7 | 0.65 | 0.6527 | pathogenic | dimeric interface | destabilizing | 2 fold increase | 1.4 fold decrease | strong phenotype | failed to rescue |
| 3 | c.590G>A | p.R197Q | 1 (1/0) | 1.10E-05 | 26.6 | 0.31 | 0.1792 | benign | OB fold | destabilizing | not tested | not tested | not tested | not tested |
| 4 | c.598C>T | p.R200W | 1 (1/0) | 1.88E-04 | 25.3 | 0.205 | 0.2595 | benign | OB fold | destabilizing | not tested | not tested | not tested | partial rescue |
| 5 | c.730G>A | p.G244S | 1 (1/0) | 3.18E-06 | 28.2 | 0.559 | 0.8115 | pathogenic | OB fold | destabilizing | 2 fold increase | 1.3 fold decrease | intermediate phenotype | partial rescue |
| 6 | c.766G>C | p.A256P | 1 (1/0) | 2.56E-06 | 25.2 | 0.293 | 0.8516 | pathogenic | OB fold | destabilizing | not tested | not tested | not tested | not tested |
| 7 | c.744_746delATC | p.S249del | 1 (1/0) | 6.16E-06 | N/A | N/A | N/A | N/A | OB fold | N/A | not tested | not tested | not tested | partial rescue |
| 8 | c.866C>T | p.T289M | 1 (1/0) | 2.74E-06 | 25.7 | 0.646 | 0.7424 | pathogenic | cofactor binding pocket | destabilizing | 2 fold increase | 1.4 fold decrease | intermediate phenotype | partial rescue |
| 9 | c.877G>A | p.G293S | 2 (2/0) | 2.54E-05 | 25.5 | 0.575 | 0.8989 | pathogenic | cofactor binding pocket | destabilizing | 1.3 fold increase | 2.1 fold decrease | intermediate phenotype | partial rescue |
| 10 | c.940C>T | p.L314F | 1 (1/0) | 2.23E-05 | 32 | 0.418 | 0.6593 | pathogenic | cofactor binding pocket | destabilizing | not tested | not tested | not tested | partial rescue |
| 11 | c.1016A>G | p.Y339C | 1 (1/0) | 2.75E-06 | 25.6 | 0.62 | 0.7836 | pathogenic | cofactor binding pocket | highly destabilizing | not tested | not tested | not tested | partial rescue |
| 12 | c.1045C>T | p.R349C | 1 (1/0) | 1.43E-05 | 31 | 0.556 | 0.629 | pathogenic | cofactor binding pocket | destabilizing | not tested | not tested | not tested | not tested |
| 13 | c.1054C>T | p.R352C | 1 (1/0) | 1.62E-05 | 27.7 | 0.606 | 0.9672 | pathogenic | cofactor binding pocket | destabilizing | not tested | not tested | not tested | partial rescue |
| 14 | c.1058C>T | p.T353M | 2 (0/2) | 4.99E-06 | 25.3 | 0.635 | 0.7127 | pathogenic | cofactor binding pocket | stabilizing | 1.5 fold increase | 1.6 fold decrease | strong phenotype | partial rescue |

N/A: not applicable

mCSM: https://biosig.lab.uq.edu.au/mcsm/stability

* an increase in Km indicates lower affinity for the substrate

[#] a decrease in Vmax indicates a decrease in the rate of the reaction when the enzyme is saturated with its substrate

[&] intermediate phenotype: centrosome detachment only / strong phenotype: centrosome detachment + spindle and centrosomal abnormalities
