## Supplemental Table 1 for "RNA methyltransferase SPOUT1/CENP-32 links mitotic spindle organization with the neurodevelopmental disorder SpADMiSS"

### Table S1: Summary of clinical and genetic findings in 24 individulas from 18 families with bi-allelic SPOUT1/CENP-32 variants

| Subject ID | # | Referring/ Testing Center | ES/GS testing | *SPOUT1/CENP-32* variant 1 | SPOUT1/CENP-32 variant 2 | Previous genetic testing | Ancestry | Consanguinity | Gender | Prenatal Complications | Age at last evaluation | Weight at last evaluation (percentile/z-score) | Height at last evaluation (percentile/z-score) | OFC at last evaluation (percentile/z-score) | Short stature | Microcephaly | Poor weight gain | ID | DD | Hypotonia | Hypertonia/spasticity | Seizures | Seizure type | EEG tracings reviewed | abnormal brain MRI or U/S findings | Eye abnormalities | Feeding issues | Dysmorphic features | Other findings | Family History |
|---|---|---|---|---|---|---|---|---|---|---|---|---|---|---|---|---|---|---|---|---|---|---|---|---|---|---|---|---|---|---|
| A-III-2 | 1 | Columbia University Irving Medical Center | Trio ES | c.292G>A (p.G98S) | c.730G>A (p.G244S) | Karyotype, CMA, mitochondrial studies normal, *HPD* and *TAT* gene sequencing negative | European | no | female | | | | | | | | | | | | | | | | | | | | | |
| B-II-3 | 2 | Institute of Human Genetics, Leipzig, Germany | Singleton ES | c.1058C>T (p.T353M) | c.1058C>T (p.T353M) | CMA normal; homozygosity mapping identified two candidate regions on chromosome 9 and 16 of 13.9 Mb and 2.7 Mb, respectively | unk | yes | male | | | | | | | | | | | | | | | | | | | | | |
| B-II-7 | 3 | Institute of Human Genetics, Leipzig, Germany | Singleton ES | c.1058C>T (p.T353M) | c.1058C>T (p.T353M) | CMA normal; homozygosity mapping identified two candidate regions on chromosome 9 and 16 of 13.9 Mb and 2.7 Mb, respectively | unk | yes | male | | | | | | | | | | | | | | | | | | | | | |
| C-II-2 | 4 | CeGaT Center for Human Genetics Tuebinge, Germany | Trio ES | c.102dupA (p.W35Mfs*77) | c.256A>G (p.N86D) | karyotype, Rett-syndrome panel negative | German/Malaysian | no | female | | | | | | | | | | | | | | | | | | | | | |
| D-III-5 | 5 | Children's Hospital of Philadelphia, PA | Trio ES | c.877G>A (p.G293S) | c.940C>T (p.L314F) | prenatal NIPS, CMA normal | unk | no | male | | | | | | | | | | | | | | | | | | | | | |
| E-V-6 | 6 | Nationwide Children's Hospital, Columbus, OH | Trio GS | c.292G>A (p.G98S) | c.292G>A (p.G98S) | CMA showed 5% AOH, 92kb VUS loss in 3q11.2, CHAt sequencing was negative, DNA methylation for PWS/AS negative | Yemen | yes | male | | | | | | | | | | | | | | | | | | | | | |
| F-II-1 | 7 | Children's National Hospital, DC | Trio ES | c.744_746delATC (p.S249del) | c.866C>T(p.T289M) | none | Afghanistan | no | female | | | | | | | | | | | | | | | | | | | | | |
| F-II-2 | 8 | Children's National Hospital, DC | No | c.744_746delATC (p.S249del) | c.866C>T(p.T289M) | none | Afghanistan | no | female | | | | | | | | | | | | | | | | | | | | | |
| G-II-1 | 9 | Baylor Hopkins Center for Mendelian Genomics | Duo ES | c.1058C>T (p.T353M) | c.1058C>T (p.T353M) | plasma amino acids and acyl carnitien profile normal, urine organic acids showed 4-5 fold increase in succinic acid levels, karyotype normal, 15qFISH for AWS/PS normal | Turkey | no | female | | | | | | | | | | | | | | | | | | | | | |
| G-II-2 | 10 | Baylor Hopkins Center for Mendelian Genomics | No | c.1058C>T (p.T353M) | c.1058C>T (p.T353M) | unk | Turkey | no | male | | | | | | | | | | | | | | | | | | | | | |
| H-II-1 | 11 | Columbia University Irving Medical Center | Trio ES | c.1016A>G(p.Y339C) | c.1054C>T(p.R352C) | unk | unk | unk | male | | | | | | | | | | | | | | | | | | | | | |
| I-II-1 | 12 | Columbia University Irving Medical Center | Trio ES | c.598C>T(p.R200W) | c.877G>A(p.G293S) | unk | unk | unk | male | | | | | | | | | | | | | | | | | | | | | |
| J-II-1 | 13 | CHU Bordeaux, France | Trio ES | c.590G>A(p.R197Q) | c.766G>C(p.A256P) | unk | French | unk | female | | | | | | | | | | | | | | | | | | | | | |
| K-II-1 | 14 | John Hopkins Aramco Health Care, Dhahran, Saudi Arabia. | Singleton ES | c.292G>A(p.G98S) | c.292G>A(p.G98S) | unk | Saudi Arabia | yes | male | | | | | | | | | | | | | | | | | | | | | |
| L-II-1 | 15 | Shahid Chamran University of Ahvaz; Narges Medical Genetics and Prenatal | Singleton ES | c.292G>A(p.G98S) | c.292G>A(p.G98S) | normal biochemical | Iran (Arab) | yes | female | | | | | | | | | | | | | | | | | | | | | |
| L-II-2 | 16 | Shahid Chamran University of Ahvaz; Narges Medical Genetics and Prenatal Diagnosis Laboratory; Golestan Medical, Educational, and Research Center; Jundishapur University of Medical Sciences, Ahvaz, Iran | No | c.292G>A(p.G98S) | c.292G>A(p.G98S) | unk | Iran (Arab) | yes | female | | | | | | | | | | | | | | | | | | | | | |
| M-V-2 | 17 | Clinical Genetics Department, Human Genetics and Genome Research Division, National Research Centre, Cairo, Egypt | Singleton ES | c.292G>A(p.G98S) | c.292G>A(p.G98S) | Normal karyotyping, extended metabolic screening | Egyptian | yes | female | | | | | | | | | | | | | | | | | | | | | |
| N-IV-2 | 18 | Clinical Genetics Department, Human Genetics and Genome Research Division, National Research Centre, Cairo, Egypt | Singleton ES | c.292G>A(p.G98S) | c.292G>A(p.G98S) | unk | Egyptian | yes | female | | | | | | | | | | | | | | | | | | | | | |
| O-II-1 | 19 | Department of Translational Genomics, Center for Genomic Medicine, King Faisal Specialist Hospital and Research Center, Riyadh, Saudi Arabia | Singleton ES | c.292G>A(p.G98S) | c.292G>A(p.G98S) | unk | Saudi Arabia | unk | female | | | | | | | | | | | | | | | | | | | | | |
| P-III-3 | 20 | Pediatric Department, Faculty of Medicine, Tanta University, Egypt | Singleton ES | c.292G>A(p.G98S) | c.292G>A(p.G98S) | unk | Egyptian | yes | female | | | | | | | | | | | | | | | | | | | | | |
| P-III-4 | 21 | Pediatric Department, Faculty of Medicine, Tanta University, Egypt | No | c.292G>A(p.G98S) | c.292G>A(p.G98S) | unk | Egyptian | yes | female | | | | | | | | | | | | | | | | | | | | | |
| Q-II-1 | 22 | Department of Development Pediatrics, The Children's Hospital and The Institute of Child Health, Multan, Pakistan | Singleton ES | c.256A>G (p.N86D) | c.256A>G (p.N86D) | unk | Pakistani | unk | male | | | | | | | | | | | | | | | | | | | | | |
| Q-II-2 | 23 | Department of Development Pediatrics, The Children's Hospital and The Institute of Child Health, Multan, Pakistan | No | c.256A>G (p.N86D) | c.256A>G (p.N86D) | unk | Pakistani | unk | male | | | | | | | | | | | | | | | | | | | | | |
| R-II-1 | 24 | Ambry Genetics,Aliso Viejo, CA | Trio ES | c.1017C>A(p.Y339*) | c.1045C>T (p.R349C) | CMA, cerebral palsy panel, neuromuscular disorder panel, SA, urine organic acids, acylcarnitine, lactate, pyruvate: normal | African American/Asian | no | female | | | | | | | | | | | | | | | | | | | | | |

**Detailed clinical information in these columns have been removed from the preprint version in compliance with medRxiv policy.**

ADHD: attention deficit hyperactivity disorder; CF: cystic fibrosis; CMA: chromosomal microaray; CSF: cerebrospinal fluid; DD: developmental delay; EEG: electroencephalogram; ES: exome sequencing; GS: genome sequencing; ID: intellectual disability; MRI: magnetic resonance imaging; NIPS: noninvasive prenatal screening; NR: not reported; OFC: occipital frontal circumference; PWS/AS: Prader-Willi syndrome/Angelman syndrome; unk: unknown; U/S: ultrasound; VUS: variant of uncertain significance
