## Supplementary Information file for "RNA methyltransferase SPOUT1/CENP-32 links mitotic spindle organization with the neurodevelopmental disorder SpADMiSS"

**Case reports (Family A – Family R)**

(*This section has been removed from the preprint version in compliance with medRxiv policy*.)

***Spout1* is expressed in neocortical progenitors and is required for cell cycle progression in mice**

In our cohort, we find that microcephaly is seen in greater than 80% of individuals when head circumference measurements were available. Therefore we reasoned that disruption of neuronal progenitor’s proliferation and/or differentiation during embryonic development of the brain could be the reason for this pathology. First we tested if *Spout1* is expressed in the neuronal progenitors of the developing neocortex. In situ hybridization with *Spout1* synthetic antisense RNA was performed on brain sections of E14.5, Е16.5 and E18.5 mouse embryos. At E14.5, the hybridization signal was detected in the proliferative compartments, the ventricular and subventricular zones (VZ and SVZ). At E16.5 and E18.5 the expression of *Spout1* was also detected in the VZ/SVZ. Additionally, Spot1 signal was also detected in the compartment of differentiating neurons, the cortical plate (CP), but was weaker in the compartment of young migrating neurons, the intermediate zone (IZ). These data indicate that *Spout1* is expressed in the cortical progenitors through entire corticogenesis. It seems to be downregulated in the migrating neurons, and activated again at later stages of cortical development, when young neurons have settled in the CP (Figure S7A-D).

In order to down-regulate *Spout1* expression in the cortical progenitors *in vivo*, we used *in utero* electroporation of plasmid constructs expressing shRNA against *Spout1* mRNA into the lateral ventricles of E13.5 mouse embryos (Figure S8). To trace electroporated cells they also were co-electroporated with a plasmid containing GFP cDNA. 24 hours later, females pregnant with electroporated embryos have been injected with a single pulse of a synthetic nucleoside analogue of thymidine, Bromodeoxyuridine (BrdU). BrdU incorporates into DNA of dividing cells at S phase of the mitotic cycle. 24 hours after BrdU injection, embryos were isolated and their brains were analyzed for presence of cells double positive for GFP and a marker of proliferating cells Ki67. Double positive cells in such experiments represent a fraction of neocortical cells that incorporated targeting constructs, and still remain in the cell cycle 48 hours after electroporation. The percentage of such cells in the control electroporation experiments accounts for 20% of all GFP positive cells. In the case of the shRNA electroporation, the proportion of cells that did not leave mitotic cycle was doubled, as comparted to the control conditions (Figure S7E-F). On the other hand, GFP/BrdU/Ki67 triple positive cells represent a fraction of electroporated progenitors that remain cycling 24 hours after BrdU injection (cells labeled at S phase of the mitotic cycle). Here, also the proportion of cells that started to differentiate was lower than in the control experiments (33% vs 48%).

These results indicate that *Spout1* in the forebrain neuronal stem cells is required for proper cell cycle progression and when its expression is downregulated, dividing cells remain in the cell cycle longer and less frequently begin neuronal differentiation. This eventually might result in fewer numbers of neurons generated in the neocortex.

**Cloning of targeting vectors**

Primer design was performed with NEBuilder assembly Tool version 2.8.2, an HA tag (HA-Human influenza hemagglutinin is a surface glycoprotein) was added to the given sequence after the ATG codon. Primers were purchased from Metabion. A murine cortex cDNA library was used as template and amplification of the reading frame was done using PrimeSTAR GXL DNA Polymerase from Takara. The pCAG-IRES-GFP-vector was digested with EcoRI-HF (New England Biolabs) in Cutsmart buffer. DNA of PCR and digested vector were separated in 2% Agarose 1x TAE electrophoresis, dissected from the Gel and purified using NucleoSpin Gel and PCR Clean-up mini kit (Macherey-Nagel). The DNA fragments were eluted in 30 µl water. 2.5 µl of PCR fragment, 0.5 µl of purified vector and 3 µl of NEBuilder® HiFi DNA Assembly (New England Biolabs) were incubated at 50°C for 30 min, and then transferred on ice. 50µl ultracompetent E. coli XL10 Gold (Agilent Technology) were added, the mix was incubated at 42°C for 45 sec, cooled down on ice for 10 sec and then directly spread on LB-Agar containing 100 µg/ml Ampicillin. After 14 hours at 37°C, colonies were picked and used to inoculate 5 ml of 1x LB-Medium with 100 µg/ml ampicillin, shaking at 37°C for 14 hours with 180 rpm.

For shRNA expression, the following primers were designed and ordered from Metabion,

(shSpout1a GATCCCCGTGGAAGGACCTCAAGATAATTCAAGAGATTATCTTGAGGTCCTTCCACTTTTT

shSpout1b AGCTAAAAAGTGGAAGGACCTCAAGATAATCTCTTGAATTATCTTGAGGTCCTTCCACGGG

shSpout2a GATCCCCGGACCTCAAGATAATGAAGAATCAAGAGTTCTTCATTATCTTGAGGTCCTTTTT

shSpout2b AGCTAAAAAGGACCTCAAGATAATGAAGAACTCTTGATTCTTCATTATCTTGAGGTCCGGG

shSpout3a GATCCCCGGTCAACTGTGGAATGAAGAATCAAGAGTTCTTCATTCCACAGTTGACCTTTTT

shSpout3b AGCTAAAAAGGTCAACTGTGGAATGAAGAACTCTTGATTCTTCATTCCACAGTTGACCGGG

shSpout4a GATCCCCGAATGAAGAAGGAGGTGAAGATCAAGAGTCTTCACCTCCTTCTTCATTCTTTTT

shSpout4b AGCTAAAAAGAATGAAGAAGGAGGTGAAGACTCTTGATCTTCACCTCCTTCTTCATTCGGG

shSpout5a GATCCCCGAAGGAGGTGAAGATTGACAATCAAGAGTTGTCAATCTTCACCTCCTTCTTTTT

shSpout5b AGCTAAAAAGAAGGAGGTGAAGATTGACAACTCTTGATTGTCAATCTTCACCTCCTTCGGG

shSpout6a GATCCCCAAGAAACTGGACCCTGGACTTTCAAGAGAAGTCCAGGGTCCAGTTTCTTTTTTT

shSpout6b AGCTAAAAAAAGAAACTGGACCCTGGACTTCTCTTGAAAGTCCAGGGTCCAGTTTCTTGGG)

Analyzed and confirmed plasmids were used for transfection and in utero electroporation experiments.

**Investigating the role of *spout1/cenp-32* in *C. elegans***

To better understand the *in vivo* functions of *SPOUT1/CENP-32*, we studied the role of the *SPOUT1/CENP-32* ortholog in *C. elegans*. The *C. elegans* ortholog, B0361.6, was an unnamed gene without a single associated published study. We renamed the gene to spot-1 for SPOuT domain containing RNA methyltransferase homolog. To determine its function, we took advantage of two independent putative null alleles, tm6282 and tm6390, that delete 404bp and 263bp, respectively (Figure S12A). Homozygous strains could not be isolated suggesting both alleles were lethal. To quantitate the lethality, each strain was crossed with the balancer strain hT2 that is marked with a pharyngeal GFP reporter ^[1]^. hT2 is homozygous lethal so the presence of GFP indicates a heterozygous strain, while a GFP negative strain indicates the respective null allele is homozygous (Figure S12B). The balancer strain in a WT background (i.e. N2) gave rise to viable GFP negative animals (Figure S12C), whereas both null alleles gave rise to extremely rare GFP negative animals, indicating both alleles are nearly 100% lethal (Figure S12C). We attempted to rescue this lethal phenotype by introducing spot-1 as an extrachromosomal array, but we failed to isolate any lines suggesting overexpression of spot-1 can also be detrimental. These experiments demonstrated that spot-1 is essential and the level of expression maybe critical for proper development.

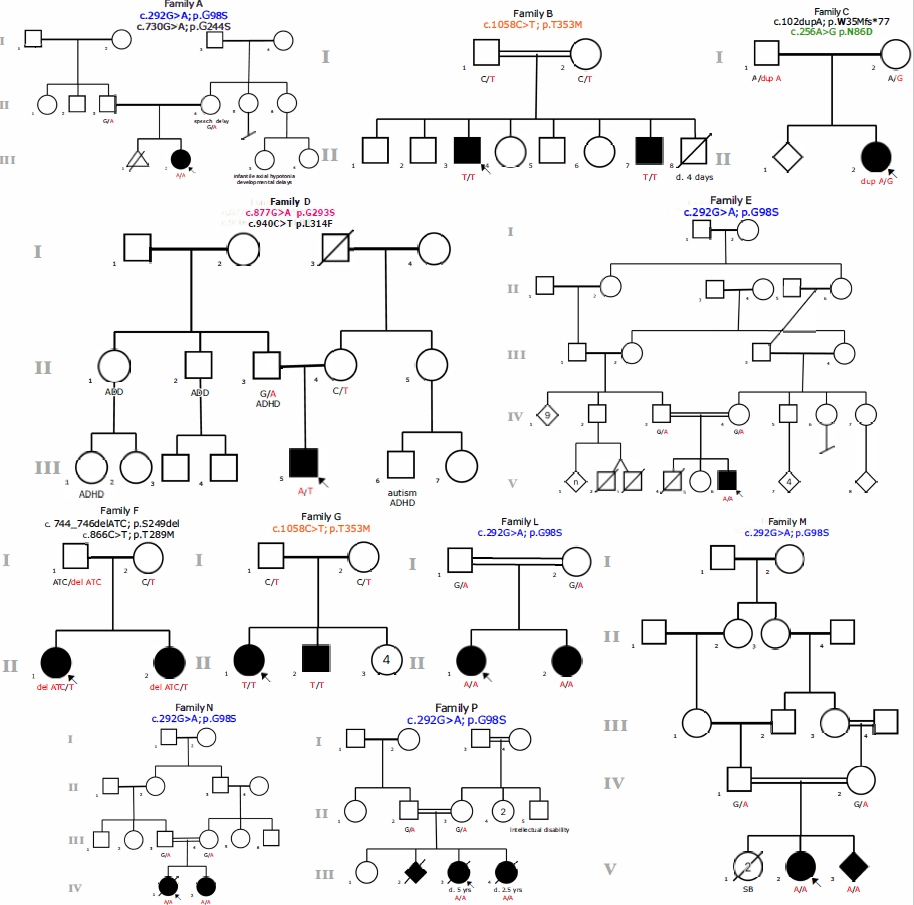

**Pedigrees have been removed from the preprint version in compliance with medRxiv policy.**

**Figure S1: Pedigrees of families A, B, C, D, E, F, G, L, M, N, and P.** Affected individuals are shown as filled symbols and variant status listed for each tested individual. Relevant clinical features for family members are listed. Variants alleles are shown in red. Double-lines indicate consanguineous relationship; SB: still birth; ADD: attention deficit disorder; ADHD: attention deficit hyperactivity disorder.

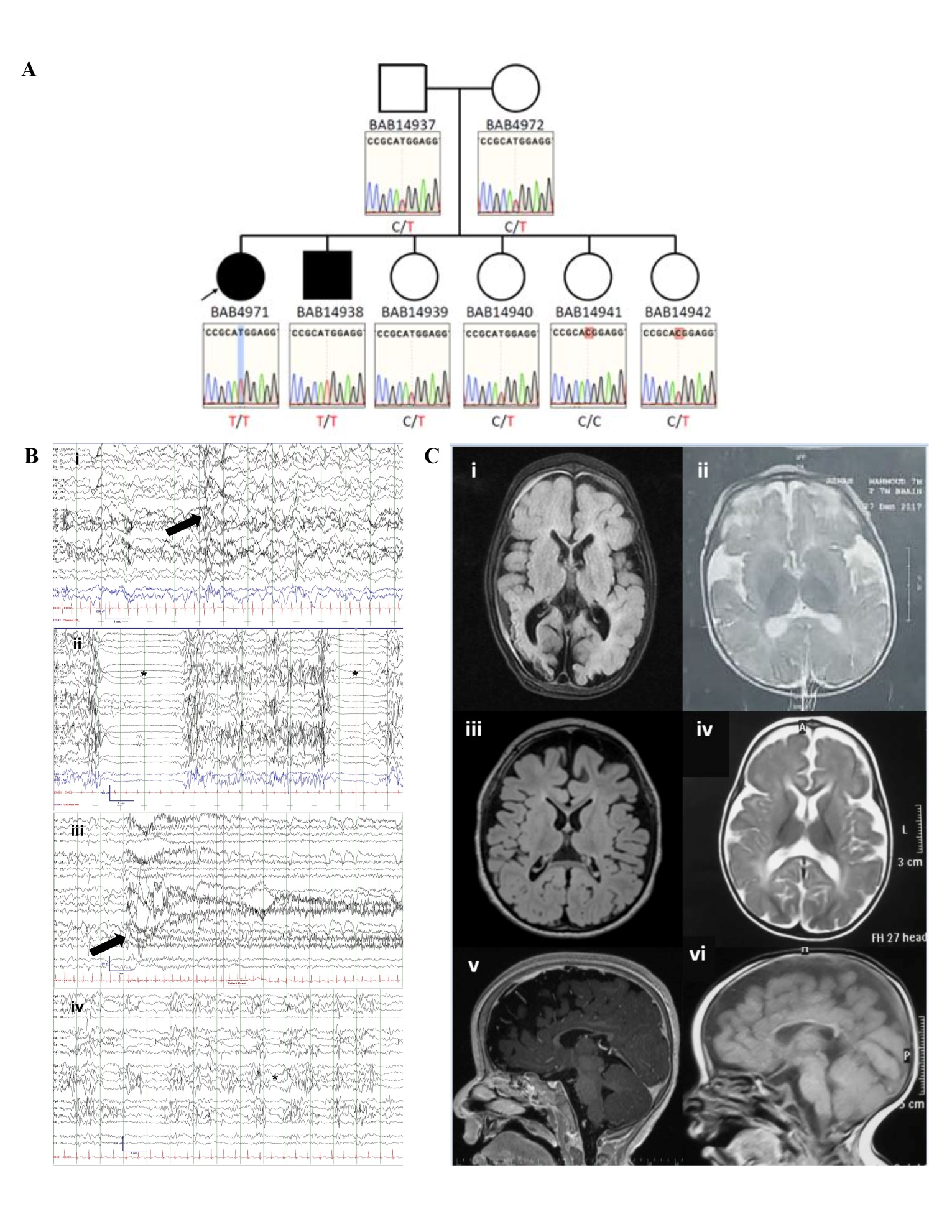
**Figure S2: Sanger sequencing, EEG, and brain MRI results.**

**Pedigree and Sanger results of Family G have been removed from the preprint version in compliance with medRxiv policy.**

(A) Sanger confirmation and segregation in family G of the *SPOUT1/CENP-32*: c.1058C>T (p.T353M) variant identified in subjects G-II-1 and G-II-2 by ES.

(B) Representative ictal and interictal EEG tracings for subject A-III-2 at ages 1-5 years in chronological order when awake (i;iii), and asleep (ii;iv). The awake background is disorganized. The sleep EEG is discontinuous with abundant multifocal spikes. Asterisks mark examples of discontinuity. Arrows mark the electrographic onset of seizures (epileptic spasm in i; tonic seizure in iii).

(C) Representative MRI of the brain. Axial T2 images from subjects L-II-1 (ii) and M-V-2 (iv), and T2 FLAIR images from subjects A-III-2 (i) and E-V-6 (iii) at ages 6 months -5 years showing cerebral atrophy, marked by increased extra-axial spaces and ex vacuo expansion of the lateral ventricles and diminished white matter displaying abnormal T2 signal hyperintensity. Subject A-III-2 also shows laminar necrosis in the right posterior. Sagittal T1 images from subjects E-V-6 (v) and M-V-2 (vi) demonstrating thinning of the corpus callosum.

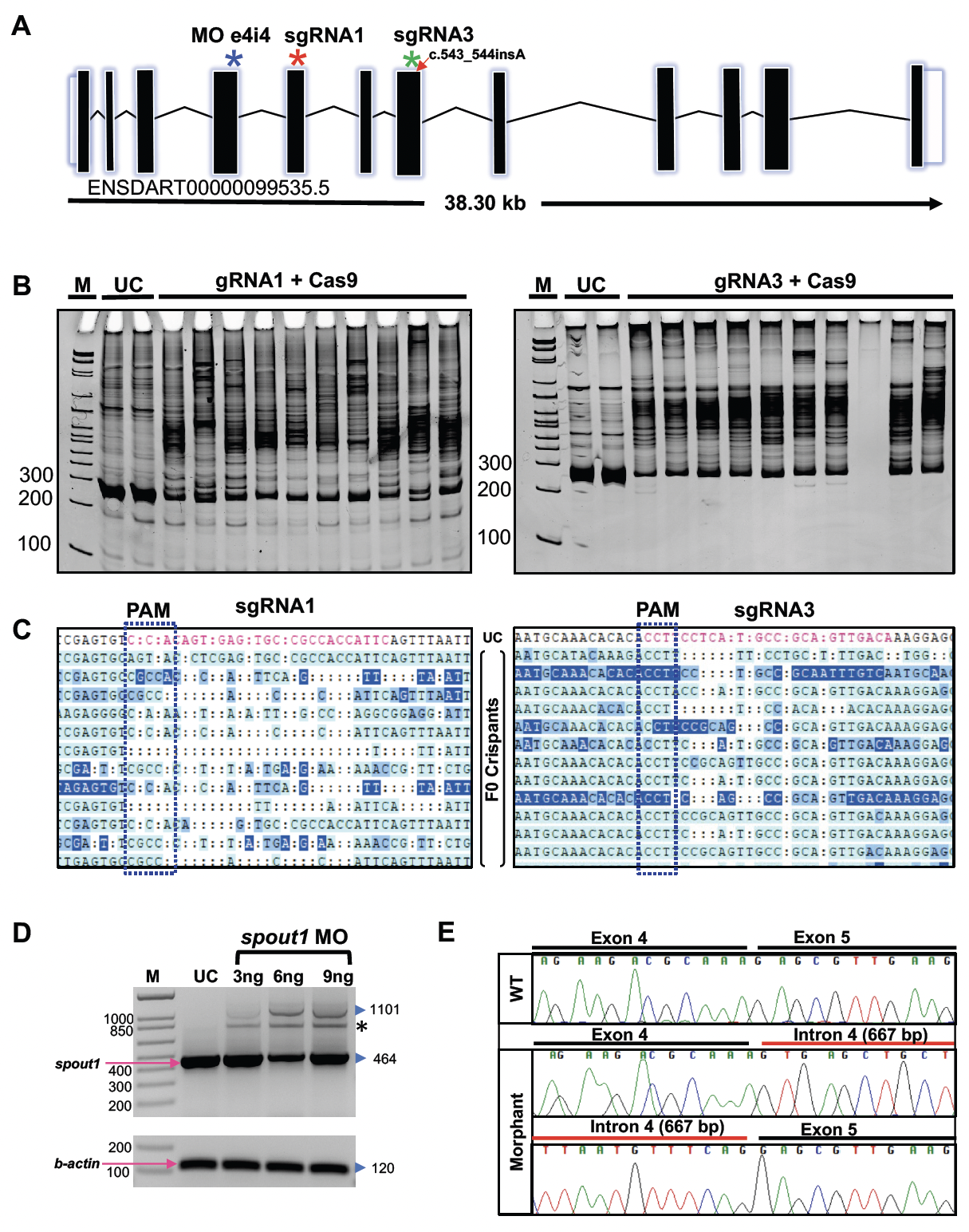

**Figure S3: *spout1/cenp-32* genome editing, and transient suppression reagents induce efficient targeting in zebrafish.**

(A) Schematic representation of the *D. rerio spout1/cenp-32* locus (GRCz11); black vertical boxes, exons; zig-zag horizontal line, introns; white vertical boxes, untranslated regions. Target regions of reagents are shown as asterisks; splice blocking morpholino (MO) at the exon 4 splice donor site, blue; single guide (sg) RNA 1 at exon 5, red; and sgRNA3 at exon 7, green; red arrow at exon 7 shows the position of the 1 bp insertion in stable mutants.

(B) Heteroduplex analysis of F0 mosaic mutants. Polyacrylamide gel electrophoresis (20% PAGE) indicates targeting events in F0 embryos. M, marker (with size labels at left); UC, uninjected control.

(C) Representative chromatograms generated from PCR-amplified product flanking the sgRNA target sites and cloned into TOPO-TA vector. Protospacer adjacent motif (PAM) is shown with blue dotted boxes. sgRNAs induce high mosaicism of frameshifting indels at each respective target (sgRNA1: 81%; sgRNA3: 78%).

(D) RT-PCR analysis of *spout1/cenp-32* morphants. 1% agarose gel image shows aberrantly spliced transcript induced by MO targeting the splice donor site of exon 4, resulting in an intron retention (667 bp) in morphants. Expected fragment size for WT, 464 bp. (*) is a heteroduplex of the WT fragment and intron-retained fragment. *b-actin* (expected fragment size 120 bp) was amplified to ensure RNA integrity.

(E) Sequence chromatograms of RT-PCR products from morphants indicate retention of intron 4.

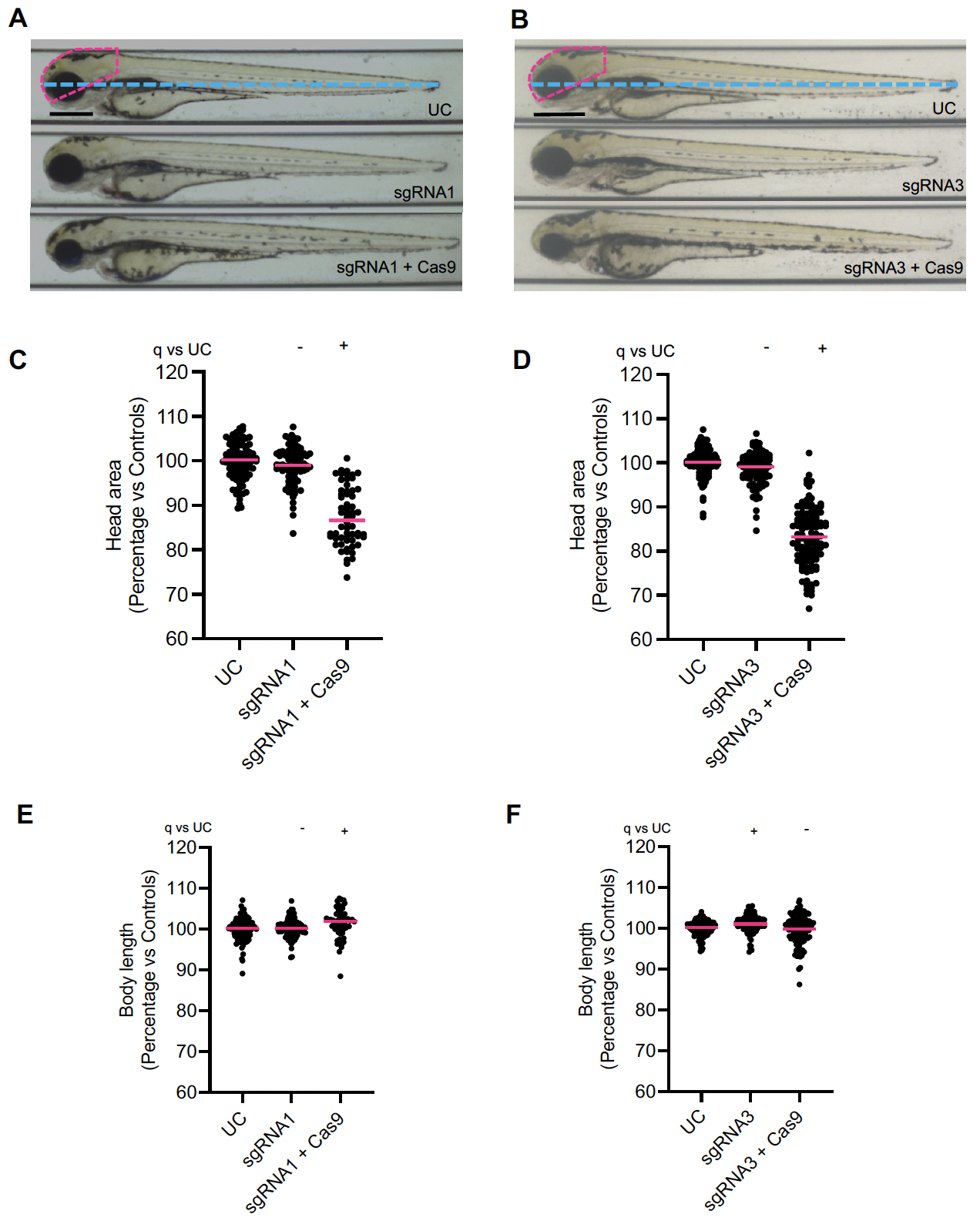

**Figure S4: CRISPR/Cas9 genome editing of *spout1/cenp-32* leads to reduced head size in F0 mosaic mutants.**

(A, B) Representative bright field lateral images of uninjected controls (UC) and F0 crispant larvae (injected with sgRNA1 or sgRNA3) acquired at 3 dpf; pink dashed outline depicts head size measured, and the blue dotted line shows the body length measured.

(C-F) quantification of lateral head size and body length measurements from combined experimental batches injected with sgRNA1 (n=2 biological replicates) and sgRNA3 (n=3 replicates). Each scale bar, 300 µm. Statistical differences were calculated using non-parametric ANOVA with Kruskal-Wallis test followed by Dunn’s multiple comparisons test by controlling False Discovery Rate (original FDR method of Benjamini and Hochberg); (+) and (-) indicates significant and non-significant differences, respectively. Median values are shown with pink horizontal lines. See Table S3 for exact adjusted q-values and numbers of larvae.

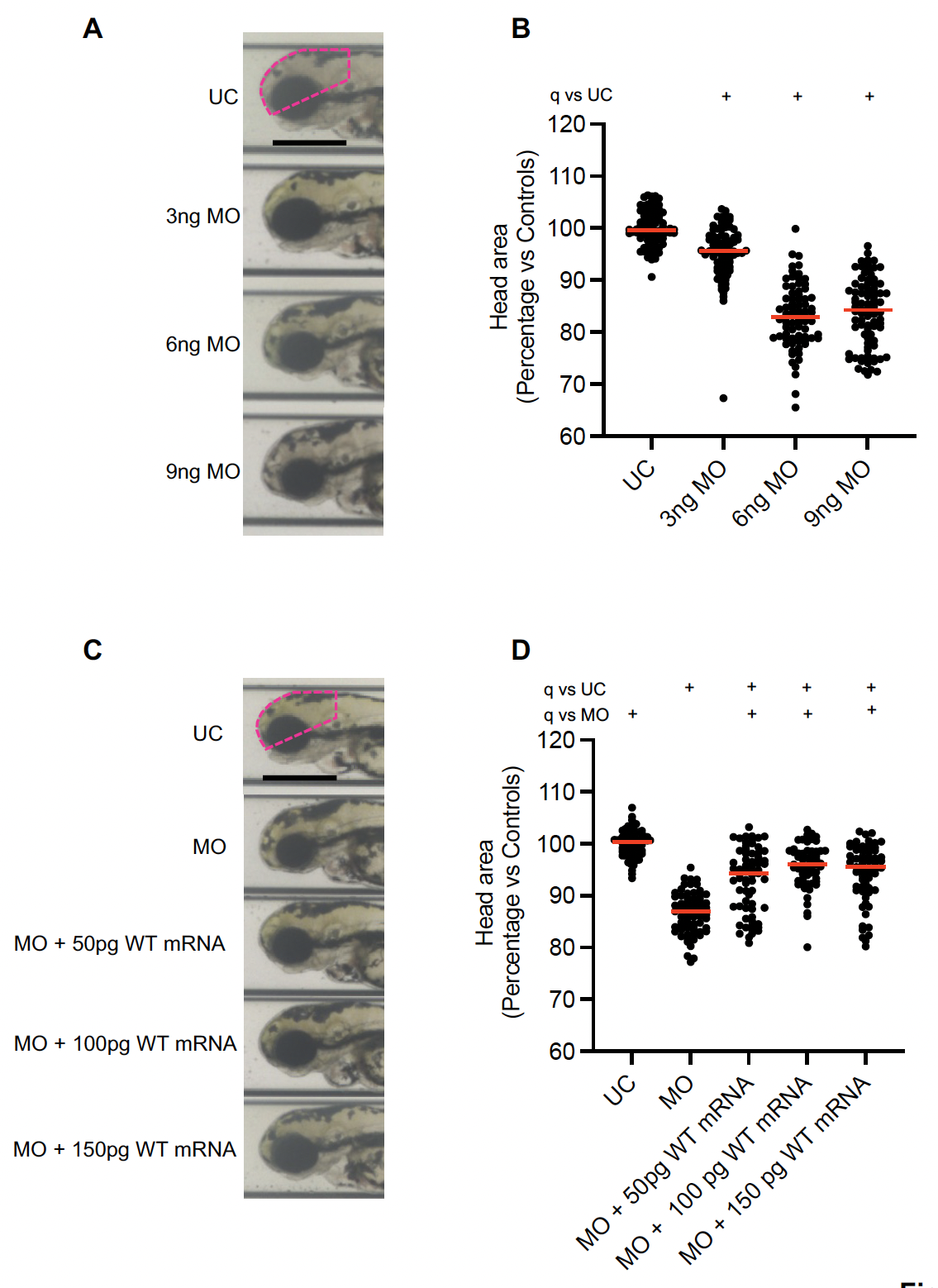

**Figure S5 *spout1/cenp-32* transient suppression and ectopic expression reagents were optimized for *in vivo* complementation assays.**

(A) Representative lateral bright field images of larvae injected with 3ng, 6ng and 9ng MO; pink dashed line indicates the area measured.

(B) Lateral head size measurements were analyzed from two independent biological replicate experiments.

(C) Representative bright field lateral images show significant rescue with increasing doses of WT *SPOUT1* mRNA.

(D) Quantification of lateral head size from two independent biological replicate experiments. Scale bar in all panels, 300 µm. Statistical differences were calculated using non-parametric ANOVA with Kruskal-Wallis test followed by Dunn’s multiple comparisons test by controlling False Discovery Rate (original FDR method of Benjamini and Hochberg); (+) and (-) indicate significant and non-significant differences, respectively. Median values are shown with pink horizontal lines. See Table S3 for exact adjusted q-vales and numbers of larvae.

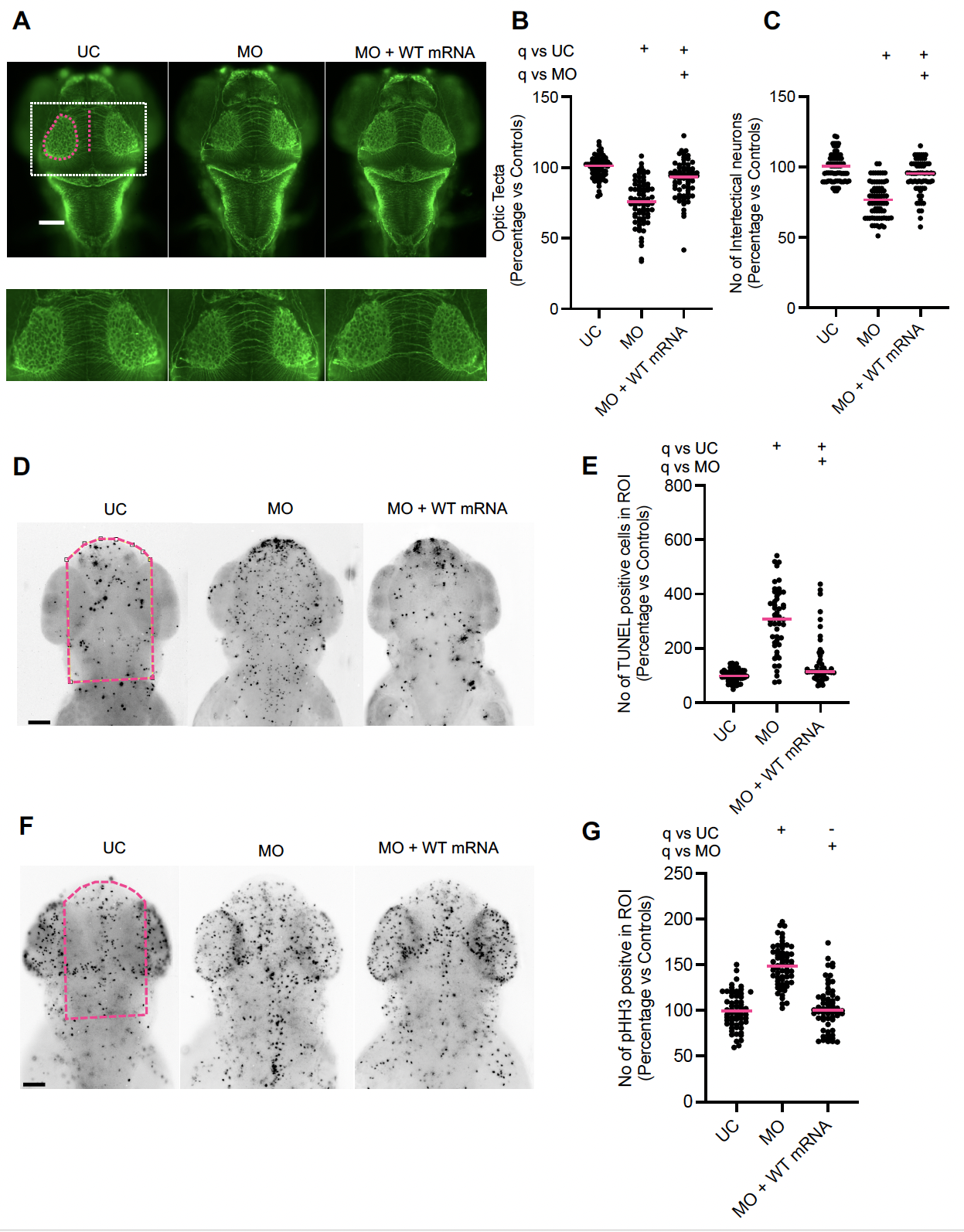

**Figure S6: Suppression of *spout1/cenp-32* induces CNS defects, apoptosis and altered cell cycle progression in zebrafish larvae.**

(A) Representative dorsal images of wholemount larvae fixed at 3 dpf and immunostained for acetylated tubulin to demarcate axon tracts. We counted commissural axons crossing the midline between the optic tecta (pink dashed line); and the area of optic tecta (pink dashed oval). Dashed white box (upper panel) represents magnified image in lower panel. Scale bar, 100 µm. UC, uninjected control; MO, morphant; WT, wild type.

(B, C) Quantification of optic tecta size and intertectal neuron number, respectively.

(D) Representative dorsal inverted fluorescent images of zebrafish larvae marked by TUNEL at 2 dpf. The region of interest (ROI) quantified is shown with a pink dotted rectangle. Scale bar, 100 µm.

(E) Quantification of TUNEL stained cells as measured in the ROI

(F). Representative dorsal inverted fluorescent images showing phospho-histone H3 (pHH3) positive cells at 2 dpf in controls and morphants. The ROI quantified is shown with a pink dotted rectangle. Scale bar, 100 µm (g) Quantification of pHH3 positive cells as measured in ROI. Data shown in B, C, E, G are combined from two biological replicates. Statistical differences were calculated using non-parametric ANOVA with Kruskal-Wallis test followed by Dunn’s multiple comparisons test by controlling False Discovery Rate (original FDR method of Benjamini and Hochberg); (+) and (-) indicate significant and non-significant differences, respectively. Median values are shown with pink horizontal lines. See Table S3 for exact adjusted q-values and numbers of larvae.

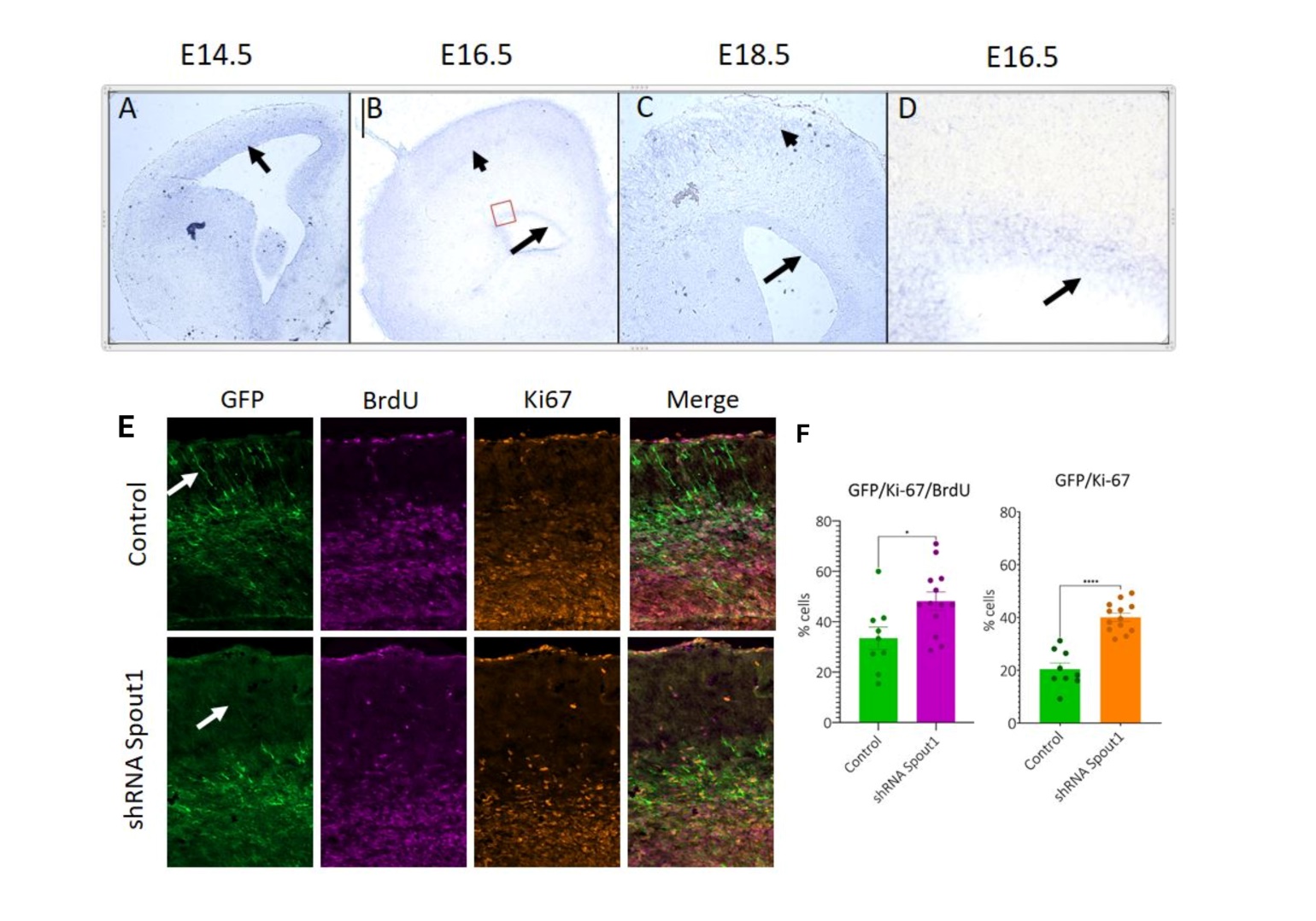

**Figure S7: *Spout1* is expressed in neocortical progenitors and down-regulation of *Spout1* in the cortical stem cells causes cell cycle progression disruption.**

(A-D) *Spout1* is expressed both in the progenitors of the developing forebrain during embryonic development (depicted by arrows) and in late differentiating neurons (depicted by small arrows). D is a high magnification image of the boxed area on B.

(E-F) Cortical progenitors electroporated with shRNA against Spout1 less frequently leave mitotic cycle both within 24 hours (left diagram) and 48 hours (right diagram) and migrate to the CP (white arrows) than wild type cells. (Note GFP positive neurons in the control CP and lack of them in shRNA electroporated brains).

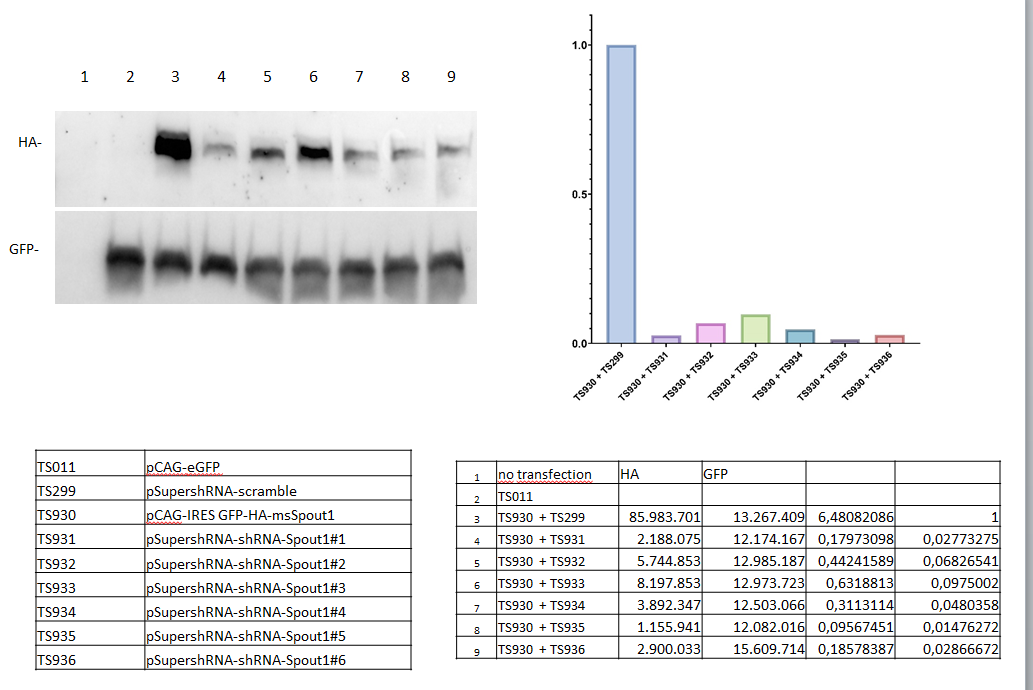

**Figure S8: Western blot analysis of knock-down efficiency of six shRNA containing constructs against Spout1 in Neuro2A cells.**

Protein detection with HA antibody in Neuro2A cells after transfection with overexpression constructs for HA-tagged mouse-Spout1 and GFP. Neuro2A cells transfected either with GFP overexpression plasmid only, GFP and a combination of HA-tagged Spout1 overexpression plasmids in combination with a control scramble shRNA expressing vector or six different shRNAs 1-6 against mouse Spout1. Each sample was transfected with 1 µg of DNA in total, 0.1 µg of pCAG-eGFP. Levels of GFP expression for transfection efficiency and HA Tag representing the amount of expressed mouse Spout1 protein are shown. For Detection of HA tag, 1:5000 purified mouse anti-HA.11 Epitope Tag Biolegend (902302) was used, for the detection of the GFP protein, 1:5000 goat anti-GFP polyclonal Rockland (600-101-215) was applied. Secondary antibodies were purchased from Dianova: HRPO-IgG (H+L) Donkey Anti-GOAT, 705-035-147 and HRPO-IG (H+L) Donkey Anti-Mouse, 610-703-124, both were diluted 1:100 000 in 3% BSA in TBS-T. Western-Blot detection was done using SuperSignal Wester Dura Extended Duration Substrate (ThermoFisher #34075), a ChemiDoc XRS device from BioRad and Software version 6.1. Quantification of signal intensity was done using Fiji Image J software v1.53t and values have been computed in Microsoft excel. The graph represents the quantification of the blot shown here. Most efficient down regulation of mouse Spout1 protein was detected with shRNA vectors 1 and 5.

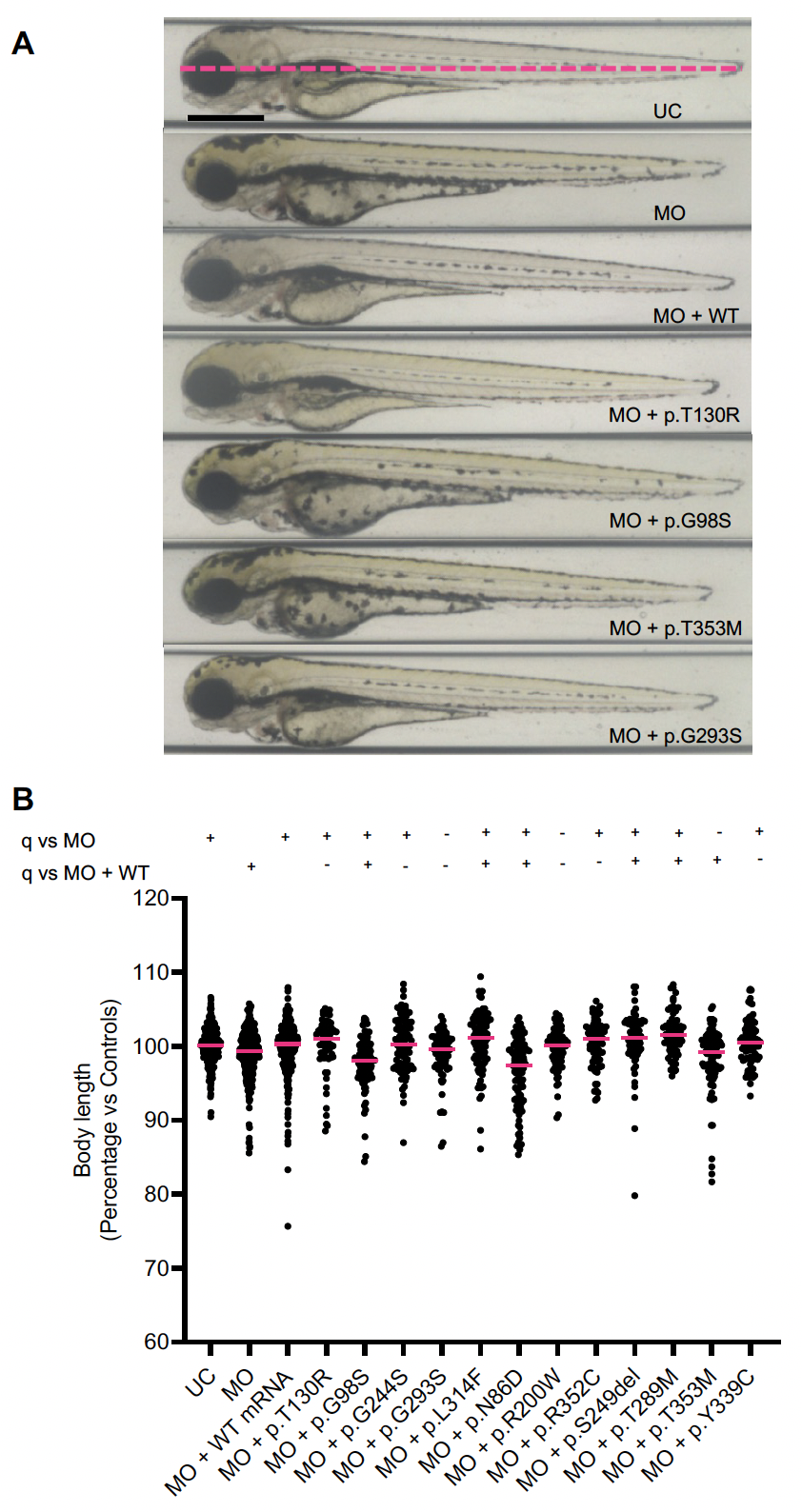

**Figure S9: Reduction of *spout1/cenp-32* in zebrafish does not induce a significant effect on body length.**

(A) Representative bright field lateral images from *in vivo* complementation assays were acquired at 3 dpf.

(B) Quantification of body length from at least 3 independent biological replicates is shown. Scale bar, 300 µm. Statistical differences were calculated using non-parametric ANOVA with Kruskal-Wallis test followed by Dunn’s multiple comparisons test by controlling False Discovery Rate (original FDR method of Benjamini and Hochberg); (+) and (-) indicates significant and non-significant differences, respectively. Median values are shown with pink horizontal lines. See Table S3 for exact adjusted q-values and numbers of larvae.

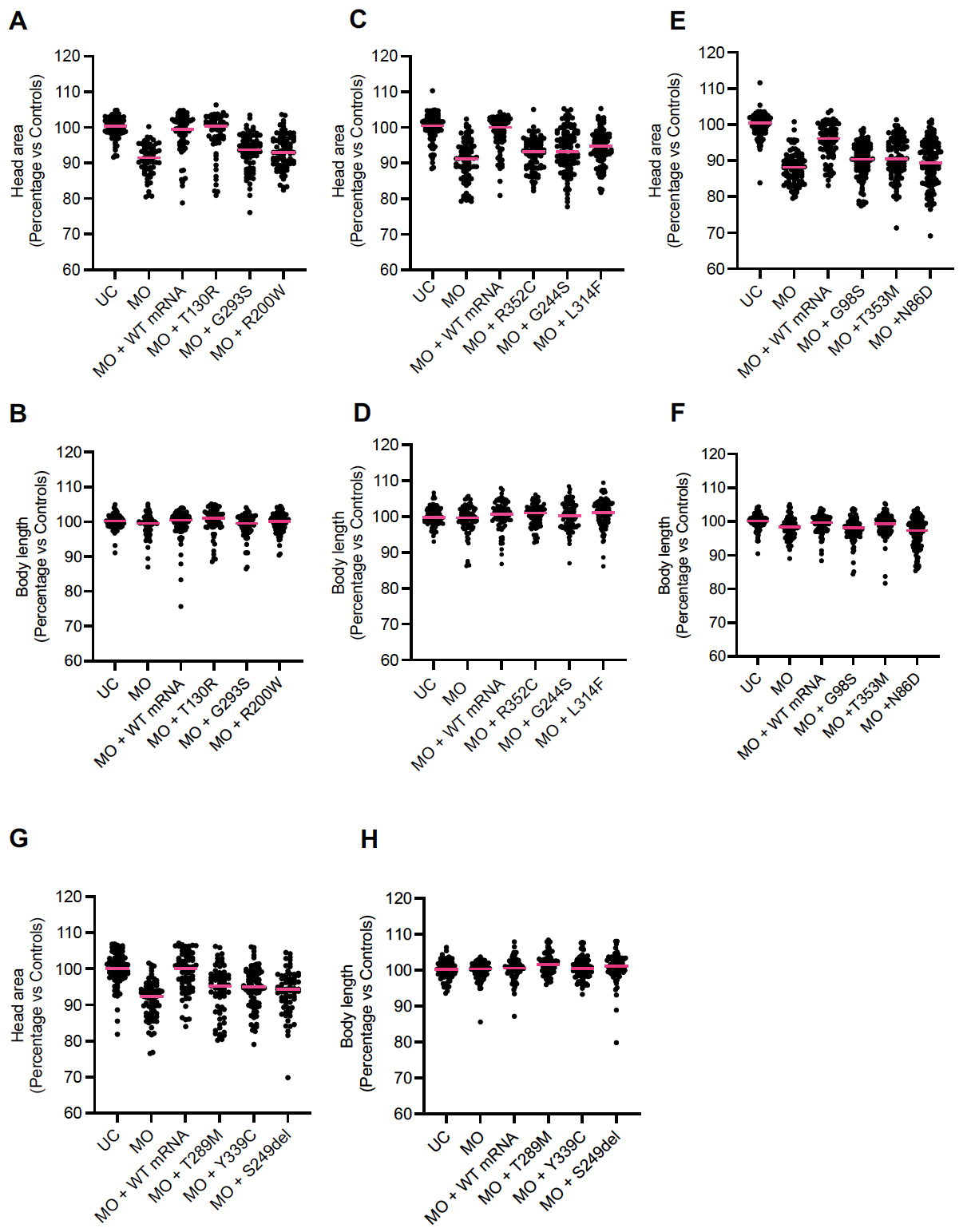

**Figure S10: Supporting data for Figure 3 and Figure S9; dot plots from independent experiments quantifying head size and body length measurements.** Each dot plot represents triplicate biological experiments.

(A, C, E, G) Dot plots correspond to *in vivo* complementation studies of head size (normalized to controls and combined in Figure 3B)

(B, D, F, H) Dot plots correspond to *in vivo* complementation studies of body length (normalized to controls and combined in Figure S9B). Median values are shown with pink horizontal lines. See Table S3 for exact adjusted q-values and numbers of larvae.

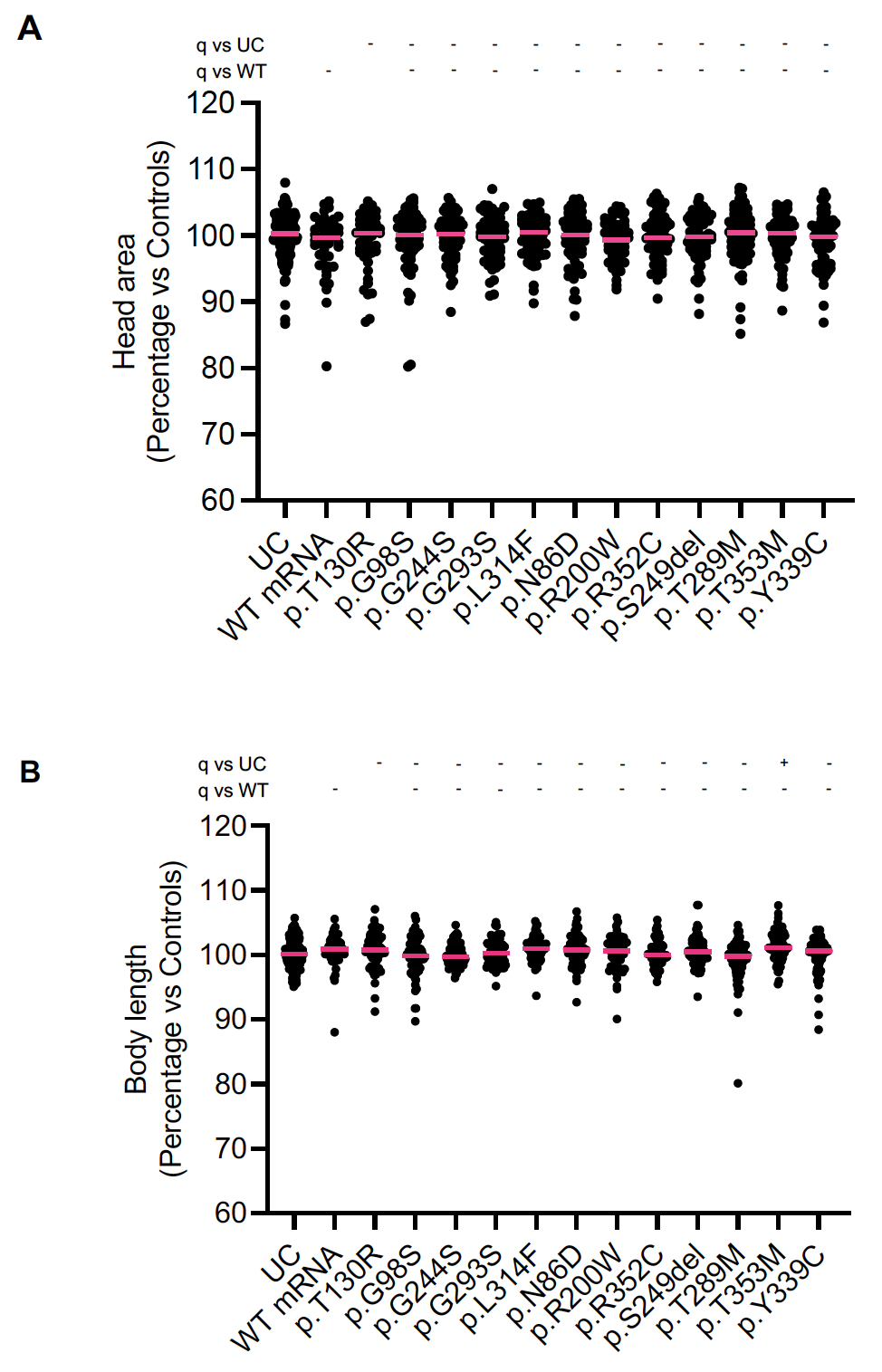

**Figure S11: Heterologous expression of human wild type or variant *SPOUT1/CENP-32* mRNA does not induce head size or body length phenotypes in zebrafish.**

(A, B) Quantification of head area or body length imaged live in 3 dpf larvae injected with capped human WT or variant mRNA. Data shown are combined from two independent biological replicates and normalized to uninjected controls (UC). See Figure 3 and Figure S9 for measurement strategies. Statistical differences were calculated using non-parametric ANOVA with Kruskal-Wallis test followed by Dunn’s multiple comparisons test by controlling False Discovery Rate (original FDR method of Benjamini and Hochberg); (+) and (-) indicate significant and non-significant differences, respectively. Median values are shown with pink horizontal lines. See TableS3 for exact adjusted q-values and numbers of larvae.

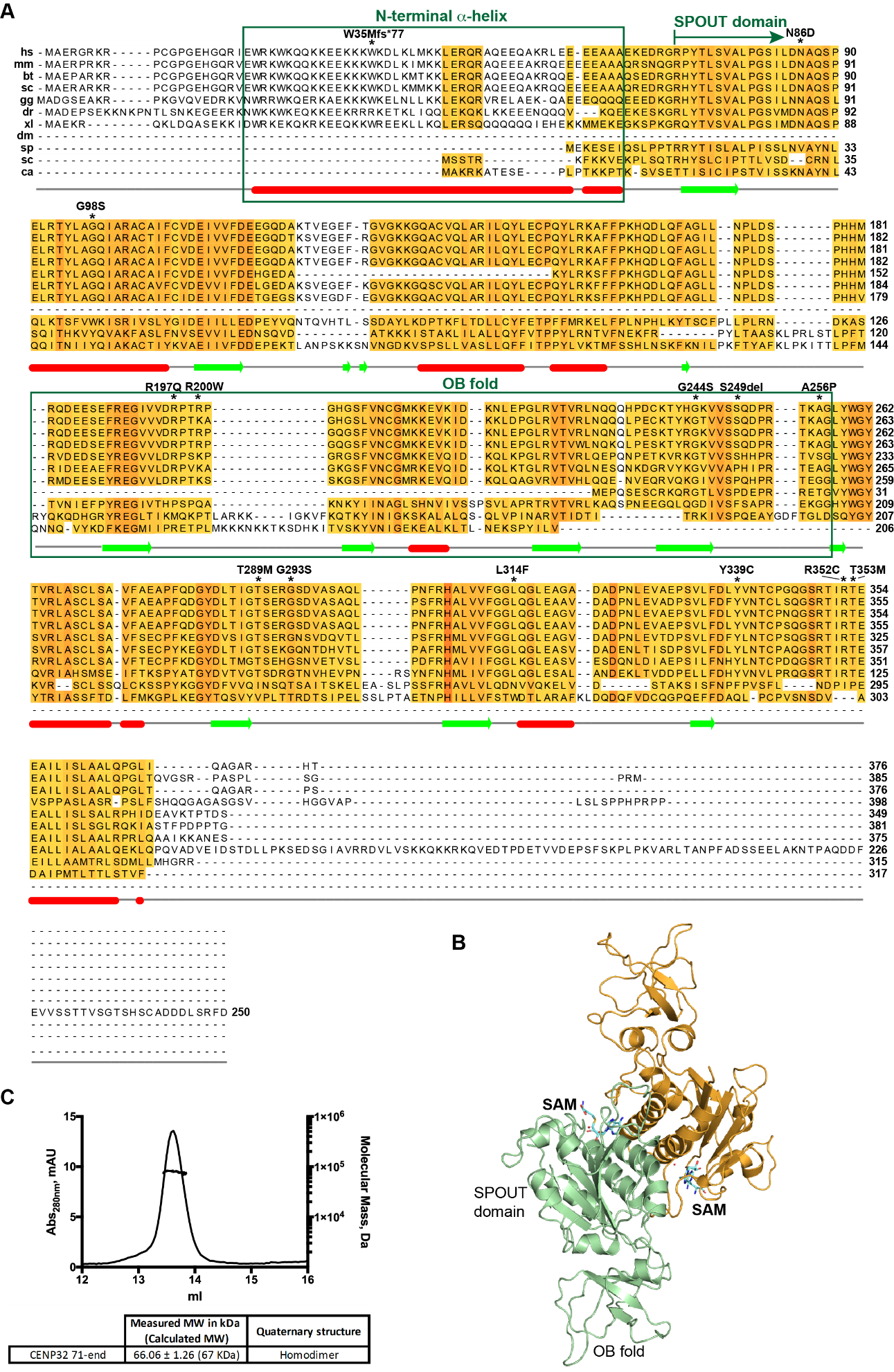

**Figure S12: SPOUT1/CENP-32 is a highly conserved methyltransferase that forms dimers.**

(A) Sequence alignment of SPOUT1/CENP-32 orthologues from *Homo sapiens* (hs), *Mus musculus*(mm), *Bos taurus* (bt), *Sus scrofa* (sc), *Gallus gallus* (gg), *Danio rerio* (dr), *Xenopus laevis* (xl), *Drosophila melanogaster* (dm), *Schizosaccharomyces pombe* (sp), *Saccharomyces cerevisiae* (sc) and *Candida albicans* (ca). The conservation score is mapped from red (highly conserved) to yellow (poorly conserved). Highlighted with green boxes are the positively charged N-terminal alpha helix and the OB fold. The N-terminal alpha helix is present only in higher eukaryotes. The start of the highly conserved SPOUT domain (R71) that exists until the end of the protein (T376) is shown as a green arrow. The positions of patient variations are marked with asterisks. Predicted secondary structure elements are shown below the sequence alignment. Multiple sequence alignment was performed with Clustal Omega (EMBL-EBI) and edited with Jalview 2.11.0 ^[2]^.

(B) Cartoon representation of the crystal structure of SPOUT1/CENP-32 bound to SAM. The main domains of SPOUT1/CENP-32 (SPOUT domain and OB fold) are highlighted.

(C) SEC-MALS of SPOUT1/CENP-32 71-376. Absorption at 280 nm (mAU, left *y*-axis) and molecular mass (kDa, right *y*-axis) are plotted against the elution volume (ml, *x*-axis). The measured molecular weight (MW) and the calculated subunit stoichiometry based on the predicted MW of different subunit compositions are shown in the table below the graph. Samples were analyzed using a Superdex 200 increase 10/300 GL column in 50 mM HEPES pH 8.0, 350 mM NaCl and 5 mM DTT at 22 °C.

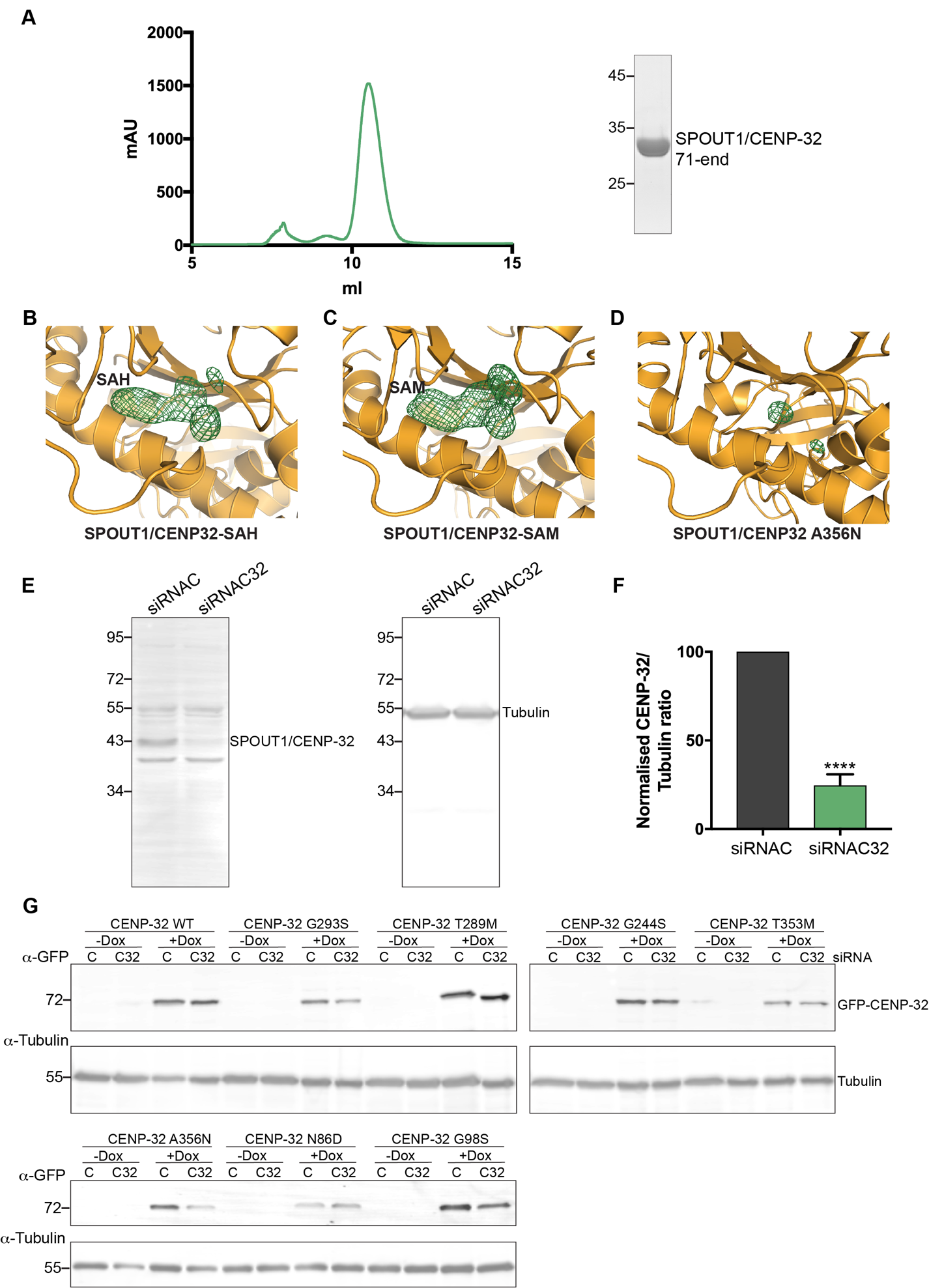

**Figure S13: Analysis of SPOUT1/CENP-32 purity used for crystallization and levels of SPOUT1/CENP-32 depletion and expression in U2OS inducible cell lines.**

(A) Prior to the crystallisation trials, SPOUT1/CENP-32 71-end was purified using a Superdex 75 10/300 GL (Cytiva) in 20 mM Tris pH 8, 200 mM NaCl and 5 mM DTT. The corresponding chromatogram (left) and SDS-PAGE analysis of the main peak of the chromatogram (right) are shown. mAU, milli absorbance units.

(B, C and D) Close-up of the cofactor binding pocket for the crystal structure of SPOUT1/CENP-32 71-376 bound to SAH (B), SPOUT1/CENP-32 71-376 bound to SAM (C) and SPOUT1/CENP-32 71-376 A356N (D) showing the electron density map of the cofactor in both SAH and SAM structures, and the lack of density in the A356N structure. Structural analysis shows that as predicted, the A356N mutation leads to an empty cofactor pocket and can act as a catalytic site mutant.

(E) Representative Immunoblot for the analysis of endogenous SPOUT1/CENP-32 levels upon SPOUT1/CENP-32 depletion using siRNA oligonucleotides.

(F) Quantification of SPOUT1/CENP-32/Tubulin ratio using uncalibrated OD values (normalized SPOUT1/CENP-32/Tubulin ratio for siRNA CENP-32 is 24.6 ± 3.6) was performed with ImageJ. Three independent experiments; mean ± SD; unpaired two-sided *t* test; ****, P ≤ 0.0001.

(G) Representative Immunoblot of SPOUT1/CENP-32-GFP constructs (CENP-32-GFP WT, CENP-32-GFP A356N, CENP-32-GFP N86D, CENP-32-GFP G98S, CENP-32-GFP G293S, CENP-32-GFP T289M, CENP-32-GFP G244S or CENP-32-GFP T353M) and the corresponding tubulin signal as a loading control, showing comparable expression levels.

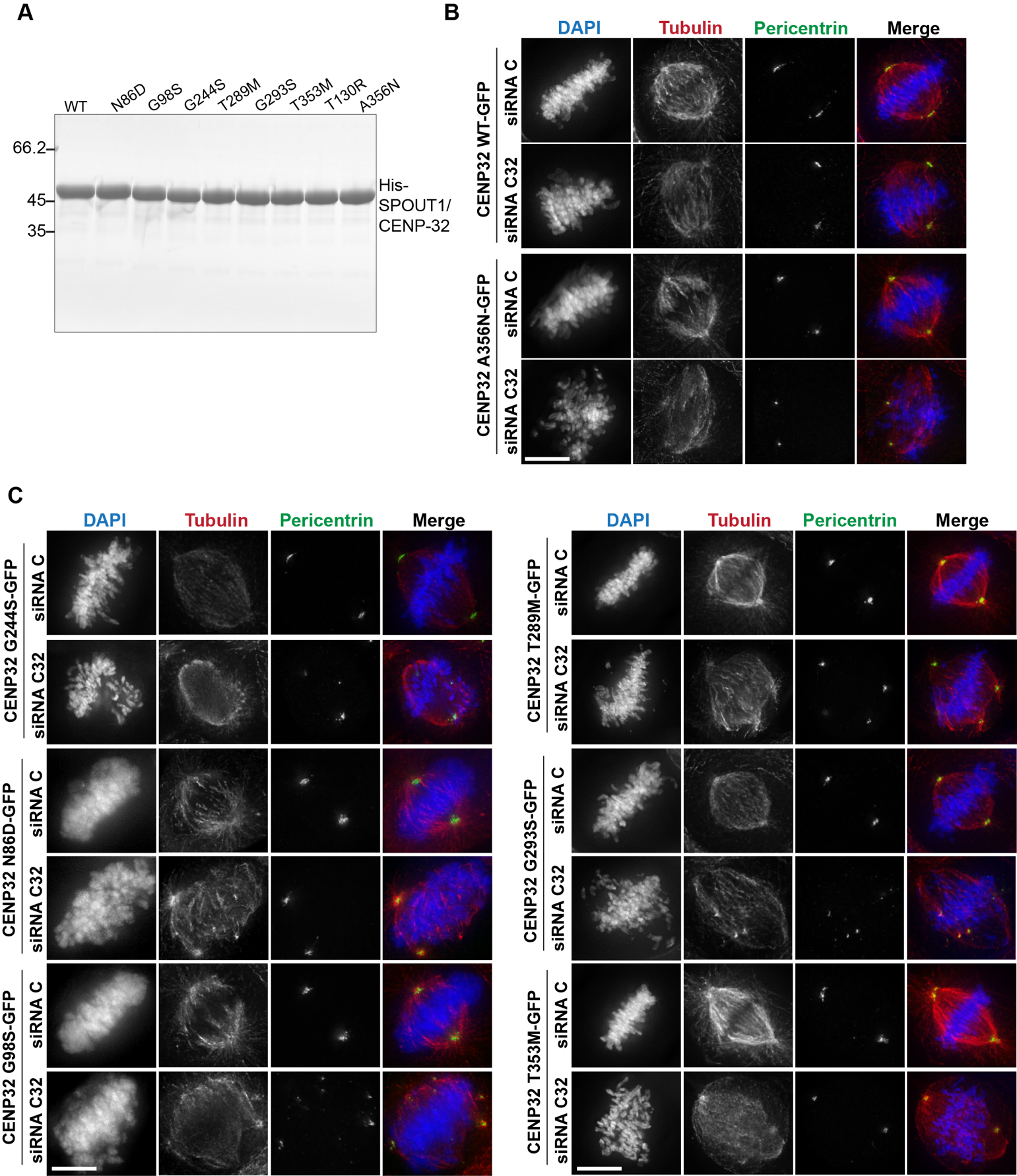

**Figure S14: Analysis of SPOUT1/CENP-32 stable cell lines, conditions without doxycycline.**

(A) SDS-PAGE analysis of SPOUT1/CENP-32 WT and SPOUT1/CENP-32 patient variants proteins used in methyltransferase assays shown in Fig. 6B and 7B.

(B and C) Representative fluorescence images of a SPOUT1/CENP-32 siRNA (C32) rescue assay in the stable U2OS cell lines with inducible expression of different SPOUT1/CENP-32 constructs. Conditions without doxycycline from Fig. 6C (B) and from Fig. 8A (C).

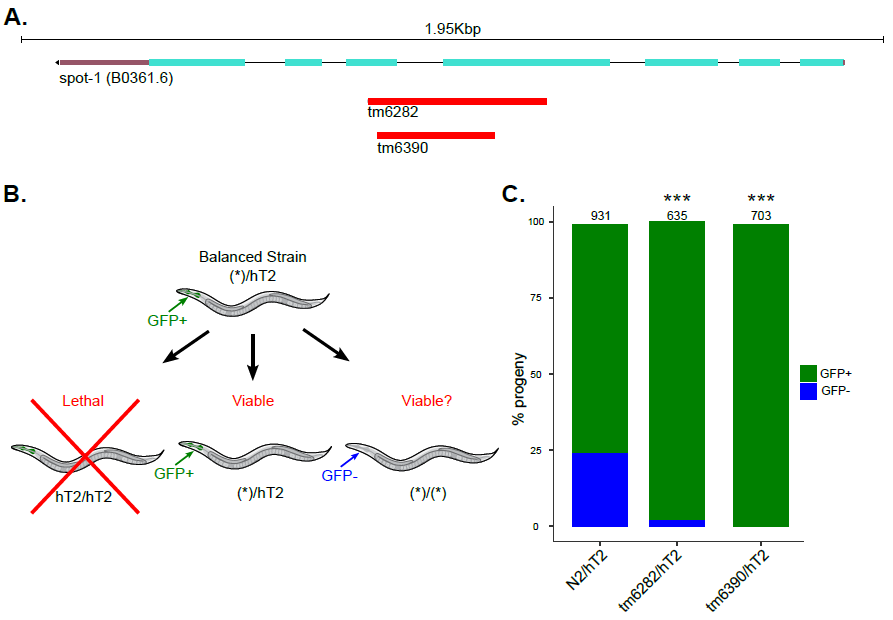

**Figure S15: The *C. elegans SPOUT1/CENP-32* ortholog (spot-1, formerly known as B0361.6) is essential**

(A) Genomic map of spot-1 (B0361.6) is shown. Two independent null alleles (i.e. tm6282 & tm6390) are depicted showing the region of the locus that are deleted, respectively.

(B) A schematic is shown for the resulting progeny of a balanced strain for an allele of interest (indicated as (*)) using the hT2 balancer. hT2 is marked with a myo-2p::GFP so the presence of GFP indicates the presence of the balancer. Animals that are homozygous for hT2 are lethal. Animals with GFP represent strains that are heterozygous for hT2 and the allele of interest (*). GFP- animals represent homozygous strains of the allele of interest (*).

(C) The percent of progeny that are GFP+ or GFP- is shown for the indicated strains. Total number of progeny counted for each indicated strain is shown by the number above the bar graph, respectively. Chi-squared test was performed comparing WT (N2) balanced strain with tm6282 & tm6390 balanced strains. *** indicates p-value of <0.0001.

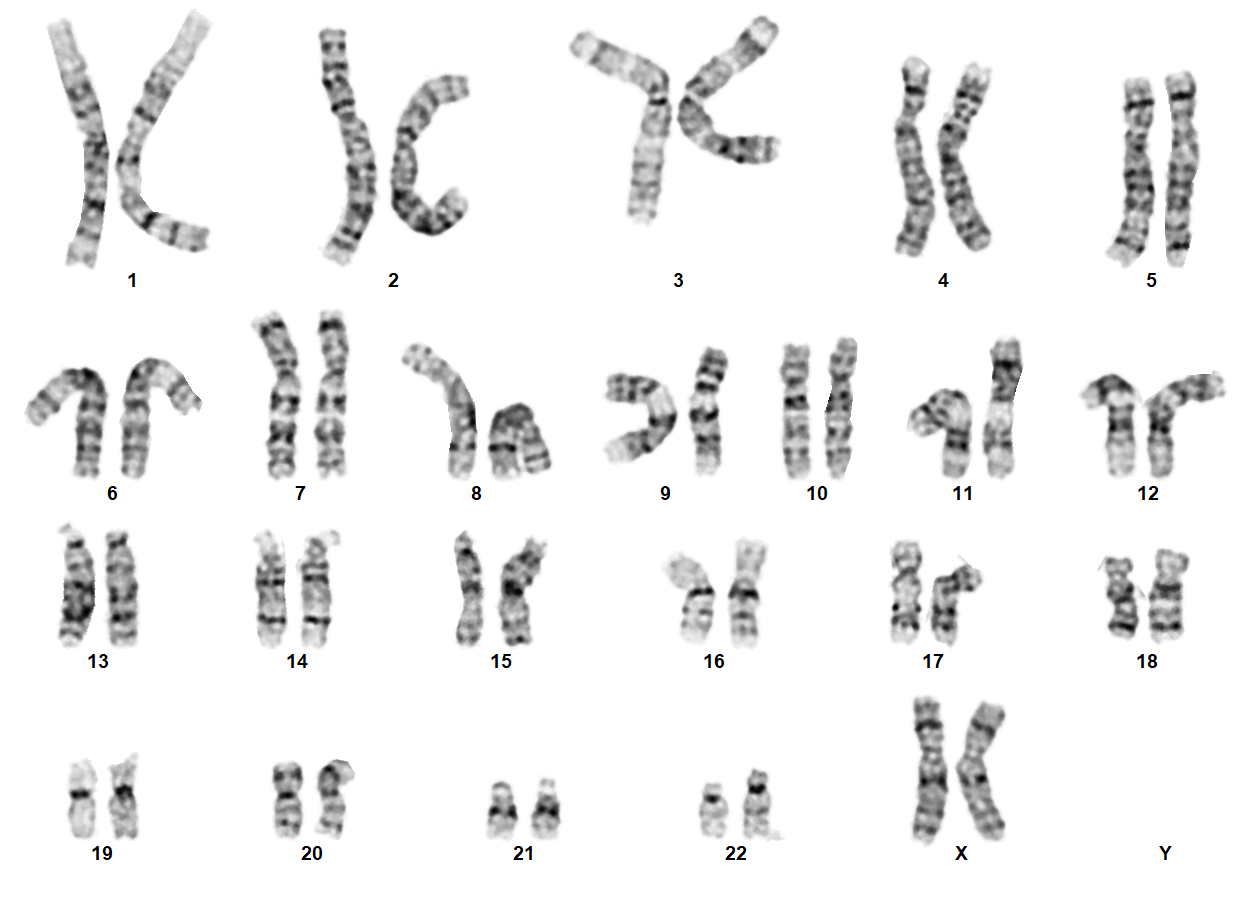

**Figure S16: Karyotype analysis for Subject A-III-2.**

Chromosome analysis for Subject A-III-2 showing a normal 46,XX karyotype in 50 cultured peripheral blood lymphocytes. Karyotypes were interpreted according to the International System for Human Cytogenetic Nomenclature (2020). The banding resolution is at 550-band-level.

**Table S3: Primers used for *spout1/cenp-32 in vivo* modeling in zebrafish**

| Primer Name | Purpose | Nucleotide sequence (5' to 3') |
| --- | --- | --- |
| **Morpholino and CRISPR sequences** | | |
| *spout1*MO e4i4 | MO induced suppression | GTCAAAATAGCAGCTCACTTTGCGT |
| *spout1* RT-PCR_F | MO efficiency | ATACAGTGAGCGTGGCTCTG |
| *spout1* RT-PCR_R | MO efficiency | CTCTCAGTCCCGTTTTCAGC |
| *b*-*actin* RT-PCR-F | Internal control | TTACCACTTCACGCCCGACTC |
| *b-actin* RT-PCR-R | Internal control | GTCACCTTCACCGTTCCAGT |
| *spout1* sgRNA1-F | Genome editing | TAATACGACTCACTATA**GAATGGTGGCGGCACTCACTG** |
| *spout1* sgRNA1-R | Genome editing | TTCTAGCTCTAAAAC**CAGTGAGTGCCGCCACCATT** |
| *spout1* sgRNA1 PCR-F | CRISPR/Cas9 efficiency | CCCAATGAGAAAAACCGTTC |
| *spout1* sgRNA1 PCR-R | CRISPR/Cas9 efficiency | GAGGATGAGATGAAGGCAGAA |
| *spout1* sgRNA3-F | Genome editing | TAATACGACTCACTATA**TGTCAACTGCGGCATGAGGA** |
| *spout1* sgRNA3-R | Genome editing | TTCTAGCTCTAAAAC**TCCTCATGCCGCAGTTGACA** |
| *spout1* sgRNA3 PCR-F | CRISPR/Cas9 efficiency | GTGCATTCGGTTGAACTGTG |
| *spout1* sgRNA3 PCR-R | CRISPR/Cas9 efficiency | AATTATAATCGTCATGTTCACTGC |
| ***spout1* Mutagenesis** | | |
| Gly98Ser-F | Mutagenesis | GCACCTACTTGGCCAGTCAGATTGCCAGAGC |
| Gly98Ser-R | Mutagenesis | GCTCTGGCAATCTGACTGGCCAAGTAGGTGC |
| Gly244Ser-F | Mutagenesis | CTGCAAGACCTACCATAGCAAAGTGGTATCATC |
| Gly244Ser-R | Mutagenesis | GATGATACCACTTTGCTATGGTAGGTCTTGCAG |
| Gly293Ser-F | Mutagenesis | CGGGACGTCAGAGCGCAGCTCAGATGTGGCCTCT |
| Gly293Ser-R | Mutagenesis | AGAGGCCACATCTGAGCTGCGCTCTGACGTCCCG |
| Asn86Asp-F | Mutagenesis | CTCCATCCTGGACGATGCTCAGTCGCCGGAG |
| Asn86Asp-R | Mutagenesis | CTCCGGCGACTGAGCATCGTCCAGGATGGAG |
| Arg200Trp-F | Mutagenesis | GTGGTGGATCGGCCCACCTGGCCAGGCCACGGCTCC |
| Arg200Trp-R | Mutagenesis | GGAGCCGTGGCCTGGCCAGGTGGGCCGATCCACCAC |
| Arg352Cys-F | Mutagenesis | GTAGCCGTACCATCTGCACGGAGGAAGCCATCCTC |
| Arg352Cys-R | Mutagenesis | GAGGATGGCTTCCTCCGTGCAGATGGTACGGCTAC |
| Ser249del-F | Mutagenesis | CATGGCAAAGTGGTATCACAGGACCCTCGCACCAAAGC (3bp deletion) |
| Ser249del-R | Mutagenesis | GCTTTGGTGCGAGGGTCCTGTGATACCACTTTGCCATG (3bp deletion) |
| Thr289Met-F | Mutagenesis | GACCTGACCATCGGGATGTCAGAGCGCGGCTCAG |
| Thr289Met-R | Mutagenesis | CTGAGCCGCGCTCTGACATCCCGATGGTCAGGTC |
| Tyr339Cys-F | Mutagenesis | CTCTTTGACCTGTGCGTCAATACCTGTCC |
| Tyr339Cys-R | Mutagenesis | GGACAGGTATTGACGCACAGGTCAAAGAG |
| Thr353Met-F | Mutagenesis | GCCGTACCATCCGCATGGAGGAAGCCATCCTC |
| Thr353Met-R | Mutagenesis | GAGGATGGCTTCCTCCATGCGGATGGTACGGC |
| Thr130Arg-F_NC | Mutagenesis | GTGGAGGGGGAATTCAGAGGAGTTGGGAAGAAGG |
| Thr130Arg-R_NC | Mutagenesis | CCTTCTTCCCAACTCCTCTGAATTCCCCCTCCAC |
| **Construct Sequencing primers** | | |
| *spout1*-seq-F1 | construct sequencing | AGGATTGAGTGGCGAAAATG |
| *spout1*-seq-F2 | construct sequencing | CTCCATCCTGGACAATGCTC |
| *spout1*-seq-F3 | construct sequencing | GCTGGTCTCTACTGGGGCTA |
| *spout1*-seq-F4 | construct sequencing | CAACTTCAGGCATGCTCTTGT |
| *spout1*-seq-R1 | construct sequencing | CATCAGCTTGAGATCCTTCCA |
| **qPCR primers** | | |
| *spout1-*Ex3_4-F | Relative gene expression | GCAGCTGGAGAAACAGAAGC |
| *spout1-*Ex3_4-F | Relative gene expression | ATCACTGATCCAGGCAGAGC |
| *spout1-*Ex11_12-R | Relative gene expression | CAGCGTGCTTTTTGACCTTT |
| *spout1-*Ex11_12-R | Relative gene expression | ATGCGATTTTCTGTCGGAGT |

**Table S4: Statistical differences in stable mutant, F0 crispants and transient *spout1/cenp-32* zebrafish models**

|  | N | Median | q vs control | q vs *spout1*^+/-^ or *SPOUT1* WT mRNA | q vs *spout1*^-/-^ or morphants | q vs WT mRNA rescue | Interpretation |
| --- | --- | --- | --- | --- | --- | --- | --- |
| *spout1* mutant head size phenotyping, related to Figure 2B | | | | | | | |
| *spout1^+/+^* head size | 44 | 100 |  | 0.1826 | 0.0001 |  |  |
| *spout1^+/-^* head size | 88 | 99 | 0.1826 |  | 0.0001 |  |  |
| *spout1^-/-^* head size | 47 | 81 |  | 0.0001 | 0.0001 |  |  |
| *spout1* mutant body length phenotyping, related to Figure 2C | | | | | | | |
| *spout1^+/+^* body length | 24 | 100 |  | 0.5593 | 0.0128 |  |  |
| *spout1^+/-^* body length | 44 | 100 | 0.5593 |  | 0.0128 |  |  |
| *Spout1^-/-^* body length | 26 | 98 | 0.0128 | 0.0128 |  |  |  |
| Optic tecta phenotyping in *spout1* mutants, related to Figure 4B | | | | | | | |
| *spout1^+/+^* | 16 | 100 |  | 0.8405 | 0.0001 |  |  |
| *spout1^+/-^* | 32 | 102 | 0.8405 |  | 0.0001 |  |  |
| *spout1^-/-^* | 20 | 42 | 0.0001 | 0.0001 |  |  |  |
| Intertectal neuron phenotyping in *spout1* mutants, related to Figure 4C | | | | | | | |
| *spout1^+/+^* | 15 | 99 |  | 0.7596 | 0.0001 |  |  |
| *spout1^+/-^* | 32 | 99 | 0.7596 |  | 0.0001 |  |  |
| *spout1^-/-^* | 20 | 45 | 0.0001 | 0.0001 |  |  |  |
| TUNEL phenotyping in *spout1* mutants, related to Figure 4E | | | | | | | |
| *spout1^+/+^* | 16 | 100 |  | 0.5276 | 0.0001 |  |  |
| *spout1^+/-^* | 42 | 108 | 0.5276 |  | 0.0001 |  |  |
| *spout1^-/-^* | 23 | 265 | 0.0001 |  | 0.0001 |  |  |
| Cell cycle progression phenotyping in *spout1* mutants, related to Figure 4G | | | | | | | |
| *spout1^+/+^* | 11 | 100 |  | 0.8887 | 0.0013 |  |  |
| *spout1^+/-^* | 29 | 103 | 0.8887 |  | 0.0001 |  |  |
| *spout1^-/-^* | 21 | 127 | 0.0013 | 0.0001 |  |  |  |
| Variant tested (head size phenotyping), related to Figure 3B | | | | | | | |
| UC | 372 | 100 |  |  | 0.0001 |  |  |
| MO | 325 | 90 |  |  |  | 0.0001 |  |
| MO + WT mRNA | 305 | 99 |  |  | 0.0001 |  |  |
| MO + p.T130R | 62 | 100 |  |  | 0.0001 | 0.4322 | Benign |
| MO + p.G98S | 132 | 90 |  |  | 0.6922 | 0.0001 | Pathogenic |
| MO + p.G244S | 98 | 93 |  |  | 0.0001 | 0.0001 | Pathogenic |
| MO + p.G293S | 75 | 94 |  |  | 0.0005 | 0.0001 | Pathogenic |
| MO + p.N86D | 117 | 89 |  |  | 0.7677 | 0.0001 | Pathogenic |
| MO + p.R200W | 82 | 93 |  |  | 0.0004 | 0.0001 | Pathogenic |
| MO + p.R352C | 82 | 93 |  |  | 0.0089 | 0.0001 | Pathogenic |
| MO + p.S249del | 66 | 94 |  |  | 0.0001 | 0.0001 | Pathogenic |
| MO + p.T289M | 66 | 95 |  |  | 0.0001 | 0.0001 | Pathogenic |
| MO + p.T353M | 66 | 95 |  |  | 0.0001 | 0.0001 | Pathogenic |
| MO + p.Y339C | 83 | 95 |  |  | 0.0001 | 0.0001 | Pathogenic |
| *spout1* F0 head size phenotyping, related to Figure S4C | | | | | | | |
| UC | 102 | 100 |  |  |  |  |  |
| sgRNA1 | 90 | 99 | 0.1228 |  |  |  |  |
| sgRNA1 + Cas9 | 60 | 87 | 0.0001 |  |  |  |  |
| *spout1* F0 body length phenotyping, related to Figure S4E | | | | | | | |
| UC | 95 | 100 |  |  |  |  |  |
| sgRNA1 | 84 | 100 | 0.7929 |  |  |  |  |
| sgRNA1 + Cas9 | 60 | 102 | 0.0084 |  |  |  |  |
| *spout1* F0 head size phenotyping, related to Figure S4D | | | | | | | |
| UC | 129 | 100 |  |  |  |  |  |
| sgRNA3 | 90 | 99 | 0.0249 |  |  |  |  |
| sgRNA3 + Cas9 | 112 | 83 | 0.0001 |  |  |  |  |
| *spout1* F0 body length phenotyping, related to Figure S4F | | | | | | | |
| UC | 128 | 100 |  |  |  |  |  |
| sgRNA3 | 90 | 101 | 0.0004 |  |  |  |  |
| sgRNA3 + Cas9 | 112 | 100 | 0.8969 |  |  |  |  |
| CNS phenotyping in *spout1* transient knockdown zebrafish, related to Figure S6B | | | | | | | |
| UC | 73 | 101 |  |  | 0.0001 | 0.0001 |  |
| MO | 74 | 76 | 0.0001 |  |  | 0.0001 |  |
| MO + WT mRNA | 67 | 94 | 0.0001 |  | 0.0001 |  |  |
| CNS phenotyping in *spout1* transient knockdown zebrafish, related to Figure S6C | | | | | | | |
| UC | 73 | 101 |  |  | 0.0001 | 0.0118 |  |
| MO | 74 | 77 | 0.0001 |  |  | 0.0001 |  |
| MO + WT mRNA | 67 | 96 | 0.0118 |  | 0.0001 |  |  |
| Apoptosis phenotyping in *spout1* transient suppression zebrafish model, related to Figure S6E | | | | | | | |
| UC | 60 | 98 |  |  | 0.0001 | 0.0043 |  |
| MO | 50 | 308 | 0.0001 |  |  | 0.0001 |  |
| MO + WT mRNA | 48 | 114 | 0.0043 |  | 0.0001 |  |  |
| Cell cycle progression phenotyping in *spout1* transient suppression zebrafish model, related to Figure S6G | | | | | | | |
| UC | 61 | 99 |  |  | 0.0001 | 0.5685 |  |
| MO | 62 | 149 | 0.0001 |  |  | 0.0001 |  |
| MO + WT mRNA | 61 | 101 | 0.5685 |  | 0.0001 |  |  |
| Morpholino dose curve (head size phenotyping) related to Supplementary Figure S5B | | | | | | | |
| UC | 90 | 100 |  |  |  |  |  |
| 3ng MO | 82 | 96 | 0.0001 |  |  |  |  |
| 6ng MO | 76 | 83 | 0.0001 |  |  |  |  |
| 9ng MO | 85 | 84 | 0.0001 |  |  |  |  |
| Rescue dose curve (head size phenotyping) related to Supplementary Figure S5D | | | | | | | |
| UC | 75 | 100 |  |  | 0.0001 |  |  |
| MO | 69 | 87 | 0.0001 |  |  |  |  |
| MO + 50pg WT mRNA | 67 | 94 | 0.0001 |  | 0.0001 |  |  |
| MO + 100pg WT mRNA | 58 | 96 | 0.0001 |  | 0.0001 |  |  |
| MO + 150pg WT mRNA | 63 | 96 | 0.0001 |  | 0.0001 |  |  |
| Variant tested (body length phenotyping), related to Supplementary Figure S9B | | | | | | | |
| UC | 363 | 100 |  |  |  |  |  |
| MO | 323 | 99 |  |  |  | 0.0004 |  |
| MO + WT mRNA | 304 | 100 |  |  | 0.0004 |  |  |
| MO + p.T130R | 63 | 101 |  |  | 0.0009 | 0.2002 |  |
| MO + p.G98S | 98 | 98 |  |  | 0.0005 | 0.0001 |  |
| MO + p.G244S | 98 | 100 |  |  | 0.001 | 0.4009 |  |
| MO + p.G293S | 75 | 100 |  |  | 0.8524 | 0.0458 |  |
| MO + p.N86D | 117 | 97 |  |  | 0.0001 | 0.0001 |  |
| MO + p.R200W | 82 | 100 |  |  | 0.0655 | 0.6663 |  |
| MO + p.R352C | 84 | 101 |  |  | 0.0001 | 0.1236 |  |
| MO + p.S249del | 66 | 101 |  |  | 0.0001 | 0.0132 |  |
| MO + p.T289M | 64 | 102 |  |  | 0.0001 | 0.0003 |  |
| MO + p.T353M | 101 | 99 |  |  | 0.4332 | 0.0012 |  |
| MO + p.Y339C | 82 | 101 |  |  | 0.0008 | 0.2862 |  |
| Variants tested (head size Phenotyping ), related to Supplementary Figure S10A | | | | | | | |
| UC | 89 | 100 |  |  |  |  |  |
| MO | 62 | 91 |  |  |  |  |  |
| MO + WT mRNA | 74 | 99 |  |  |  |  |  |
| MO + T130R | 62 | 97 |  |  |  |  |  |
| MO + G293S | 75 | 94 |  |  |  |  |  |
| MO + R200W | 82 | 93 |  |  |  |  |  |
| Variants tested (body length phenotyping), related to Supplementary Figure S10B | | | | | | | |
| UC | 89 | 100 |  |  |  |  |  |
| MO | 61 | 100 |  |  |  |  |  |
| MO + WT mRNA | 74 | 100 |  |  |  |  |  |
| MO + T130R | 63 | 101 |  |  |  |  |  |
| MO + G293S | 75 | 100 |  |  |  |  |  |
| MO + R200W | 82 | 100 |  |  |  |  |  |
| Variants tested (head size phenotyping), related to Supplementary Figure S10C | | | | | | | |
| UC | 91 | 101 |  |  |  |  |  |
| MO | 88 | 91 |  |  |  |  |  |
| MO + WT mRNA | 76 | 100 |  |  |  |  |  |
| MO + R352C | 82 | 93 |  |  |  |  |  |
| MO + G244S | 98 | 93 |  |  |  |  |  |
| MO + L314F | 106 | 95 |  |  |  |  |  |
| Variants tested (body length phenotyping), related to Supplementary Figure S10D | | | | | | | |
| UC | 93 | 100 |  |  |  |  |  |
| MO | 88 | 100 |  |  |  |  |  |
| MO + WT mRNA | 76 | 101 |  |  |  |  |  |
| MO + R352C | 84 | 101 |  |  |  |  |  |
| MO + G244S | 98 | 100 |  |  |  |  |  |
| MO + L314F | 105 | 101 |  |  |  |  |  |
| Variants tested (head size phenotyping), related to Supplementary Figure S10E | | | | | | | |
| UC | 111 | 100 |  |  |  |  |  |
| MO | 95 | 88 |  |  |  |  |  |
| MO + WT mRNA | 84 | 96 |  |  |  |  |  |
| MO + G98S | 103 | 90 |  |  |  |  |  |
| MO + T353M | 101 | 90 |  |  |  |  |  |
| MO + N86D | 117 | 89 |  |  |  |  |  |
| Variants tested (body length phenotyping), related to Supplementary Figure S10F | | | | | | | |
| UC | 110 | 100 |  |  |  |  |  |
| MO | 95 | 98 |  |  |  |  |  |
| MO + WT mRNA | 85 | 100 |  |  |  |  |  |
| MO + G98S | 98 | 98 |  |  |  |  |  |
| MO + T353M | 95 | 99 |  |  |  |  |  |
| MO + N86D | 117 | 97 |  |  |  |  |  |
| Variants tested (head size phenotyping), related to Supplementary Figure S10G | | | | | | | |
| UC | 81 | 100 |  |  |  |  |  |
| MO | 80 | 92 |  |  |  |  |  |
| MO + WT mRNA | 71 | 100 |  |  |  |  |  |
| MO + T289M | 66 | 95 |  |  |  |  |  |
| MO + Y339C | 83 | 95 |  |  |  |  |  |
| MO + S249del | 66 | 94 |  |  |  |  |  |
| Variants tested (body length phenotyping), related to Supplementary Figure S10H | | | | | | | |
| UC | 71 | 100 |  |  |  |  |  |
| MO | 78 | 100 |  |  |  |  |  |
| MO + WT mRNA | 69 | 101 |  |  |  |  |  |
| MO + T289M | 64 | 102 |  |  |  |  |  |
| MO + Y339C | 82 | 101 |  |  |  |  |  |
| MO + S249del | 66 | 101 |  |  |  |  |  |
| Overexpression (head size phenotyping), related to Supplementary Figure S11A | | | | | | | |
| UC | 236 | 100 |  |  |  |  |  |
| WT mRNA | 49 | 100 | 0.7094 |  |  |  |  |
| p.T130R | 53 | 100 | 0.7094 | 0.7094 |  |  |  |
| p.G98S | 74 | 100 | 0.8857 | 0.7094 |  |  |  |
| p.G244S | 77 | 100 | 0.8857 | 0.7094 |  |  |  |
| p.G293S | 62 | 100 | 0.7094 | 0.7094 |  |  |  |
| p.N86D | 76 | 100 | 0.7094 | 0.7094 |  |  |  |
| p.R200W | 51 | 99 | 0.7094 | 0.8857 |  |  |  |
| p.R352C | 73 | 100 | 0.7094 | 0.7094 |  |  |  |
| p.S249del | 73 | 100 | 0.8857 | 0.7094 |  |  |  |
| p.T289M | 86 | 100 | 0.8857 | 0.7094 |  |  |  |
| p.Y339C | 73 | 100 | 0.8857 | 0.7094 |  |  |  |
| p.T353M | 74 | 100 | 0.7094 | 0.8857 |  |  |  |
| Overexpression (body length phenotyping), related to Supplementary Figure S11B | | | | | | | |
| UC | 223 | 100 |  |  |  |  |  |
| WT mRNA | 48 | 101 | 0.348 |  |  |  |  |
| p.T130R | 53 | 101 | 0.2927 | 0.9395 |  |  |  |
| p.G98S | 74 | 100 | 0.6678 | 0.2829 |  |  |  |
| p.G244S | 74 | 100 | 0.5361 | 0.2784 |  |  |  |
| p.G293S | 62 | 100 | 0.8074 | 0.6182 |  |  |  |
| p.N86D | 71 | 101 | 0.2706 | 0.9395 |  |  |  |
| p.R200W | 53 | 101 | 0.8074 | 0.6932 |  |  |  |
| p.R352C | 71 | 100 | 0.8074 | 0.3638 |  |  |  |
| p.S249del | 71 | 100 | 0.4745 | 0.8039 |  |  |  |
| p.T289M | 80 | 100 | 0.259 | 0.0723 |  |  |  |
| p.T353M | 70 | 101 | 0.0009 | 0.2829 |  |  |  |
| p.Y339C | 72 | 101 | 0.8039 | 0.6182 |  |  |  |

**Table S5: Data collection and refinement statistics for SPOUT1/CENP-32 crystal structure.**

|  | **CENP-32-SAH bound** | **CENP-32-SAM bound** | **CENP-32-A356N** |
| --- | --- | --- | --- |
| **Wavelength** |  |  |  |
| **Resolution range** | 25.52 - 2.38 (2.53 - 2.38) | 55.01 - 2.62 (2.88 - 2.62) | 37.62 - 2.5 (2.69 - 2.5) |
| **Space group** | C 2 2 21 | C 2 2 21 | C 2 2 21 |
| **Unit cell** | 75.02 81.17 135.5 90 90 90 | 75.07 80.83 135.69 90 90 90 | 75.24 80.694 136.363 90 90 90 |
| **Total reflections** | 215581 (36235) | 82646 (20351) | 63879 (12747) |
| **Unique reflections** | 16700 (2680) | 12626 (3066) | 14321 (2840) |
| **Multiplicity** | 12.9 (13.5) | 6.5 (6.6) | 4.5 (4.5) |
| **Completeness (%)** | 98.55 (96.30) | 99.08 (98.02) | 97.33 (98.30) |
| **Mean I/sigma(I)** | 21.20 (4.12) | 16.11 (3.81) | 11.22 (2.72) |
| **Wilson B-factor** | 61.84 | 72.90 | 67.46 |
| **R-merge** | 0.07095 (0.6979) | 0.08553 (0.6634) | 0.06425 (0.4695) |
| **R-meas** | 0.07407 (0.7258) | 0.0932 (0.7196) | 0.07257 (0.5331) |
| **R-pim** | 0.0209 (0.1975) | 0.0365 (0.2761) | 0.03278 (0.2461) |
| **CC1/2** | 0.999 (0.971) | 0.998 (0.947) | 0.999 (0.963) |
| **CC*** | 1 (0.993) | 1 (0.986) | 1 (0.991) |
| **Reflections used in refinement** | 16700 (2680) | 12626 (3066) | 14321 (2840) |
| **Reflections used for R-free** | 846 (156) | 628 (153) | 711 (146) |
| **R-work** | 0.2246 (0.3099) | 0.2145 (0.3533) | 0.2113 (0.3522) |
| **R-free** | 0.2621 (0.3866) | 0.2799 (0.4047) | 0.2723 (0.4517) |
| **CC(work)** | 0.954 (0.878) | 0.934 (0.432) | 0.915 (0.551) |
| **CC(free)** | 0.946 (0.733) | 0.888 (0.409) | 0.882 (0.293) |
| **Number of non-hydrogen atoms** | 2201 | 2180 | 2205 |
| **macromolecules** | 2201 | 2178 | 2204 |
| **ligands** | 0 | 0 | 0 |
| **solvent** | 0 | 2 | 1 |
| **Protein residues** | 285 | 283 | 286 |
| **RMS(bonds)** | 0.007 | 0.010 | 0.011 |
| **RMS(angles)** | 0.93 | 1.67 | 1.75 |
| **Ramachandran favored (%)** | 94.29 | 93.53 | 93.97 |
| **Ramachandran allowed (%)** | 5.36 | 3.60 | 5.67 |
| **Ramachandran outliers (%)** | 0.36 | 2.88 | 0.35 |
| **Rotamer outliers (%)** | 4.31 | 6.64 | 3.39 |
| **Clashscore** | 9.42 | 10.69 | 5.96 |
| **Average B-factor** | 88.78 | 86.17 | 85.80 |
| **macromolecules** | 88.78 | 86.47 | 85.81 |
| **ligands** |  | 63.54 |  |
| **solvent** |  | 60.38 | 50.86 |

Statistics for the highest-resolution shell are shown in parentheses.

**Table S6:** Plasmids and guide RNA sequences designed for generating KO cells using the CRISPR-Cas9 system.

Plasmid	Target gene	Sequence (5’ → 3’)

pTORA14AAVS1	*AAVS1*	GGGGCCACUAGGGACAGGAU

**Table S7:** Sequences of the primers used for SPOUT1/CENP-32-GFP stable cell line studies.

Primer	Sequence (5’ → 3’)

o-EGFP_MK243	tcgcggccgcacgcgtgccACcatggtgagcaagggc

o-DEST131_RV_pMK243	tatgctgcagagatcCCtCTAgaCTActTTAgacc

o-DEST243_CENP-32_FW71	tcgcggccgcacgcgagatctATGCGGCCCTACACACTGAGCG

o-DEST243_GFP_RV1	agtcactatggtcgagtcgcggccgctttacttg

o-AAVS1_FW	attgtcactttgcgctgccc

o-AAVS1_RV2	aagagtgagtttgcccaagc

pMK243_mut_detect_RV	gaccttgatatgctgcctgc
